# Medication Against Conflict

**DOI:** 10.1101/2022.03.22.22272752

**Authors:** Andrea Berlanda, Matteo Cervellati, Elena Esposito, Dominic Rohner, Uwe Sunde

## Abstract

The consequences of successful public health interventions for social violence and conflict are largely unknown. This paper closes this gap by evaluating the effect of a major health intervention – the successful expansion of anti-retroviral therapy (ART) to combat the HIV/AIDS pandemic – in Africa. To identify the effect, we combine exogenous variation in the scope for treatment and global variation in drug prices. We find that the ART expansion significantly reduced the number of violent events in African countries and sub-national regions. The effect pertains to social violence and unrest, not civil war. The evidence also shows that the effect is not explained by general improvements in economic prosperity, but related to health improvements, greater approval of government policy, and increased trust in political institutions. Results of a counterfactual simulation reveal the largest potential gains in countries with intermediate HIV prevalence where disease control has been given relatively low priority.

JEL-classification: C36, D47, I15, O10

## 1 Introduction

Adverse health conditions and social violence constitute major problems for developing countries. This is particularly the case in Africa, which is plagued by widespread social violence, insecurity, and unrest. At the same time, Africa is particularly affected the spread of communicable diseases as reflected by, e.g., the HIV/AIDS pandemic. Around 24 million people in Africa still live with HIV, with an annual death toll of half a million and a prevalence of a fourth of the population in some countries. Besides the serious health consequences, the HIV pandemic led to lower labor productivity, increased expenditures for medication and assistance, and increased poverty. Around the turn of the millennium, following the massive increase in HIV prevalence and in view of projections of global infections and mortality, scientists and international organizations raised serious concerns regarding the grim outlook for the socio-economic consequences of the pandemic. This included warnings of the risk of widespread anger, social unrest and violence in the face of inadequate health policies (Birmingham, 2000; Kumar, 2000; Fourie and Schönteich, 2001; Feldbaum et al., 2006; de Waal, 2010). International organizations emphasized repeatedly that the dissatisfaction with government policies and the erosion of trust in institutions associated with this dissatisfaction potentially plays an important role for the failure of many countries in building peaceful and inclusive societies (see, e.g., OECD, 2017; United Nations, 2021). However, whether major health interventions can help to reduce conflict and social violence remains largely unknown.

The HIV epidemic provides a unique laboratory to investigate this question. Soon after the turn of the millennium, a massive worldwide roll-out of antiretroviral therapy (ART) for HIV-positive individuals followed as consequence of a substantial decline in the costs of drug production. This health intervention yielded substantial health improvements, reductions in mortality, and a recovery of labor productivity. Yet, surprisingly little is known about the consequences of major health interventions, such as the ART roll-out, for social violence. Existing empirical studies have focused on population dynamics, weak institutions, ethnic tensions, natural resource competition, income and commodity price shocks, short-term weather driven shocks, and climate as root causes of conflict and social violence. The findings of these studies often have no clear implications for policy, or policy implications that are difficult to implement. Only recently, some empirical work has pointed at the potential role of health shocks for outbreaks of civil conflict, but without analyzing the effect of public health interventions. At the same time, public health interventions, such as the roll-out of ART, were very successful in improving public health. To the extent that labor productivity and opportunity costs play important roles for explaining incentives to engage in protests, riots and other forms of social violence, the success of such public health interventions may reduce social grievances. Hence, besides restoring labor productivity and fostering individual medical and economic well-being, public health interventions might remove important factors behind protests and social violence and help restoring confidence and trust in institutions and the government. However, evidence for the hypothesis that major health interventions like the ART expansion reduce conflict is still missing.

This paper closes this gap in the literature by investigating whether health interventions play a potentially relevant role in reducing conflict and social violence. To address this question, we perform a first systematic empirical investigation of the effects of a large-scale health policy, the ART roll-out to combat HIV/AIDS, on violent events in Africa. We ask whether, by successfully combating the HIV/AIDS pandemic in Africa, the ART expansion also led to a reduction in conflict and social violence, and, if so, through which channels. The identification strategy is based on exogenous variation in the exposure to the ART expansion in a country or sub-national region. We filter out time-invariant confounders and global shocks, and draw on an identification strategy that combines cross-sectional variation in the potential for ART treatment (based on HIV prevalence before the availability of ART) with time variation in the access to ART at the global level (based on the global variation in prices and production costs of ART that was driving the dynamics of ART coverage worldwide). The identification strategy is implemented using different measures to construct this interaction, and the analysis is conducted with a variety of estimation methods and robustness checks.

We find robust evidence that the expansion of ART coverage led to a significant reduction in the number of violent events in African countries and sub-national regions. This reduction pertains, in particular, to riots and demonstrations related to economic and human rights motives, but not to large scale armed conflict. The effect works partly through a reduction in economic grievances, but does not merely reflect an improvement in overall economic well-being. In particular, we find evidence for an independent effect of health interventions that does not work through economic well-being per se. A large set of potential confounds can be ruled out as driving the result. An analysis of potential channels reveals that the expansion of ART was associated with increase in individual trust in institutions like the parliament and the local government, and with an increase in individual approval of government policies related to the management of HIV, of basic health provision, and economics in general, but not with policies related to education. Taken together, these findings imply that ill health may be a potent driver of social unrest and violence, and that besides improving health and economic conditions, public health interventions can also help curbing social violence. Results of a counterfactual simulation analysis suggest that efforts to increase the ART coverage would have had the largest quantitative potential gains in African countries with intermediate HIV prevalence and in which HIV treatment has been given relatively low priority.

Our analysis contributes to the recent literature in several ways. It has become increasingly clear that social violence, in the form of protests and riots, rather than civil war, constitutes the vast majority of conflict events in Africa and thereby poses a major impediment for development (see, e.g., Straus, 2012). A growing body of research has provided evidence that variation in productivity and opportunity costs is relevant for explaining the incentives to engage in protests, riots and other forms of social violence. Existing empirical studies on the root causes of social violence have focused on income and commodity price shocks (Dube and Vargas, 2013; Bazzi and Blattman, 2014; McGuirk and Burke, 2020; Berman et al., 2021), short-term weather driven shocks (Miguel et al., 2004; Dell et al., 2014; König et al., 2017; Harari and La Ferrara, 2018), and climate (Theisen et al., 2013; Burke et al., 2015; Breckner and Sunde, 2019). Related work has pointed at the role of weak institutions (Besley and Persson, 2011), ethnic tensions (Esteban et al., 2012), natural resource competition (Caselli et al., 2015; Berman et al., 2017; Rohner, 2018) and population (Acemoglu et al., 2020). Few recent studies have found evidence for weather-related health shocks as a previously largely overlooked cause of social violence (Cervellati et al., 2017, 2021). Evidence for the impact of public health interventions, and an evaluation of whether and through which channels public health interventions affect social violence, however, is largely missing. Our paper fills this gap.

Addressing the role of major health interventions is of foremost importance from a policy perspective. To our knowledge, there exists no systematic evaluation of the effects of major health interventions on social conflict. This way, our work complements research on the effects of policy for social violence, which has considered foreign aid (de Ree and Nillesen, 2009; Savun and Tirone, 2012; Nunn and Qian, 2014), cash transfers (Crost et al., 2014), infrastructure investments (Berman et al., 2011), reconciliation (Ciliers et al., 2016), and employment policies (Blattman and Annan, 2016; Fetzer, 2020), but which has neglected the role of health interventions. In fact, the results of the existing literature suggest that the effects of policy interventions to prevent or reduce social violence were generally mixed, and policies that resulted in the disbursement of appropriable cash were generally much less successful than policies that led to a higher opportunity cost of fighting (see, e.g., Rohner and Thoenig, 2020, for a survey). This is exactly what health interventions accomplish, so our evidence contributes an important missing piece of evidence regarding the scope of policy interventions against conflict. Our findings also provide evidence that support arguments that health interventions help fostering trust in states and policies (see, e.g., Khemani, 2020). On a more general level, the result that health interventions also help reducing social violence implies that taking into account additional societal and economic benefits beyond pure individual health effects is key for appraising the impact of public health policy. The findings therefore complement findings of purely economic effects of major health interventions such as the ART expansion (see, e.g., Tompsett, 2020) and suggest the usefulness of a broad assessment of the overall benefits of other health policies, such as, for example, extending the global availability of COVID-19 vaccination.

From the perspective of political economy, the findings indicate that public health interventions are a key source of political legitimacy for institutions and incumbent governments, with potential implications for other public investments in health. In particular, our findings complement recent evidence on the role of economic hardship and public support for democracies as well as the rise of populism (e.g., Algan et al., 2017; Claassen, 2020). Evidence for public health interventions helping to restore trust in government contributes a new aspect to the literature on the role of state performance for political trust (see, e.g., Citrin and Stoker, 2018, for a survey) and is consistent with recent evidence for the effect of life expectancy on democratic attitudes (Lechler and Sunde, 2019). In light of recent calls by international organizations for the need of fostering trust in institutions in order to maintain economic and political security (OECD, 2017; United Nations, 2021), our results are informative about policies that are effective in this dimension.

The remainder of the paper is organized as follows. Section 2 describes the data and the empirical methodology. Section 3 presents the empirical results, robustness checks, and evidence for the underlying mechanisms. Section 4 concludes with a discussion of the policy implications.

## 2 Data and Empirical Strategy

### 2.1 Background: HIV and ART in Africa

The human immunodeficiency virus (HIV) impairs the function of white blood cells in the immune system (CD4 cells) and replicates itself inside these cells. As consequence, infected individuals experience a weakening of the immune system, making the body vulnerable to infections and some types of cancer. In advanced stages, the infection turns into the acquired immunodeficiency syndrome (AIDS), which ultimately leads to death. Recent evidence suggests that the most infective strain of HIV crossed from chimpanzees to humans probably before 1920 in Cameroon, while the beginning of the spread of HIV across Africa has been traced back to Kinshasa, located in today’s Democratic Republic of Congo, around 1920 (Gao et al., 1999; Faria et al., 2014). From the late 1970s onwards, the spread of HIV turned into an epidemic that swept across Africa and the entire world. By 1980, about half of human infections in the Democratic Republic of Congo were observed outside of Kinshasa. In Africa, the virus subsequently diffused out of the Democratic Republic of Congo, first towards the great lakes area and then along the East of Africa, eventually reaching the Mediterranean basin and South Africa as well as the North-West towards Nigeria during the 1990’s (Kalipeni and Zulu, 2012). By 2000, an estimated 26 million adults and children lived with HIV/AIDS in Africa, constituting more than 70 percent of the global infections.^1^

During the early 1980s, the then still unknown disease rapidly spread across the world, leading to the first clinical and epidemiological observations of AIDS in 1981. The severity led to intense microbiological research. During the early 1980s, the retrovirus responsible for AIDS (the HIV-1 virus) was isolated successfully, and subsequent discoveries concerned the transmission and life cycle of HIV. The identification of the main receptors of the HIV led to the development of combination antiretroviral therapy (ART) in the late 1990s (see, e.g., Barré-Sinoussi et al., 2013, for a survey of the history of HIV research). Parallel to the scientific advances, campaigns to inhibit a further spread of HIV and the development and widespread distribution of drugs to treat HIV/AIDS also became important issues on the political agendas of national governments and international organizations. This led to mounting pressure by international non-government organizations (NGOs) on pharmaceutical companies to no longer prevent the distribution of generics, which culminated in the introduction of generic drugs for antiretroviral therapy in 2001. The subsequent expansion of the availability of ART, which was heavily supported by the Global Fund, led to a significant reduction in morbidity and mortality and a restoration of immunity in infected persons. Ultimately, the availability of ART transformed HIV infections from fatal to a manageable chronic disease with moderate implications for life expectancy if treated appropriately, and international organizations and NGOs continue to exert great effort on expanding ART coverage, particularly in Africa.

### 2.2 Data

The analysis is based on observational data at the country level for 50 African countries and at sub-national administrative level 1 for 170 regions over 18 African countries over the period 1990–2017.

We use geo-localized data for events of social violence from multiple sources. The baseline analysis uses the Social Conflict Analysis Database (SCAD), which is a compilation of violent events that is based on global press coverage. The SCAD data represents a complete and extensive measure of social violence of different forms (protests, demonstrations, riots, strikes, and other forms of social disturbances) and comprises a classification of different event types, including organized events, spontaneous events, and events related to elections, economic grievances, or human rights. In addition, the data contain information on event sizes, in terms of casualties and participants. Based on the narratives contained in the data set, it is also possible to isolate different event types related to targets or actors involved in events (such as NGOs, health workers, or civil servants), which allows a detailed investigation of the mechanisms underlying outbreaks of social violence. The main advantage of the SCAD data in comparison to other frequently used sources of civil conflict is that it focuses on social violence defined as social and political unrest, as opposed to large-scale organized armed conflicts. Moreover, the SCAD data are available for long time periods and exhibit high data quality.

For robustness, we also use data on riots and protests from alternative sources such as the Armed Conflict Location and Event Data (ACLED) and data on organized violence involving the state collected by the Uppsala Conflict Data Program (UCDP). Detailed descriptions of the various data sources, the definitions of violent events, and the different aspects covered by the different data sets are contained in the Supplementary Appendix. Data for HIV prevalence and Antiretroviral Therapy (ART) coverage at the country level are provided by UNAIDS for 50 African countries. HIV prevalence information is based on model estimates by UNAIDS, which collects all country estimates and reviews them in order to guarantee that estimates are comparable across regions and countries over time. Information on ART coverage is based on national registers of antiretroviral therapy, and is compiled by UNAIDS. Regional HIV prevalence (at administrative level 1) is constructed by us using survey data from the Demographic and Health Survey Program (DHS), which represents a comprehensive source of sub-national information to map HIV prevalence. We assembled sub-national level measures of HIV prevalence for 170 regions over 18 African countries (see Figures 1 and A1 in the Supplementary Appendix).

**Figure 1:**
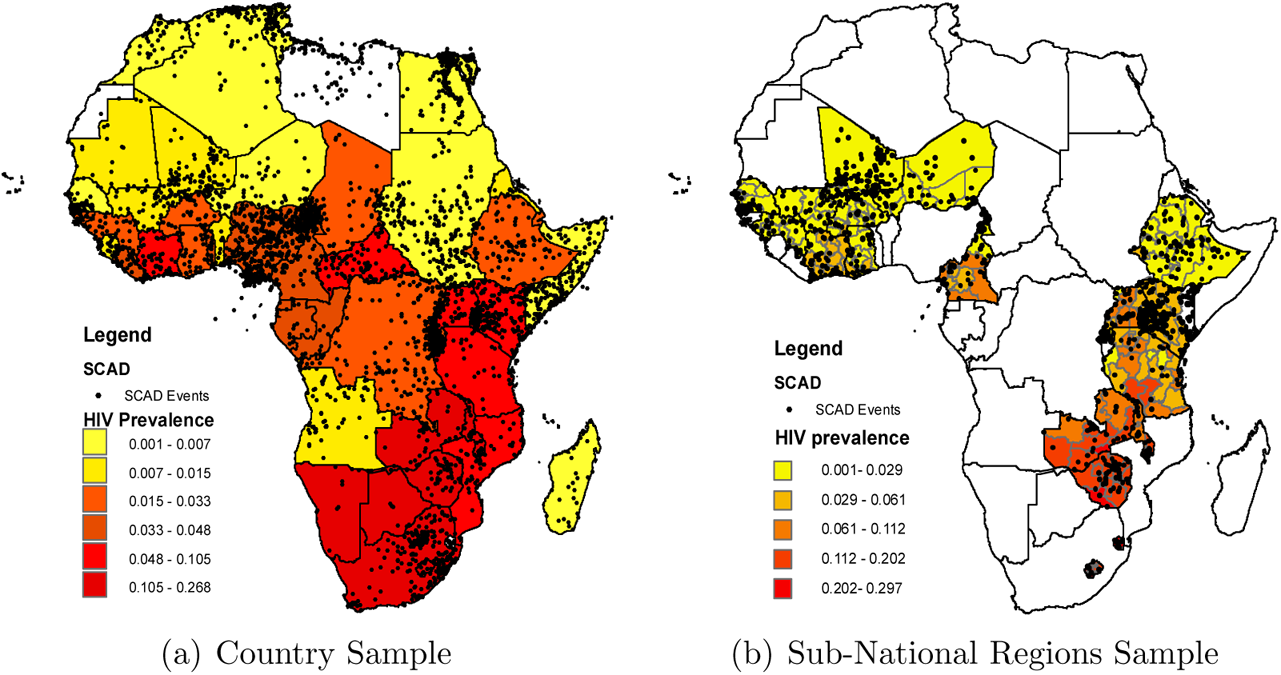
HIV Prevalence and Social Violence in Africa: SCAD *Note:* Panel (a): HIV prevalence in 2001 and location of violent events (SCAD) aggregated at the country level. Panel (b): HIV prevalence and location of violent events (SCAD) at the level of sub-national regions as contained in the sample. See Appendix A.3.1 for summary statistics.

The identification strategy makes use of data on the global price of the first line of combined ART treatment as well as of information about the costs of the active pharmaceutical ingredients used in the production of these treatment lines. The respective data have been collected by the WHO Global Price Reporting Mechanism and by the Global Fund Pooled Procurement Mechanism Reference Pricing and cover approximately 70-80 percent of the global transactions of the respective drugs.

Additional data on population and life expectancy at birth is provided by the UN Population Division and various Census reports. Information on GDP (in constant 2010 US✩) is from the World Bank. Information on trust in institutions is extracted from individual survey responses from the Afrobarometer.

Detailed information about data sources, variable definitions and the construction of the variables used for estimation as well as of the construction of the samples at the country-level and at sub-national level is contained in the Supplementary Appendix (Section A.1).

### 2.3 Empirical Approach: Graphical Illustration

Figure 1 shows a map of HIV prevalence in 2001 and the distribution violent events (SCAD) aggregated over the entire observation period for the country sample and for the sample of sub-national regions. The figure illustrates a geographical distribution of HIV prevalence that exhibits higher levels in sub-Saharan Africa and in the South-East of the continent. The visual inspection suggests a correlation between HIV prevalence and violent events.

Figure 2(a) plots the increase of HIV prevalence in Africa during the 1990s as depicted by the solid line, as well as the expansion of ART during the early 2000s (dashed line), which led to a reversal in the dynamics of HIV infections. This reversal prevented an estimated 9.5 million deaths and brought considerable economic benefits (Forsythe et al., 2019), mainly by contributing to a substantial reduction in mortality (Bor et al., 2013; Tompsett, 2020) and leading to a recovery of labor productivity (Habyarimana et al., 2010; Bor et al., 2012; Baranov et al., 2015). Figure 2(b) illustrates that the increase in HIV prevalence during the 1990s was associated with a relative increase in social violence in countries with high HIV prevalence in comparison to countries with low HIV prevalence. This relative increase in violent events in high HIV countries peaked during the early 2000s and was followed by a reversal that coincided with the ART expansion. The empirical analysis below investigates the hypothesis that the expansion in ART was causally responsible for the relative decline in social violence.

**Figure 2:**
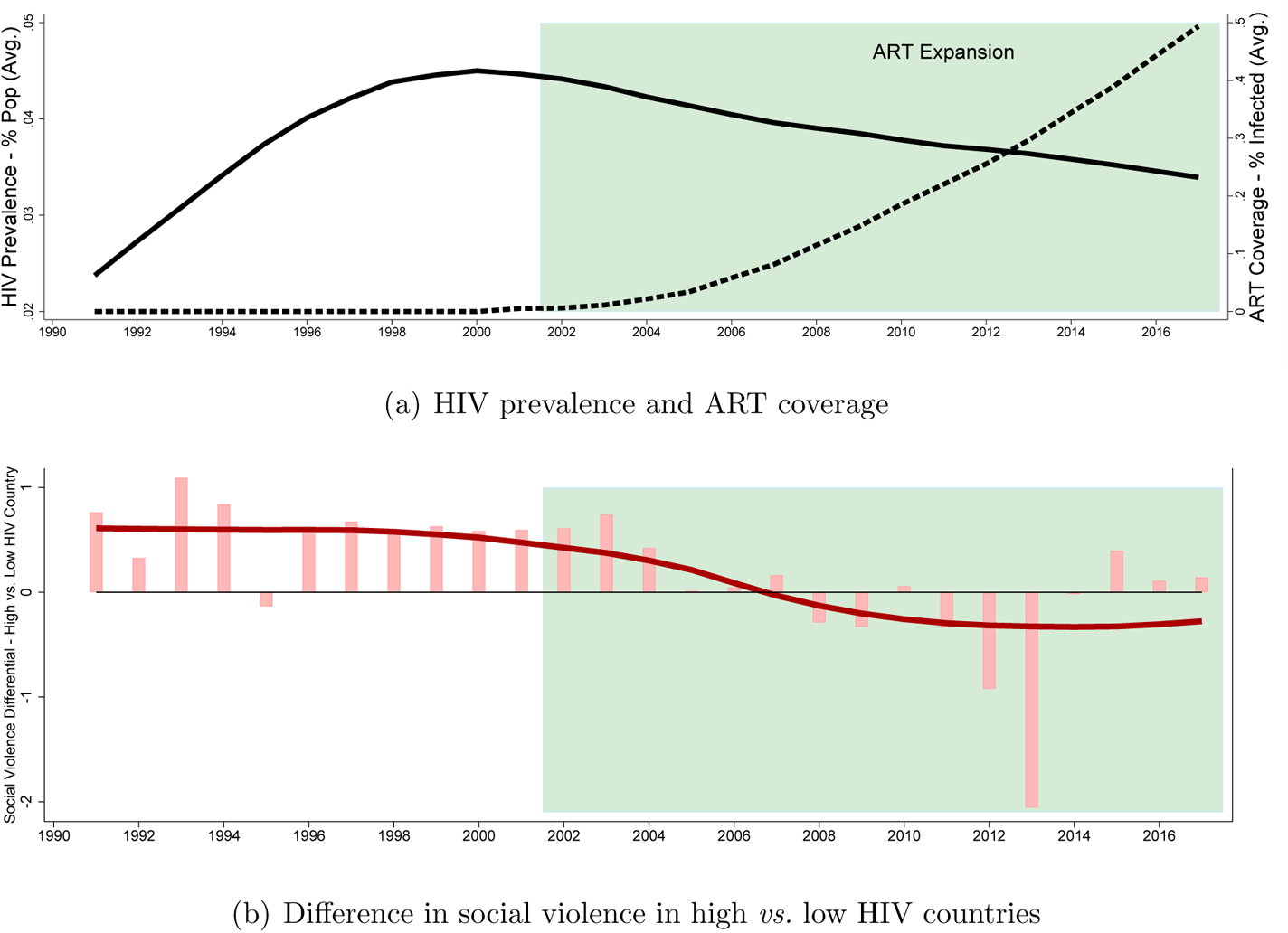
HIV Prevalence, ART Coverage, and Social Violence in Africa *Note:* Panel (a): Evolution of average HIV prevalence as percentage of the population in Africa (solid line), and of average ART coverage as percentage of infected population (dashed line); data from UNAIDS, averages weighted by country population. Panel (b): Difference in social violence in countries with HIV prevalence above and below the median in 2001 (bars, based on SCAD database) and non-linear time trend in terms of log polynomial smooth (line). Average social violence is weighted by country population; differences in high and low HIV countries are normalized by average population-weighted social violence for Africa in each year.

### 2.4 Estimation and Identification

#### Estimation

The empirical analysis is based on the model

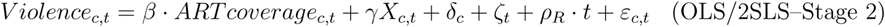

which is a regression of social violence in country *c* and year *t* on ART coverage, control variables *X_c,t_*, country fixed effects *δ_c_*, year fixed effects *ζ_t_*, African-subregion-specific linear time trends *ρ_R_ · t*, and an error term *ε*.

The identification of the effect of ART coverage on social violence, *β*, relies on the assumption that ART coverage is uncorrelated with unobserved or omitted confounding factors contained in the error term *ε*. Violation of this assumption leads to biased estimates and may materialize in a spurious effect. In particular, this would be the case if ART coverage and the level of social violence are both correlated with unobserved variables that create problems of omitted variables or reverse causality. Examples for omitted confounders include institutions: if countries with better institutions and public governance, or just a better economic performance, are more effective in providing health services and ART coverage and, at the same time, more effective in reducing social tensions and violence, this would imply simultaneity bias. Examples for reverse causality include political pressure: if social violence in terms of strikes and demonstrations in a country leads to an intensified effort to treat HIV by governments, international organizations or international aid donors, this would reflect reverse causality from violence to ART coverage. The inclusion of country fixed effects and country-level controls accounts for concerns related to systematic variation across countries and at the country level. Likewise, the inclusion of time fixed effects accounts for factors that affect social violence in a given year and that are common to all countries, which include the possible role of events that affect several countries at the same time and whose influence might vary over time (such as the Arab spring movements, or health initiatives by international donors or organizations). The inclusion of time trends for macro regions within Africa, and of country-specific trends related to HIV prevalence accounts for time-varying effects related to variation or interventions in specific macro areas (e.g., the increase in Islamic militant violence in Northern Africa during the early 2000s) and trends related to country-specific initial conditions in terms of HIV prevalence at the onset of the ART expansion.

#### Identification Strategy

To further address identification concerns related to omitted variables or reverse causality and to identify the effects of ART coverage on social violence, the analysis employs an identification strategy that is based on instrumental variables and 2SLS estimation. The instrument for ART coverage combines cross-sectional variation in the local scope for ART, *Z_c,_*_2001_, and the global time-series variation in the access to ART, *ART_IV,t_*. The resulting first stage regression model with instrument *Z_c,_*_2001_ *· ART_IV,t_* is

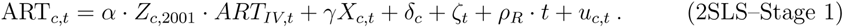

We use different measures in both dimensions to construct the instrumental variable as discussed in detail below.

In the intention-to-treat (ITT, or reduced form) analysis, social violence is regressed directly on the instrument. This analysis does not require a reliable geo-referenced and time-varying measure of ART coverage as instrumented variable and can thus also be conducted at the subnational region level, where such data are unavailable. For comparability, we conduct intention-to-treat estimations at the national and sub-national levels. The estimation framework is given by

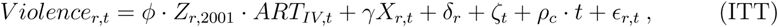

for data at the level of countries or sub-national, administrative regions *r*; the specification includes country-specific linear time trends *ρ_c_·t*. Throughout, the main effect (linear term) of the cross-sectional term of scope for ART, *Z_c,_*_2001_ or *Z_r,_*_2001_, is absorbed by the (country or region) fixed effects, *δ_r_*, and the time-varying instrument for ART coverage *ART_IV,t_* is absorbed by the year fixed effects *ζ_t_*.

#### Identification Assumptions

The major concerns for identification are related to unobserved third factors that correlate with both social violence and ART coverage (e.g., quality of institutions and governance), or to reverse causality, with the incidence of events of social violence influencing the access to ART in a given year and country or region (e.g., by political pressure on governments or international donors). The instrumentation addresses these concerns and allows identifying the effect of interest, *β* (or *φ*, respectively).

Technically, we construct the instrument using a measure for the cross-sectional differences in potential for ART *Z_c,_*_2001_ – capturing *where* ART coverage had a greater scope to increase upon availability. This is combined with a measure of the global expansion of ART treatment intensity *ART_IV,t_* – capturing *when* ART coverage increased. The validity of the instrument requires relevance (i.e., the instrument should be a relevant predictor of ART coverage) and the exclusion restriction: the instrument should affect social violence only through its effect on ART coverage. Concretely, this requires that the *interaction* between cross-sectional variation in potential for treatment in terms of HIV prevalence prior to the ART expansion (as of 2001), and global dynamics in the access to ART treatments, is exogenous to the incidence of social violence in a given year in a country or region, and hence that *Z_c,_*_2001_ *· ART_IV,t_* (or *Z_r,_*_2001_ *· ART_IV,t_*, respectively) is uncorrelated with *ε*.

The rationale for the instrumentation approach is based on a differences-in-differences logic. First, an increase in ART coverage is expected to have had more pronounced effects in countries or regions that exhibited a higher level of HIV prevalence at the time when ART became widely available. In the baseline implementation of the instrumentation strategy, the scope for ART coverage at the onset of roll out in HIV treatment, *Z_c,_*_2001_ is measured by the HIV prevalence in a country in 2001, i.e., prior to the world-wide ART expansion, *HIV_c,_*_2001_. Contrary to country-level data, HIV prevalence is not available for the same year (2001) in all regions; instead, *HIV_r_* measures the HIV prevalence in the respective region in the year closest to 2001 for which data are available. The details of the variable construction can be found in the Supplementary Appendix (Appendix Sections A.1.3). Before 2001, ART treatments were effectively not available in Africa, and hence the cross-country variation in potential for ART (HIV prevalence) is unrelated to potential confounds for the analysis that affect subsequent ART coverage, such as institutions, economic development, or political pressure, and that are absorbed by an extensive set of control variables. The direct effect of this local (country-specific or sub-national region-specific) variation in potential for ART expansion on violence is accounted for by (country or region) fixed effects, and time-varying effects of HIV prevalence before 2001 on violence are accounted for by the inclusion of control variables and flexible specifications of country-specific trends related to HIV prevalence.

Second, the worldwide expansion of ART availability, captured by *ART_IV,t_*, largely occurred for reasons unrelated to what happened within each African country. In particular, the time variation in global ART expansion is related to global factors like the decline in the price for medication and the resulting increase in availability of ART that were the result of international political agreements and innovation in the pharmaceutical industry and that were unrelated to region-specific or country-specific time trends in HIV prevalence or social violence. To implement a measure of global dynamics in ART availability and construct the instrument, we collected time series data of the prices for the most common first line of ART treatment regimens for adults and construct the instrument by interacting *Z_c,_*_2001_ with the median world price of ART treatment regimens as measure for ART treatment intensity *ART_IV,t_* (ART Price). This measure only provides indirect information about the availability of ART treatments in a country, but it has the advantage that the instrument does not respond, by construction, to the country-specific level of social violence or any policy intervention that is specifically targeted to a given country in a given year. Moreover, conceptually, this measure is directly related to actual ART treatment in a country since the reduction in prices constitutes the ultimate driver of the increase in ART coverage. The global dynamics were unrelated to the dynamics of social violence in particular African countries and the direct effects of global dynamics in ART availability are accounted for by time fixed effects and region-specific time trends.

The instrument combines these two dimensions, the potential for ART, *Z_c,_*_2001_, and global variation over time in the access to ART treatment responsible for the ART expansion, *ART_IV,t_*, to predict the country-specific expansion of ART coverage, and hence captures variation that is exogenous to the evolution of social violence in specific African countries. Controlling for country/region fixed effects and time-varying covariates accounts for systematic variation that might violate the exclusion restriction for the measures of cross-sectional heterogeneity in scope for ART. Moreover, the inclusion of year fixed effects, time-trends for African regions, or country-specific time trends accounts for trends in social violence that might violate the exogeneity of the global dynamics in ART expansion. Below, we present additional results for alternative constructions of the instrument. While conceptually capturing the same underlying phenomenon, the interactions of different cross-sectional measures of potential for ART treatment with different measures of the global expansion of ART treatment intensity differ in terms of data quality and potential concerns regarding the validity of the identifying assumptions of the corresponding interaction term that is used as instrument.

Our baseline instrumentation approach complements recent work by Acemoglu et al. (2020) that combines cross-sectional variation in mortality from several diseases prior to treatment and assumes that the respective mortality declined to zero in the context of the global epidemiological transition to investigate the effects of population dynamics on civil conflict (adapting a similar strategy from earlier work by Acemoglu and Johnson, 2007). In contrast, rather than using a proxy for the latent mortality decline based on the cross-sectional pre-treatment variation in mortality, our approach makes use of several alternative proxies for cross-sectional treatment scope in combination with several measures of time variation in treatment intensity that are based on global dynamics of the price or cost of treatment, or of the actual treatment intensity outside Africa. Our approach also differs from recent work on the economic effects of ART expansion by Tompsett (2020) who made use of time variation in ART coverage in low and middle income countries. In addition to our baseline instrumentation based on time variation in prices and cost of drugs, in the robustness checks below we also conduct the analysis using global variation in ART coverage in low and middle income countries outside Africa, thereby accounting for concerns of endogeneity due to cross-country spill-overs across African countries.

## 3 Results

### 3.1 Baseline Instrumentation

Table 1 presents the main results. All coefficients reported in this and other tables correspond to standardized regressors to ensure direct comparability. OLS regressions deliver a significantly negative association between ART coverage and the incidence of social violence at the country level (Table 1 Column 1). The 2SLS estimates reveal that the association remains significant and is quantitatively even larger. These results suggest an upward bias in the OLS estimates towards zero. This is consistent with possible problems of measurement error related to ART coverage, problems of simultaneity due to omitted factors that correlate positively with social violence and ART coverage, or with reverse causality due to social violence directly influencing HIV prevalence (McInnes, 2009; Iqbal and Zorn, 2010). One example of a confound are more foreign military or humanitarian interventions in countries or regions with a high incidence of social violence that are associated with better access to health provisions and, in particular, greater ART coverage. In fact, the evidence is consistent with such a confound. In particular, the data show a positive correlation between foreign interventions (measured in terms of the cumulative annual Global Fund disbursement) and social violence (events) of 0.23. At the same time, the correlation between foreign interventions and ART coverage is 0.49. Hence, failing to account for such foreign interventions might induce an upward bias in the OLS estimates. The use of an instrument that combines cross-sectional variation in the potential for ART treatment determined *before* the availability of ART, with time variation in the global expansion of the availability of ART *outside* Africa, provides exogenous variation that allows for a consistent estimation of the causal effect of ART treatment on social violence. By making use of the interaction between country-specific potential for ART treatment and the global increase in treatment intensity as instrumental variable, the instrumental variables approach therefore accounts for these identification concerns.

**Table 1:** Effect of ART Expansion on Social Violence

|  | EVENTS OF SOCIAL VIOLENCE |  |  |  |
| --- | --- | --- | --- | --- |
|  | OLS | 2SLS | ITT |  |
|  | (1) | (2) | (3) | (4) |
| ART | -0.316**<br>(0.157) | -0.966**<br>(0.362) |  |  |
| $Z_{i,2001} \times ART_{IV,t}$ | | | -0.163***<br>(0.060) | -0.166***<br>(0.034) |
| Instrument |  |  |  |  |
| $Z_{i,2001}$ | | $HIV_{c,2001}$ | $HIV_{c,2001}$ | $HIV_{r,2001}$ |
| $ART_{IV,t}$ | | ART Price | ART Price | ART Price |
| Observations | 1,394 | 1,394 | 1,394 | 4,760 |
| Clusters | 50 | 50 | 50 | 170 |
| Adj-R2 | 0.23 | 0.17 | 0.23 | 0.09 |
| Kleibergen-Paap |  | 33.77 |  |  |
| *Country f.e.* | ✓ | ✓ | ✓ | ✓ |
| *Year f.e.* | ✓ | ✓ | ✓ | ✓ |
| *Time Trend* | ✓ | ✓ | ✓ | ✓ |
| *HIV Trend* | ✓ | ✓ | ✓ | ✓ |
| *Population* | ✓ | ✓ | ✓ | ✓ |
| *Region f.e.* | × | × | × | ✓ |

Similar results are also found with an intention-to-treat approach at the country level or at the sub-national level (Table 1 Columns 3 and 4). Next, we present results for alternative constructions of the instrument that address various potential concerns.

### 3.2 Alternative Constructions of the Instrument

#### Alternative Measures of Global ART Expansion *ART_IV,t_*

Conceptually, the instrumentation approach combines cross-sectional variation in the scope for ART expansion, *Z_c,_*_2001_, with time variation in the dynamics of the expansion of ART treatment intensity *ART_IV,t_* as reflected by the global decline in the price of the most important treatment regimens. This variation in prices provides relevant information that is plausibly exogenous from the perspective of single countries in Africa. Nevertheless, this measure is subject to some potential limitations. One limitation for the quantitative interpretation of this instrument is that a reduction in the prices paid by a government might release budget resources that can be used for alternative policies and, accordingly, might lead to a reduction of, e.g., protests or strikes. Moreover, to the extent that prices for ART treatments might be determined by monopolistic pricing of pharmaceutical multi-national corporations, international organizations might exert an indirect effect through their influence on price negotiations. This would show up in terms of a decline in prices due to lower mark-ups over costs, raising concerns about simultaneity.

To address these concerns, we constructed an alternative measure for *ART_IV,t_* that is based on the evolution of the cost of the active pharmaceutical ingredients of the main first line ART treatments for adults (ART Cost). The conceptual advantage of this measure is that it captures the dynamics of global production costs and thereby directly reflects the decline in costs that was related to the increase in the amount of treatments produced worldwide. The reduction in costs of production also maps into the reduction in prices, but the time variation of the two series differs because of variation in the markups charged by pharmaceutical companies, particularly due to increasing competition associated with the introduction of active principles produced as generics. Thus, the validity of the instrument based on ART Cost is based on similar arguments as that based on ART Price, but this measure has the appealing feature of not relying on changes of markups of pharmaceutical companies, which might be influenced by political pressure. This makes the information about cost even less susceptible to pressures by international organizations since this variation is related to an increase in research competition and patent expiration. Hence, the use of ART Cost is conceptually preferable for instrument construction. However, this instrument is subject to more severe data limitations in terms of availability and coverage, which requires interpolation of data for years with missing information. This reduces the variability of the measure and leads to lower predictive power on the first stage.

As a second alternative measure for *ART_IV,t_*, we constructed the evolution of a synthetic price index of the main first line ART treatments for adults (Synth. Price). This measure is exclusively based on information about the initial price of first line treatment regimens of the first line of ART treatment in 2001, prior to the expansion of ART demand (and thus prior to potential influence of donors on the price development), and on price data after the major expansion (2015-2017). For the intermediate time period, the price index is constructed based on the assumption of a constant proportional decline of the price-over-cost mark-up each year. This decline approximates the global dynamics of treatment costs and prices in line with the sharp initial decline that is followed by more moderate reductions as market prices converge to the limit price. The appealing feature of this price index is that only two data points are involved in its construction, alleviating potential concerns about a demand-driven price decline that violates the exclusion restriction due to systematic variation in prices in response to an ART expansion in particular countries. The synthetic price (Synth. Price) index is therefore, by construction, unrelated to political interventions and to any other sort of demand-driven price decline while resembling the typical evolution of drug prices after the end of patent exclusivity, and after the introduction of generic drugs.

As a third alternative measure for *ART_IV,t_*, we make use of data on expansion of ART coverage in low and middle income countries outside Africa (ART Cov). The ART coverage in other low and middle income countries outside Africa captures effective variation in access, and thus comes closest conceptually to the variation captured by the instrumented variable (ART coverage in African countries), while offering high data quality and longer and more coherent time coverage compared to the data on the dynamics of prices and costs of treatment regimens. A similar identification strategy has previously been applied successfully at the country level to explore the economic effects of ART expansion (Tompsett, 2020), who made use of time variation in ART coverage in low and middle income countries. Differently from this application, the approach applied here combines cross-sectional heterogeneity in disease (HIV) prevalence prior to the treatment expansion in combination with time variation in ART coverage in low and middle income countries *outside* Africa. The use of global ART coverage outside Africa is conceptually not affected by the level of ART coverage in a specific country and the inclusion of year fixed effects accounts for global shocks. In particular, this addresses potential endogeneity through cross-country spill-overs across the African continent and ensures that the interaction is exogenous to social violence at the country-year level, conditional on the control variables in the empirical specification. However, the dynamics in global ART coverage might also be problematic for various reasons. In particular, a direct role of international actors or organizations in extending ART coverage worldwide might raise potential concerns about simultaneity. While global ART coverage is a proxy for lower ART prices, this variation might also correlate with the intensity of aid from the Global Fund and other donors, thus picking up dynamics that are linked to social violence, but that do not necessarily reflect the ART expansion due to cheaper drug provision. One might even consider the possibility of reverse causality if social violence reflects protests that led to a stronger response to HIV globally, such that higher ART coverage in low and middle income countries not only correlates with ART coverage in Africa but also with fewer protests by advocates of intensified HIV control.

In sum, while conceptually capturing the same variation, the three alternative dynamic instrument components differ in terms of data quality and potential concerns regarding the validity of the identifying assumptions, and thus provide useful alternatives to assess the sensitivity of the baseline instrumentation. Details of the construction of each of these variables can be found in the Supplementary Appendix (Sections A.1.5, A.1.6, and A.2).

#### Alternative Measure of Scope For ART Expansion (*Z_i,_*_2001_)

The baseline instrumentation uses HIV prevalence in the respective country or region in 2001, prior to the ART expansion, as measure of cross-sectional variation in the scope for ART expansion, *Z_c,_*_2001_. This instrumentation parallels recent work that has used disease prevalence prior to the epidemiological transition (Acemoglu et al., 2020). To address potential concerns about the exogeneity of the cross-sectional variation in the scope for ART expansion, *Z_c,_*_2001_, we conduct extensive robustness checks, including tests of parallel trends, placebos, different base years, or additional controls and interaction terms, which are reported below. The most salient concern about the use of cross-sectional variation in HIV prevalence in 2001 is a potential correlation with institutions and other factors that would otherwise question the exclusion restrictions. The extensive specifications with country fixed effects and additional controls, and recent findings that interaction terms with one potentially endogenous factor require weaker identification assumptions than standard exclusion restrictions (Bun and Harrison, 2019), alleviate this concern.

To further account for the concern that persistent factors such as institutions or culture might affect pre-ART-expansion HIV prevalence as well as post-ART-expansion social violence and ART coverage jointly, thus posing a threat to the exclusion restriction, we constructed an alternative proxy measure of the country-specific scope of ART expansion that exclusively relies on geography, *HIV_geo_*. Concretely, we compute the geography-related exposure to HIV as the effective distance from Kinshasa, using exclusively information about first nature geographic characteristics and a minimum criterion for population, based on the fast-marching method (Sethian, 1996, 1999). The resulting proxy variable therefore reflects the effective distance to the origin of the HIV epidemic that measures the potential exposure to HIV. This measure is therefore a valid predictor of the crosssectional distribution of HIV prevalence in Africa during the late 20th century prior to the ART expansion. By construction, this measure is not driven by institutional, cultural or political features that could have affected the evolution of the HIV epidemic in a country prior to 2003 and that might challenge the exclusion restrictions. The details of the variable construction can be found in the Supplementary Appendix (Appendix Section A.1.4).

#### Results for Alternative Instrument Constructions

Table 2 presents results of 2SLS regressions with instruments constructed from these different measures of *Z_i,_*_2001_ and *ART_IV,t_* in comparison to the results for the baseline measures that are presented in Column (1). The remaining columns show results for different combinations of measures to construct the instrument *Z_i,_*_2001_ *× ART_IV,t_*. The first stage results indicate that all instruments are relevant, with somewhat weaker but still acceptable performance of instruments based on the geography-based measure HIV*_geo,_*_16_*_K_*.^2^ The comparison of second stage estimates from the different specifications of the instrumental variable suggest that the results are not sensitive to the different instrument constructions. This provides strong support for the validity of the main findings and their robustness to potential confounds discussed before.

**Table 2:** Effect of ART Expansion on Social Violence – Alternative Instrumentation

| SOCIAL VIOLENCE (LOG EVENTS) - SCAD DATA |  |  |  |  |  |  |
| --- | --- | --- | --- | --- | --- | --- |
|  | 2SLS | 2SLS - ALTERNATIVE IV CONSTRUCTIONS |  |  |  |  |
|  | (1) | (2) | (3) | (4) | (5) | (6) |
| ART | -0.966**<br>(0.362) | -0.954**<br>(0.361) | -0.922**<br>(0.356) | -0.999**<br>(0.388) | -1.197**<br>(0.578) | -1.227**<br>(0.575) |
| Instrument |  |  |  |  |  |  |
| $Z_{i,2001}$ | $HIV_{c,2001}$ | $HIV_{c,2001}$ | $HIV_{c,2001}$ | $HIV_{c,2001}$ | $HIV_{geo,16K}$ | $HIV_{geo,16K}$ |
| $ART_{IV,t}$ | ART Price | ART Cost | ART Synth. Price | ART Cov | ART Price | ART Synth. Price |
| Observations | 1,394 | 1,394 | 1,394 | 1,394 | 1,366 | 1,366 |
| Clusters | 50 | 50 | 50 | 50 | 49 | 49 |
| Adj-R2 | 0.17 | 0.18 | 0.18 | 0.17 | 0.08 | 0.07 |
| Kleibergen-Paap | 33.77 | 31.42 | 38.12 | 34.69 | 11.40 | 11.14 |
| *Country f.e.* | ✓ | ✓ | ✓ | ✓ | ✓ | ✓ |
| *Year f.e.* | ✓ | ✓ | ✓ | ✓ | ✓ | ✓ |
| *Time Trend* | ✓ | ✓ | ✓ | ✓ | ✓ | ✓ |
| *HIV Trend* | ✓ | ✓ | ✓ | ✓ | ✓ | ✓ |
| *Population* | ✓ | ✓ | ✓ | ✓ | ✓ | ✓ |

The results of statistical tests on instrument selection reveal that the joint validity of all instruments is never rejected while none of the instruments is redundant in the sense that asymptotic efficiency of the estimation is improved by each instrument. Since the instruments are highly correlated, the null of orthogonality is rejected for each combination of instruments, suggesting the use of a single instrument at a time, rather than a combination of instruments. More details on identification are contained in the Appendix Section A.2.

### 3.3 Synthetic Control Approach

Figure 3 provides an illustration of the impact of ART on social violence based on the synthetic control method for causal inference in comparative case studies (Abadie and Gardeazabal, 2003; Abadie et al., 2010, 2015; Abadie, 2021). The synthetic control method is an alternative data driven procedure to construct a time-varying counterfactual unit for each treated unit by creating a weighted combination of different control units. Unlike in a difference-in-difference approach, the synthetic control method thereby allows controlling for time-varying confounders and provides a systematic data-driven way of constructing a synthetic counterfactual for treated units had they not received the treatment. In the figure, treated units correspond to the 25% of countries with the highest levels of ART coverage (relative to HIV infected individuals) after 2001.^3^ The donor pool of countries that are used to construct the counterfactual for each treated unit consists of the 25% of countries with lowest ART coverage after 2001. Alternatively, the analysis was replicated with the bottom 10% of countries in terms of average ART coverage after 2001 as donor pool. The graph illustrates that the trend in social violence is almost identical in the treatment and synthetic control groups before the onset of the ART expansion. After the onset, the treatment group experienced substantially lower levels of social violence than the control group.

**Figure 3:**
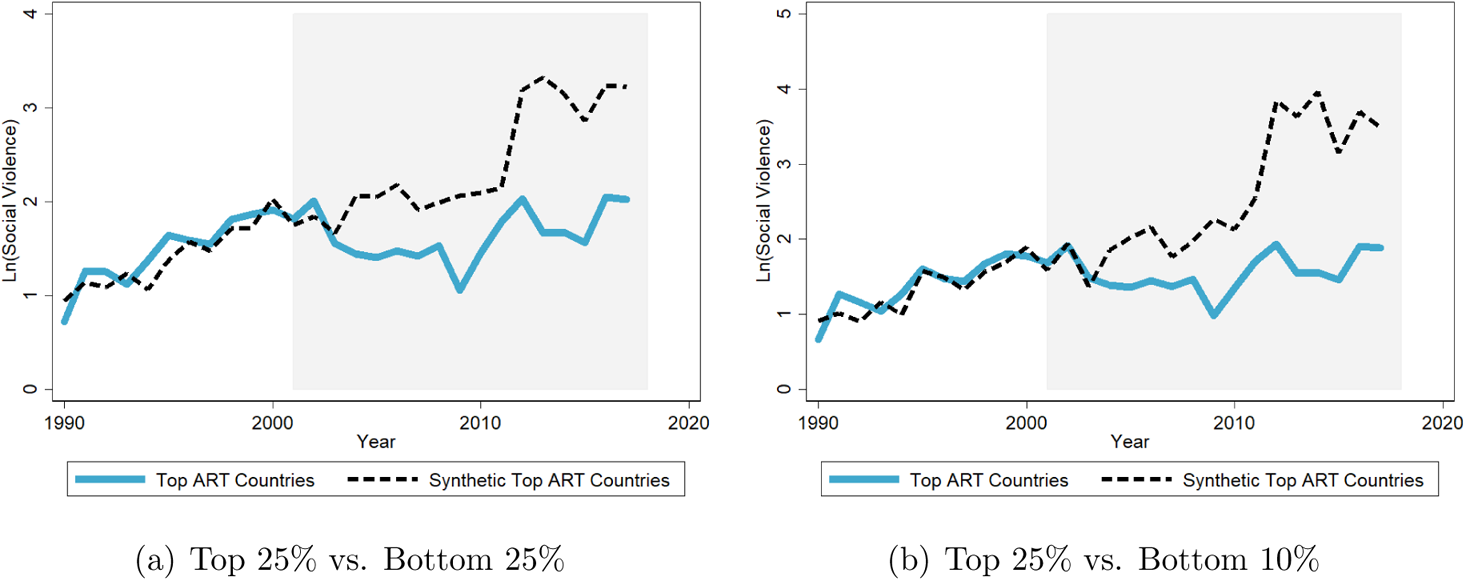
ART Expansion and Social Violence: Synthetic Control Approach *Note:* Results based on the synthetic control method. For each treated unit, the incidence of social violence is computed under the average treatment and for the synthetic counterfactual. The graph plots averages across all treated units. With the intervention period beginning in 2001, the synthetic control is computed for each treated unit by minimizing the mean squared prediction error (MSPE) relative to the treated units during the pre-intervention period 1990 to 2000. As predictor variables for the construction of the weighted counterfactual of each treated unit, the procedure uses the average log number of conflict events, population and HIV prevalence (all measured between 1990 to 2000), the fraction of the country area within 100 km from the coast, the fraction of desert and of tropical forest, latitude and longitude.

### 3.4 Robustness

In this subsection, we briefly describe the results of various robustness checks. The detailed results are contained in the Supplementary Appendix.

#### Parallel Trends Assumption and Alternative Base Years

Regarding the plausibility of the parallel trends assumption, i.e., a similar dynamic evolution of social violence in the absence of differential ART coverage, neither the raw data nor group-year averages reveal any evidence for systematic trend differences in social violence across countries with different HIV prevalence in 2001 (Appendix Figures A5 and A6). To test the sensitivity of the results with respect to the choice of 2001 as base year for HIV prevalence to measure the scope of ART expansion, the estimation was repeated with alternative base years with similar results (Tables A5 and A6, Figure A7).

As a more formal way to assess the plausibility of the parallel trend assumption, we constructed event-study graphs based on estimators that are robust to heterogeneous treatment effects, across units or over time. In particular, the graphs map the reduced form effect of the instrumental variable on social violence, while the inclusion of lags and leads allows displaying the dynamics of the effect over time. The estimation is performed using the routine devised by Chaisemartin and Haultfoeuille (2020). In terms of controls, each estimation replicates an analogous specification as in Table 1 Column (3). In addition to the instantaneous effect, each estimation is performed including 8 placebo effects to assess the effect dynamics and the plausibility of parallel trends assumption. The specification also allows for 10 dynamic effects to explore the evolution of the effect over time. The results are shown in Figure 4 with the omitted coefficient reported as zero. The results show consistently insignificant effects of the pre-trends, which is reassuring regarding the plausibility of the parallel trends assumption. Concretely, countries that are about to experience a more pronounced increase in ART coverage, as proxied by the instrument, are not experiencing any different decrease/increase in social violence in the years preceding the increase in ART coverage. In terms of timing of the effect, the graph indicates that the effect materializes right on impact and then increases monotonically over time in terms of magnitude. Similar patterns emerge for alternative instrument constructions (see Appendix Figure A8).

**Figure 4:**
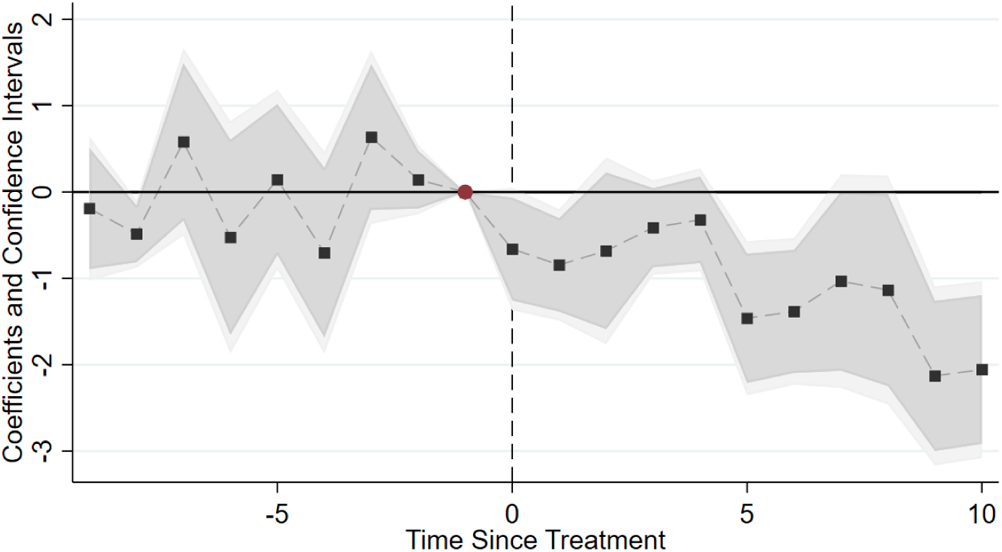
Reduced Form Estimates – Event Study Plots Note: The figure plots event study graphs for the coefficient of interest from the ITT model. The empirical specification is as in Table 1 Column (3) of the paper, using *HIV_c,_*_2001_*×*ART Price as instrument. The estimation is conducted using the routine devised by Chaisemartin and Haultfoeuille (2020). Dark shades show the corresponding 90-% confidence interval, light shades the corresponding 95% confidence interval.

#### Price/Cost Data for Alternative Treatment Regimens

The results are not sensitive to the use of price and cost information for a specific first line treatment regimen. In particular, the results are similar when using the price for an alternative, first line regimen (Table A7). This suggests that the findings are not driven by the specific ART treatment used to construct the instruments. A similar comment applies when alternative time-varying measures of ART expansion are combined with the alternative measures of cross-country variation in the scope for expansion (Table A8).

#### Falsification: Malaria Prevalence and Malaria Treatment

To investigate the validity of the instrument and the exclusion restriction, a falsification exercise was conducted by exploiting information about the prevalence of malaria and about the time evolution of prices for anti-malaria treatment. Malaria is a disease of major importance in Africa that has been subject to extensive health campaigns promoted by governments and international organizations. Moreover, weather-driven malaria shocks have been documented to lead to outbreaks of social violence (Cervellati et al., 2021). Similar to HIV treatment, the treatment of malaria has attracted funds from international organizations and global efforts have led to substantial reductions in prices for treatments. The validity of the identification strategy implies that alternative instruments that combine scope for HIV treatment with the price of malaria treatment, or variation in global access to ART treatment with scope for malaria treatment should not predict ART coverage. The results of these falsification tests show that a combination of information about malaria prevalence with time variation in the expansion of ART coverage, or of pre-expansion HIV prevalence with variation in world prices of anti-malaria treatments, do not predict ART coverage (Appendix Table A9).

#### Placebo and Overidentification: Heterogeneity in Institutional Quality

As alternative test of the validity of the instrument and the exclusion restriction, the estimation was conducted when using various measures of institutional quality as cross-sectional component of the instrumentation stage, replacing the scope for treatment, *Z_c,_*_2001_, in the instrument *Z_c,_*_2001_*·ART_IV,t_* on the first stage of the 2SLS framework (2SLS–Stage 1) by the institutional placebo *X_c,_*_2001_ and estimating the model with instrument *X_c,_*_2001_ *· ART_IV,t_* (Tables A10 and A11). Alternatively, an extended version of model (2SLS–Stage 1) was estimated using *X_c,_*_2001_ *· ART_IV,t_* as additional control (Tables A12 and A13). Both sets of robustness checks provide no indication of a violation of the identification assumptions and confirm the main results.

#### Relaxing the Assumption of Strict Exogeneity of the Instrument

To investigate the sensitivity of the results with respect to violations of the exclusion restriction of strict exogeneity, we estimated extended specifications that relax the restriction that the direct effect of the instrument on the outcome is exactly equal to zero (Conley et al., 2012). These estimates reveal that, in order for the effect of interest to be not statistically different from zero, the direct effect, conditional on all controls, would have to be almost of the order of magnitude as the effect of interest, which we consider implausible (Figure A9).

#### Sub-samples

The results are not sensitive to the inclusion/exclusion of single countries (Figures A10 and A11). Similar findings emerge when accounting for separate, non-linear time trends across African regions (Table A14). Separate estimates for different sub-samples of countries suggest that the instruments are stronger for the sub-samples of sub-Sahara Africa, or the sample of countries with high HIV prevalence within Africa, but the overall results are similar to the baseline (Table A15).

#### Confounds Related to International Aid, Institutions, and Economic Development

The potential role of specific time-varying country-specific characteristics that could drive both health policies and social violence has been discussed in the context of the differences between OLS and 2SLS results. We explored this aspect in various dimensions. Results from extended specifications with controls for the level of health aid (measured by the extent of Global Fund donations received by a country in a given year), for the level of development (in terms of GDP per capita), or for the quality of public governance (in terms of democracy), confirm the baseline results (Table A16). Moreover, instrument performance is not affected by including these controls, which provides support for the notion that the identifying variation contained in the instruments is exogenous and not driven by these time-varying country-specific features.

#### Scale

Similar results were found when using ART coverage of the population instead of relative to the population living with HIV in a country (Table A17). Likewise, the finding that the expansion of ART coverage led to a reduction in social violence consistently emerges for when considering social violence relative to the population as measured by the log events per population (Table A18).

#### Confounds Related to International Organizations or Interest Groups

The discussion of the identification assumptions suggests a potential violation of the exclusion restriction as the result of interventions at the international level that led to changes in ART coverage or prices. To explore this possibility, text analysis was applied to the narratives of SCAD events to assess if the results are driven by specific event types or events involving particular groups of participants. This analysis reveals that the results are not driven by events involving NGOs or health workers, public employees or strikes (Tables A19 and A20). This suggests that the main findings are not driven by events that could be connected to political pressure on governments or international organizations or, for instance, the use of budget resources freed by the reduction in prices under the pressure of specific categories of actors or events.

### 3.5 Mechanisms

The results presented so far provide evidence that is consistent with the hypothesis that health interventions, such as the ART expansion, led to a reduction in social violence. In the following, we present additional results on the potential mechanisms behind these findings.

#### Health Improvements vs. Generic Improvements in Economic Prosperity

We begin the analysis by investigating whether the negative effect of improved public health on social violence and unrest might merely be related to generic improvements in economic prosperity that have been documented in the context of the ART expansion (see Tompsett, 2020).^4^

In an attempt to investigate whether the effect could be mediated by generic improvements in economic well-being, we conducted an additional analysis in which we replaced ART treatment as instrumented variable by income per capita (or income per capita growth) or life expectancy, and estimated the effect of the ART expansion on social violence that works through income or health. The results reveal that the ART expansion, as instrumented by the interaction of the cross-sectional scope for expansion and the global dynamics of the ART expansion, is only a weak predictor of GDP per capita, with a weak first stage performance (with F-statistics around 1). In the second stage, the estimates provide no evidence that the effect of the ART expansion on social violence might work through income (see Appendix Tables A24–A27). Repeating the same analysis for life expectancy as mediating factor in fact reveals strong and statistically highly significant effects on the first stage. In the second stage, the results show a significant reduction in social violence as life expectancy increased in the context of the ART expansion, but this effect only materializes for social violence as measured in the SCAD data, not for major armed conflicts as measured in the UCDP data (see Appendix Tables A28 and A29). This evidence corroborates the previous findings and suggests that health interventions play an important independent role in reducing social violence that go beyond variation in economic living conditions. In sum, these findings support the hypothesis that health interventions exhibit a “dividend” that is distinct from the direct effects on health and economic well-being by reducing social violence.

#### Types of Violent Events

In a next step, we conduct an additional analysis on the different types of violent events and replicate the analysis for data on social violence and civil conflict from various different sources.

Figure 5 plots the standardized IV estimates of the coefficient of interest for the baseline specification and for different types of social violence and the different instrument constructions (using ART Price, ART Cost, and ART Cov) at the country level. The significantly negative effect of the ART expansion emerges for the data on social violence from the Social Conflict Analysis Database (SCAD) as well as for data from the Armed Conflict Location and Event Data (ACLED) or data from the Global Database of Events, Language, and Tone (GDELT). Similar to SCAD, both alternative data sources contain information about frequent and recurrent events of social violence, in terms of protests and riots, and deliver comparable estimates (see also for detailed results for ACLED data base, Tables A30–A31; and for GDELT data, Table A32). In contrast, we find no significant effect of ART expansion on civil conflict, as measured by the Uppsala Conflict Data Program (UCDP), consistent with the conjecture that health interventions mainly affect social violence (Table A33). Additional results for different subsets of conflicts without casualties versus with casualties, or for different numbers of participants, confirm this view (see Table A34). To rule out the possibility that social violence and ART coverage could be related to ongoing major civil conflicts, we also estimated an extended specification with social violence based on the SCAD data as dependent variable, while including ongoing major armed conflicts (based on the UCDP data) as additional control. The results are basically unaffected, again highlighting that health interventions mainly affect social conflict (Table A35).

**Figure 5:**
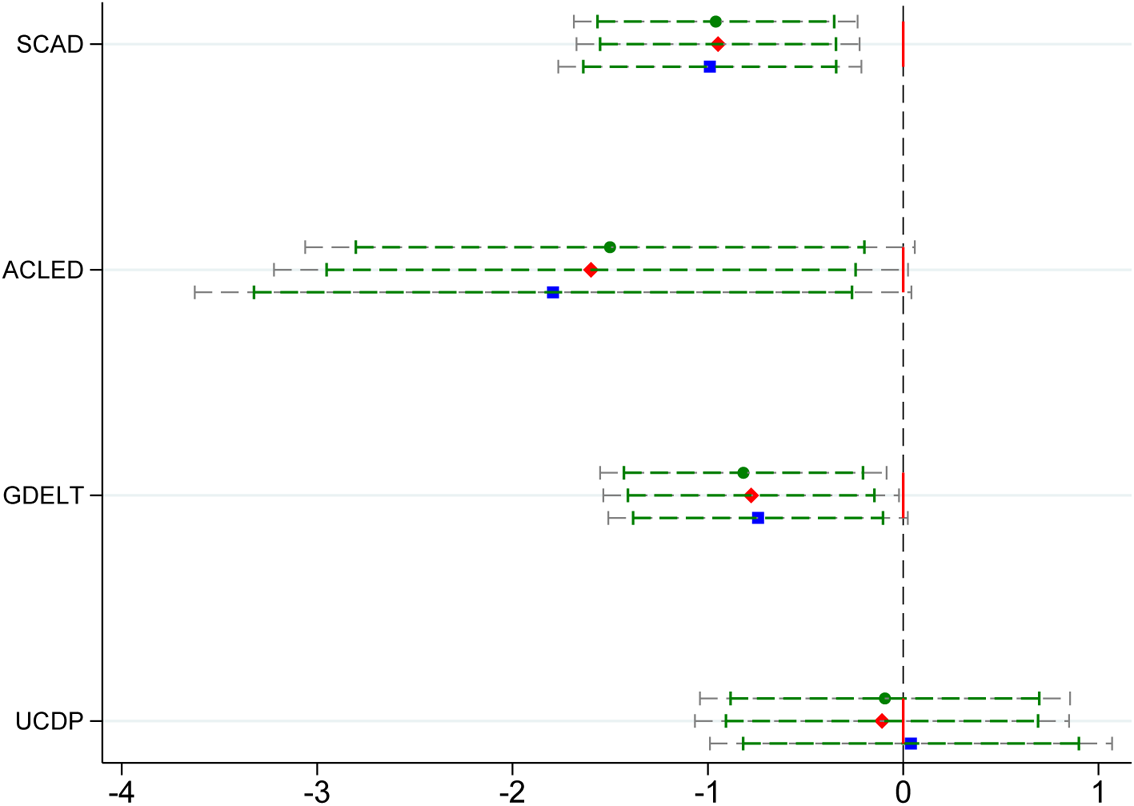
Mechanisms: Types of Social Violence *Note:* 2SLS estimates of *β*, country level (see Section 2.4). Instrument: interaction between cross-sectional variation in the potential for ART treatment, *Z_i,_*_2001_ (measured by HIV prevalence at country level, 2001), and a time-varying measure of ART expansion, *ART_IV,t_* (measured by the global variation in the median world price of ART treatment regimens, ART Price, or, alternatively, by the cost of ART treatment regimens, ART Cost, or global ART coverage outside Africa, ART Cov); the interaction term has been standardized. Coefficients are based on the same specification as in Table 1 Column (2). Dependent variable is log events of social violence from the different data sets (SCAD, ACLED, GDELT, UCDP, see text for details).

#### Motives for Social Violence

To investigate the underlying mechanisms in more detail, we replicate the estimation at the sub-national level using events of different types and with different underlying motives, based on categories provided by the SCAD database, as dependent variable. We conduct the analysis for subsets of violent events as dependent variables. Violent events are classified based on the information about types of events (all, spontaneous, organized) or motives (events related to elections, economic factors, human rights) in the SCAD data set. Intention-to-Treat regressions are based on the (log) number of events in a particular category as outcome variable (see Appendix Sections A.1.1 and A.4.8.1 for details).

Figure 6 reports the respective intention-to-treat estimates of the coefficient of interest at the sub-national region level for the different instrument constructions (using ART Price, ART Cost, and ART Cov). The results document that the reduction in violent events is particularly pronounced for organized events of social violence. This is consistent with fewer demonstrations, strikes, or other forms of social discontent as result of better provision with ART. In contrast, no significant effect is found for spontaneous outbreaks of social violence. In terms of motives behind social violence, we find that better treatment coverage reduces social violence related to economic factors and human rights, but we find no significant reduction in violence associated with political or electoral reasons (see also Appendix Table A36).

**Figure 6:**
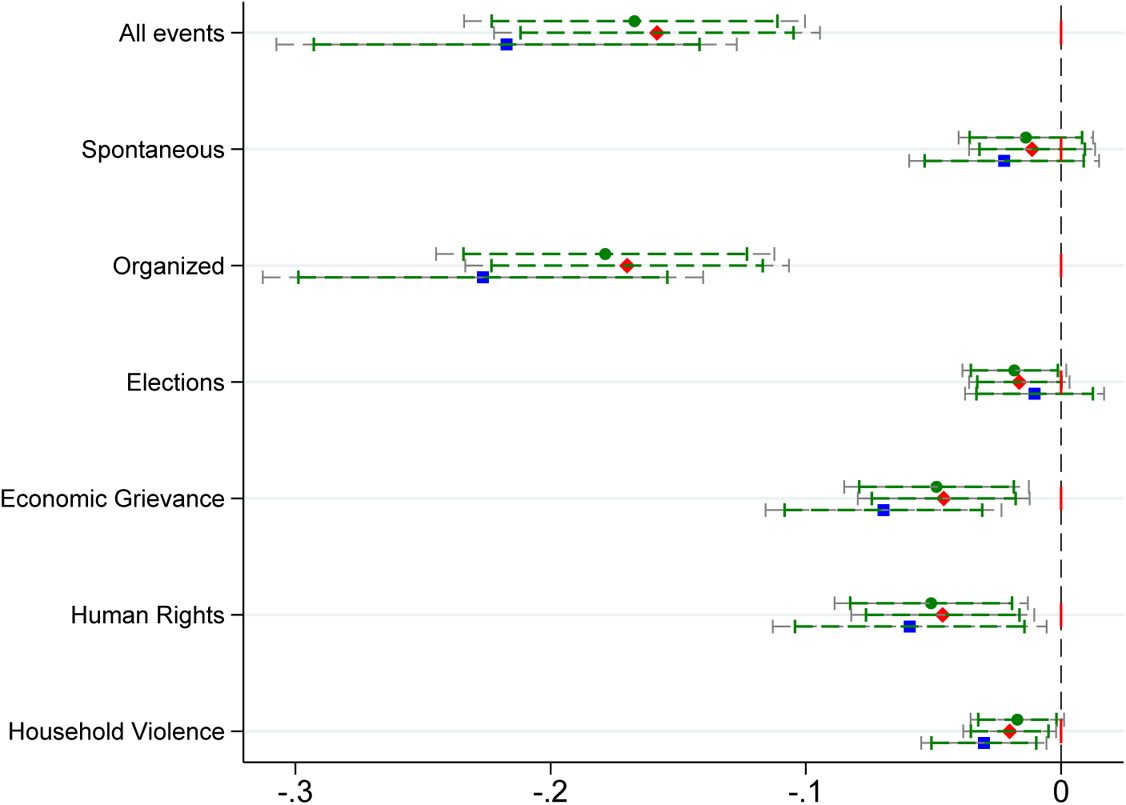
Mechanisms: Motives for Social Violence *Note:* Intention-to-treat estimates of *φ*, regional level (see Section 2.4). Instrument: interaction between cross-sectional variation in the potential for ART treatment, *Z_i,_*_2001_ (measured by HIV prevalence at region level, 2001), and a time-varying measure of ART expansion, *ART_IV,t_* (measured by the global variation in the median world price of ART treatment regimens, ART Price, or, alternatively, by the cost of ART treatment regimens, ART Cost, or global ART coverage outside Africa, ART Cov); the interaction term has been standardized. Coefficients are based on the same specification as in Table 1 Column (4). Dependent variable is log events of social violence, classified by motives; classification of social violent events is based on the codification in the SCAD database. Household Violence: intimate partner violence; Source: Demographics and Health Surveys (DHS), see also text and Appendix A.4.7.6.

To explore the effects on other forms of violence experienced by individuals at the household level, we collected data for intimate partner violence from the Demographics and Health Surveys (DHS) and replicated the analysis for this dependent variable. Despite serious data limitations in terms of spatial and temporal coverage, we find patterns for violence at the household level that are qualitatively comparable to the findings for social violence (bottom row of Figure 6 and Appendix Figure A13). The fact that the results generalize to different coding of violent events is interesting as it provides an indication that the results are not limited to a particular type of social violence. At the same time, the results again indicate that health interventions mainly reduce social violence events, which might reflect a reduction in discontent with the government or the (health-related) living conditions, but not major armed conflict or civil wars. This is also consistent with the findings from the analysis of potential confounds related to international interventions or interest groups, which revealed that the effects mainly emerge for social violence that does not involve organized actors like strikes, NGOs, or public health workers or civil servants (Tables A19 and A20). Moreover, the consistency of the results is reassuring regarding the validity of the instruments because the exclusion restrictions are conceptually different and less likely to be violated in light of the discussion above.

#### Approval of Government Policy

The results shown so far point towards a greater approval of governmental policies as potential mechanism behind the reduction in social violence. In particular, the successful expansion ART led to improved health and, indirectly, an alleviation of concerns about overall economic and political living conditions, which might have led to a decline in, e.g., organized social unrest like protests and riots. To investigate the empirical validity of this conjecture, we conducted additional analysis based on survey data from the Afrobarometer. In particular, we consider responses to questions that relate to the individual approval of policy, as dependent variables. Survey questions about individual approval of government policies range from individual assessments of how well the government handles HIV/AIDS, to provision with basic health, management of the economy, or combatting crime (see Appendix Section A.1.7.2 and A.4.8.2 for details).

Figure 7 displays the intention-to-treat estimates of the coefficient of interest corresponding to the survey responses to the subjective approval of government policies in various dimensions. A higher ART coverage is associated with a significantly higher approval rate of the government’s management of HIV, whereas the approval of the government’s management of education is unaffected by the ART expansion. This suggests that the decline in violence is associated with a more positive assessment of the government’s actions in the dimension of dealing with HIV. This also has implications for a greater approval of government policies in the domain of basic health provision, the economic domain, and, to a lesser extent, with a reduction of crime (see also Appendix Table A37).

**Figure 7:**
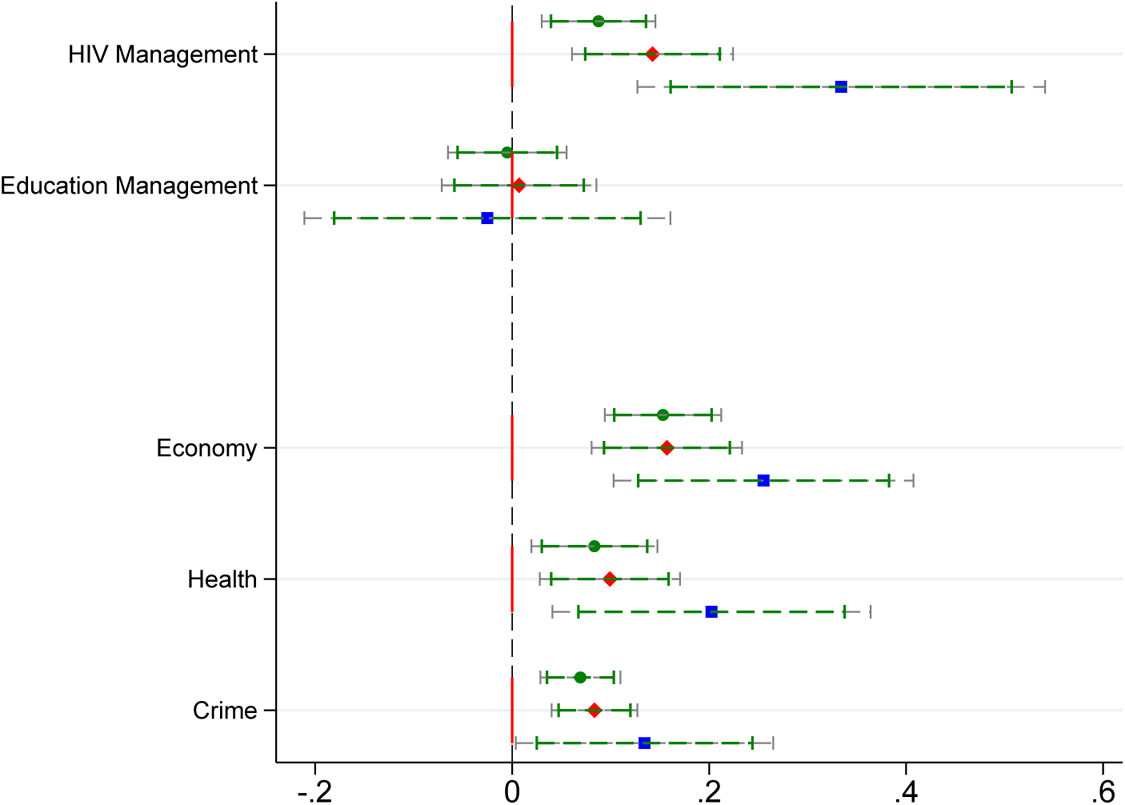
Mechanisms: Approval of Government Policy *Note:* Intention-to-treat estimates of *φ*, regional level (see Section 2.4). Instrument: interaction between cross-sectional variation in the potential for ART treatment, *Z_i,_*_2001_ (measured by HIV prevalence at region level, 2001), and a time-varying measure of ART expansion, *ART_IV,t_* (measured by the global variation in the median world price of ART treatment regimens, ART Price, or, alternatively, by the cost of ART treatment regimens, ART Cost, or global ART coverage outside Africa, ART Cov); the interaction term has been standardized. Coefficients are based on the same specification as in Table 1 Column (4). Dependent variables: survey responses to questions how well the current government handles various policy issues (HIV/AIDS, basic health provision, the economy, and crime). Data are from Afrobarometer. Confidence at 90 % (green) and 95% (grey). Corresponding estimates at sub-national region level are reported in Table A37.

#### Trust in Institutions

Finally, better perception of government policies dealing with HIV might also contribute to a greater trust of individuals in institutions and policy makers. The critical role of trust in institutions for building peaceful and inclusive societies, and the role of satisfaction with government policies, has been emphasized repeatedly by international organizations (see, e.g., OECD, 2017; United Nations, 2021), but direct evidence for how specific policies, in particular health policies, contribute to strengthening trust in institutions is still lacking.

To shed light on greater trust in institutions as part of the potential underlying channel of transmission we conducted additional analysis based on survey responses to questions that relate to trust in various dimensions as dependent variables. Again, survey questions are from the Afrobarometer and refer to trust in specific institutions (the parliament, the local government, the police); survey questions about approval of government policies range from individual assessments of how well the government handles HIV/AIDS, basic health, the economy, or crime (see Appendix Sections A.1.7.2 and A.4.8.2 for details).

Figure 8 displays the intention-to-treat estimates of the coefficient of interest corresponding to the survey responses about trust in institutions. The findings reveal that the ART expansion, as proxied by the instrument for a greater ART coverage at the sub-national level, is associated with greater trust in the national parliament as well as in the local government. In contrast, no significant effect is found for trust in institutions that are related to implementing law and order (represented by the police), rather than policy making in relation to HIV. This suggests that health interventions do not generically increase the trust in institutions, but trust in specific institutions and actors that individual respondents associate with the successful implementation of these interventions (see also Appendix Table A38).^5^

**Figure 8:**
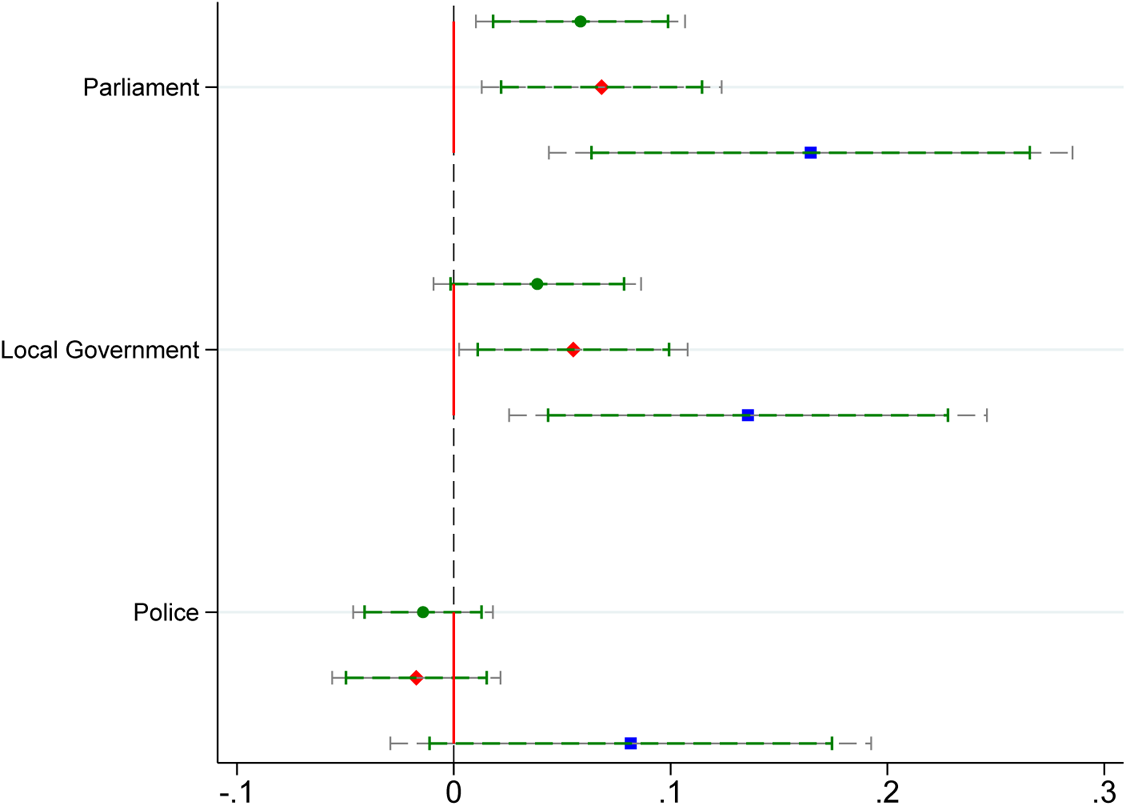
Mechanisms: Trust in Institutions *Note:* Intention-to-treat estimates of *φ*, regional level (see Section 2.4). Instrument: interaction between cross-sectional variation in the potential for ART treatment, *Z_i,_*_2001_ (measured by HIV prevalence at region level, 2001), and a time-varying measure of ART expansion, *ART_IV,t_* (measured by the global variation in the median world price of ART treatment regimens, ART Price, or, alternatively, by the cost of ART treatment regimens, ART Cost, or global ART coverage outside Africa, ART Cov); the interaction term has been standardized. Coefficients are based on the same specification as in Table 1 Column (4). Dependent variables: survey responses to questions about trust in institutions (parliament, local government, police). Data are from Afrobarometer. Confidence at 90 % (green) and 95% (grey). Corresponding estimates at sub-national region level are reported in Table A38.

Together, these results indicate that once the root cause of hardship is alleviated by effective treatment of the disease, here in terms of the management of HIV by increasing ART coverage, approval with governmental policies that lead to improved health and economic conditions is increased. Consistent with the conceptual considerations, this greater approval of the governmental responses and policies is associated with higher trust in institutions that are seen as responsible for the policies, and goes along with a decline in social violence and unrest.

## 4 Discussion

By documenting that the expansion of ART coverage in the context of the HIV/AIDS epidemic in Africa led to a significant reduction in social violence, our findings extend earlier evidence regarding the consequences of the ART expansion for increased life expectancy, incomes, and quality of life in various dimensions. From a policy perspective, the findings suggest another channel through which health interventions have the potential to break the vicious cycle of poor living conditions, short-sighted behavior, and lack of development: health interventions can play an important role in reducing social tensions and unrest. Considering the impact of health policies over a time horizon in which population responses can be expected to be minor, we find that public health improvements lead to a significant decline in social violence, but have no effect on large-scale civil conflicts. Quantitatively, the 2SLS results of Table 1 suggest that a 10% increase in ART coverage implies a reduction in events of social violence of about 25% of the unconditional mean. As the potential for treatment depends on HIV prevalence, the interpretation of an average effect of ART coverage is not straightforward.

To provide an illustration of the quantitative relevance of the results, we conducted a simulation exercise that contrasts, for each country, the observed average number of violent events with the number of violent events predicted by the model estimates. The predicted effect of ART treatment is evaluated relative to a simulated counterfactual scenario in which each country is assigned an ART coverage equivalent to the average of the 10% of countries with the highest ART coverage during a given year. With the exception of ART coverage, all other influences are evaluated at the respective means of the explanatory variables, in a given year.

Figure 9 contrasts a country’s actual average HIV prevalence in 2001 and average ART coverage with the predicted reduction in social violence that the country would have experienced as a result of the counterfactual treatment.^6^ By combining the model estimates with actual ART coverage in the countries with the highest coverage, this provides an indication of the predicted effects of conceptually feasible policy interventions. The results indicate that an extended ART coverage comparable to that of the 10% countries with highest ART coverage would have led to a reduction in violent events of around 5 percent on average. The benefits from intensified treatments in terms of reductions in social tensions would have been larger for some countries with intermediate or below-average HIV prevalence and treatments like South Sudan, Guinea-Bissau or Nigeria, or for countries that implemented the treatment with delay, like South Africa.^7^

**Figure 9:**
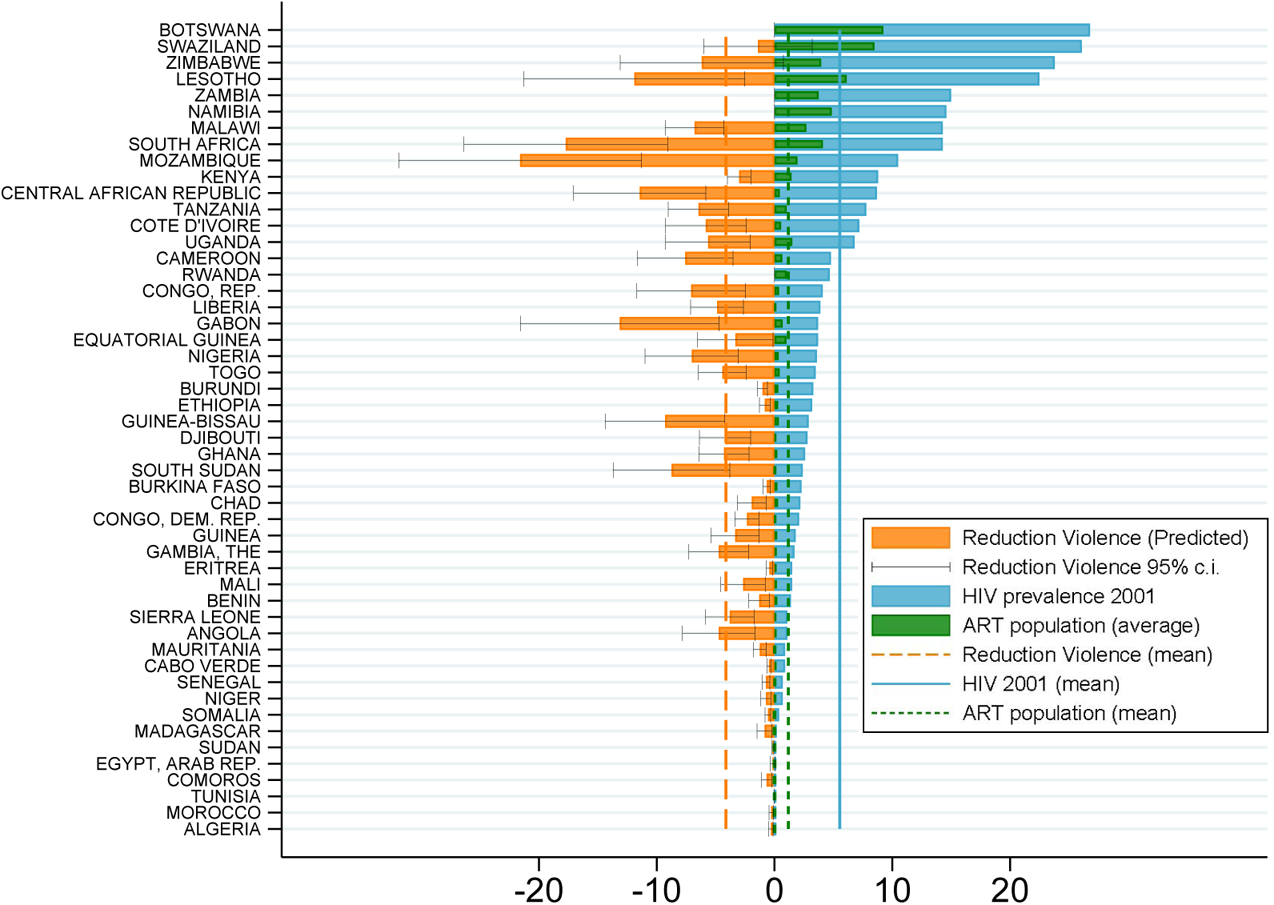
Quantification and Policy Implications: Counterfactual Predictions *Note:* Reduction in Social Violence: Counterfactual % reduction in violent events between 2000 and 2017 based on a simulation of country-specific ART coverage set to the level of ART coverage of the 10% countries with the highest ART coverage in a given year. Simulation based on same specification as baseline model estimates (Table 1(2), see Appendix Table A17), changes relative to observed values. Effects for countries with ART coverage in the top 10% are set to 0. HIV prevalence 2001: observed prevalence, used as proxy for ART potential (see text for details). ART Coverage: Observed coverage by country (relative to population, average 2000–2017). Vertical lines indicate the average of each respective variable (orange: predicted reduction in violent events; green: average ART treatments 2000-2017; blue: HIV prevalence in 2001).

Our results call for sustained efforts to fight HIV and other infectious diseases by showing that the predicted benefit from intensified ART provision is relatively larger in countries with below-average or intermediate disease (HIV) prevalence and where disease control has been given relatively lower priority. Evidence that the ART expansion reduced social violence through improved trust in institutions corroborates suggestions that health interventions can foster trust in states and policies and contributes a new perspective on the mixed evidence for the role of policy interventions to curb conflict. By improving individual health, promoting labor productivity, and attenuating social tensions, health interventions have the potential to generate a “triple dividend”. In light of these findings, further research extending beyond ART treatment, and tackling other current challenges, such as the unequal global access to COVID-19 vaccination is warranted to explore the external validity of the effects of public health interventions on social violence documented here. Another promising direction for future research is to further explore the role of health policies for building trust in institutions.

## Data Availability

All data used in the manuscript are publicly available online. A replication dataset will be made available after publication in a journal.

## A.1 Data

### A.1.1 Data: Measuring Social Violence

#### A.1.1.1 Social Conflict Analysis Database - SCAD

##### Primary Sources and Data Construction

The main measure of events of social violence is based on information from the Social Conflict Analysis Database –SCAD (Salehyan, Hendrix, Hamner, Case, Linebarger, Stull and Williams 2012). These data contain information on protests, demonstrations, riots, strikes, and other forms of social disturbances in Africa. The data set does not include violent events related to organized armed conflict such as rebellions, civil wars, and international war. The SCAD data and documentation about coding procedures can be accessed at https://www.strausscenter.org/scad.html (last accessed 3.8.2020).

The SCAD database covers all countries with a population of more than 1 million in Africa and compiles events reported by Associated Press (AP) and Agence France Presse (AFP), accessed through Lexis-Nexis. The relevant articles have been selected according to a search protocol based upon keyword searches of country names and five terms: “protest,” “riot,” “strike,” “violence,” and “attack”. Before coding, each retrieved article was then examined by a member of the SCAD team, discarded if not related, and otherwise kept and coded into different types of events. The SCAD team double-coded 10 percent of the country-years to verify the accuracy of the coding procedures. Particular attention was devoted to avoid double or triple counting a single event, whenever an event was documented by multiple articles. Last, the geographic location of the event was added by searches on a wide variety of platforms.

##### Definition of Event Types

Following the SCAD codebook, events are classified into different categories depending on events being spontaneous/organized and on the underlying motive of the unrest. The analysis here primarily focuses on events defined as follows:

- *Organized Events*: all events for which “clear leadership or organization(s) can be identified”.
- *Spontaneous Events*: all events for which “clear leadership or organization(s) cannot be identified”.
- *Elections*: events that mention “elections” as the first issue of the social disturbance.
- *Economic Grievance*: events that mention “economy, jobs” and/or “economic resources/assets” as the first issue of the social disturbance.
- *Human Rights*: events that mention “human rights, democracy” as the first issue of the social disturbance.

##### Additional Information on Event Size

The analysis also uses information about the number of participants and the number of fatalities related to violent events, classified into the following categories:

- *No casualties*: Social disturbance characterized by no casualties.
- *Casualties*: Social disturbance resulting in at least one death.
- *Few Participants*: Social disturbance with at most 100 participants (or missing information regarding the size).
- *Many Participants*: Social disturbance with more than 100 participants.

##### Construction of Data for Analysis

Over the period 1990-2017, 17,644 events, localized across 50 African countries, are retrieved. The only event category excluded is Intra-Government violence, which represents events reflecting social conflict within the army or the police forces. Events are linked to the respective country/region based on information about latitude and longitude. To generate a measure of social violence at sub-national levels, only geo-localized events are used. The main measure of social violence denotes the total number of violent events in the country/region in a given year.

*Country-level data:* The data set used in the analysis covers 50 African countries over the period 1990-2017. The number of events ranges between a minimum of 0 to a maximum of 617, with an average of 11.9 events per country/year. More than 77% of all country-year observations exhibit at least one event. For a subset of events (7,750 out of 17,644 events) information on the approximate number of participants of the event is available.

*Sub-national level data:* The data for the analysis at the level of sub-national regions is restricted to events that are geo-coded. Events that are attributed to the country capital because of lack of information are discarded. The data set used in the analysis covers 170 administrative level-1 units in 18 countries over the period 1990-2017. Administrative level-1 units correspond to the largest sub-national administrative units (comparable to federal states in the United States or Bundesländer in Germany). The number of events in a sub-national unit ranges between 0 and 61, with an average of 0.8 events per sub-national unit/year. The share of subnational region-year observations with at least one event is 25%. For 1,858 out of 4,004 events, information on the number of participants of the event is available.

##### Definition of Types of Riots/Protests based on SCAD Narratives

Using information about the actors involved in an event provided by the short narratives that describe a given event allows identifying events involving specific categories. Using a “bag-of-words” approach, the analysis focuses on the following categories:

- *Health Workers*: We identify events involving health workers by a keyword search through all event fields of the keywords: “nurse” OR “doctor” OR “paramedic” OR “midwife” OR ((“health” OR “health care” OR “hospital” OR “medical”) AND (“worker” OR “staff” OR “employee” OR “personnel” OR “professional”)).
- *Civil Servants*: We identify events involving civil servants by a keyword search through all event fields of the keywords: “civil servant” OR “public sector worker” OR “government employee” OR “civil service worker” OR “teacher” OR “professor” OR “government official” OR “city official”.
- *NGOs*: We identify events involving non-governmental and human rights activists by a keyword search through all events fields of the words: “NGO” OR “activist” OR “non-governmental”.

##### Appraisal of SCAD

The SCAD database has several major advantages for the analysis presented here. First, the data focus on social violence defined as social and political unrest, excluding large-scale organized armed conflicts. Access to ART treatment is expected to be particularly relevant for social violence as opposed to large scale conflict, which is not merely affected by individual-level factors –like access to health treatment– but driven by a variety of macro-level factors related to collective action. Second, the data has long time coverage and high data quality. In particular, the procedures of data construction of the SCAD data avoid duplication of events, which is a particularly serious concern for comparable data sets – especially for data sets that (unlike SCAD) rely exclusively on automated text extraction algorithms. Third, the same sources are used for retrieving episodes of violence for all countries/regions/years in the sample – which is of great importance for the consistency of coding. These features (the very consistent definition of events of interest with exclusive focus on social violence, avoiding double-counting, and the consistency in the sources used) explain the fact that the number of total events is lower in the SCAD data than in comparable data sets, such as the ACLED database (described below).

#### A.1.1.2 The Armed Conflict Location And Event Data - ACLED

##### Primary Sources and Data Construction

As alternative data source for social violence, the analysis makes use of the Armed Conflict Location and Event Data – ACLED (Raleigh, Linke, Hegre and Karlsen 2010). These data contain information on all forms of political disorder for nearly 100 countries since 1997. Political violence is defined as “the use of force by a group with a political purpose or motivation”. ACLED includes accurate information about the timing of events, the actors and the exact location, with events reflecting violent acts between and across non-state groups, militias, unnamed agents, violent political agents and riots and protests. ACLED data and documentation about coding procedures can be accessed at https://acleddata.com/curated-data-files/.

ACLED data are collected by experienced researchers and retrieved from a wide range of local, regional and national sources. These sources include local, regional, national and continental media (including Telegram and Twitter); reports from NGOs or international organisations; and information from local conflict observatories. The data collection procedure can be summarized as follows: each week individual researchers scrutinize information on available reports; this information is then aggregated and coded by a first reviewer; a second reviewer cross-checks the available information; notes and details are inspected by a third final reviewer.

##### Definitions of Event Types

Following the ACLED codebook, events are classified into different categories. The analysis here primarily focuses on riots and protests, which are classified as follows:

- *Riots* are violent events where demonstrators or mobs engage in disruptive acts, including but not limited to rock throwing, property destruction, etc. They may target other individuals, property, businesses, other rioting groups or armed actors. Rioters may begin as peaceful protesters, or may be intent on engaging in spontaneous and disorganized violence from the beginning of their actions. Contrary to armed groups, rioters do not use sophisticated weapons such as guns, knives or swords. “Crude bombs” (e.g. Molotov cocktails, petrol bombs, firecrackers) may be used in rioting behavior.
- *Protests* are defined as public demonstrations in which the participants do not engage in violence, though violence may be used against them. Events include individuals and groups who peacefully demonstrate against a political entity, government institution, policy, group, tradition, businesses or other private institutions. Events that are not coded as protests are symbolic public acts such as displays of flags or public prayers (unless they are accompanied by a demonstration), protests in legislatures such as parliamentary walkouts or MPs staying silent, strikes (unless they are accompanied by a demonstration), and individual acts such as self-harm actions (e.g. individual immolations or hunger strikes).

##### Construction of Data for Analysis

Over the period 1997-2017, 41,407 events, localized across 50 African countries, are retrieved. Events are linked to the respective country/region based on information about latitude and longitude.

*Country-level data:* The data set covers 50 African Countries over the period 1997-2017. The number of events ranges between a minimum of 0 to a maximum of 1,782, with an average of 38.7 events per country/year. More than 81% of the country-year observations exhibit at least one event.

*Sub-national level data:* The data for the analysis at the level of sub-national regions is restricted to the subset of events that are geo-coded. The data set used in the analysis covers 170 administrative level-1 units in 18 countries over the period 1997-2017. The number of events ranges between 0 and 556, with an average of 2.8 events per sub-national unit/year. The unconditional probability of experiencing a social violence event for a subnational unit is 38 percent. The share of region-year observations with at least one event exceeds 38%.

##### Appraisal of ACLED

The main advantage of the ACLED database for the purpose of this analysis is its reliance on a rich pool of primary and secondary sources, which allows mapping a relatively high number of events. The main shortcoming is that the pool of sources is not necessarily stable across country/regions and years. These features make it an ideal secondary data source to perform robustness checks for the results obtained with the SCAD data.

### A.1.2 Data from the Uppsala Conflict Data Program (UCDP)

**Primary Sources and Data Construction.** As alternative data source for social violence, we also make use of data on conflict events collected by the Uppsala Conflict Data Program (UCDP GED). The UCDP data contain violent events, defined as “the incidence of the use of armed force by an organized actor against another organized actor, or against civilians, resulting in at least 1 direct death in either the best, low or high estimate categories at a specific location and for a specific temporal duration (Sundberg and Melander 2013). The data contain information about location, timing, and involved actors of events.

UCDP data are based on automated searches of global news companies, media, international organizations, NGOs, historical achives and other sources of information, which are then evaluated by human coders. UCDP data and documentation can be accessed at https://www.pcr.uu.se/research/ucdp/.

**Definitions of Event Types.** UCDP data contain information on armed conflicts, which represents a contested incompatibility that concerns government and/or territory where the use of armed force between two parties, of which at least one is the government of a state. UCDP data only contain organized events involving at least one fatality; incidents without information on fatalities are not included. Conflict events in the UCDP data that meet these criteria therefore represent organized violence of considerable intensity.

**Construction of Data for Analysis.** Over the period 1990-2017, 35,264 events, localized across 50 African countries, are retrieved. Events are linked to the respective country.

*Country-level data:* The data set covers 50 African Countries over the period 1997-2017. The number of events ranges between a minimum of 1 to a maximum of 583, with an average of 55.1 events per country/year. All of the country-year observations in Africa exhibit at least one event.

**Appraisal of UCDP Data.** For the purpose of the analysis, the UCDP data represent information about social conflict that involves organized violence of substantial intensity. This measure complements the SCAD data used in the baseline analysis, which excludes all events defined as “a contested incompatibility that concerns government and/or territory where the use of armed force between two parties, of which at least one is the government of a state, results in at least 25 battle-related deaths in one calendar year”. This makes it an ideal source of information to investigate the mechanism and the sensitivity of the results to different types of violent conflict.

#### A.1.2.1 Intimate Partner Violence from Demographic and Health Surveys (DHS)

Information about intimate partner violence is based on data from the Demographic and Health Surveys (DHS) Program (DHS Program Spatial Data Repository, funded by USAID; spatialdata.dhsprogram.com [Last accessed August 3, 2020]). The DHS collects nationally representative data on health and population in developing countries. Data on domestic violence in the DHS are available for some, but not all, countries surveyed, as the information on domestic violence stems from an optional module of questions (for further information, see: https://dhsprogram.com/data/Guide-to-DHS-Statistics/17_Domestic_Violence.htm#References4). The set of questions related to domestic violence is further administered to a sub-sample of randomly selected respondents.

##### Construction of Data for Analysis

Regional-level information on domestic violence is available for the following countries (years): Burkina Faso (2010), Cameroon (2004, 2011), Cote d’Ivoire (2011), Ethiopia: 2016), Ghana (2008), Kenya (2003, 2008, 2014), Malawi (2004, 2010, 2015), Mali (2006, 2012), Rwanda (2005, 2010, 2014), Senegal (2017), Tanzania (2010, 2015), Zambia (2007, 2013), Zimbabwe (2005, 2010, 2015). The analysis is based on data aggregated by the statcompiler (see https://www.statcompiler.com/en/); we retrieved information on the percentage of women aged 15-49 who have experienced physical violence in the past 12 months (often or sometimes). This delivers 219 observations, across 124 Administrative level-1 units within 13 countries, between 2003 and 2017. Around one-fifth of women in the sample (20,2 percent) experienced physical violence (often or sometimes) in the 12 months before the survey. The variable ranges from a minimum of 0.2 (Centre-Nord, DHS region, Burkina Faso, 2010) to a maximum of 0.575 (Mara, DHS region, Tanzania, 2010).

##### Appraisal of Intimate Partner Violence from DHS

For the purpose of our analysis, this data is of great importance for being able to study the extent of individual violence taking place in the household context. A drawback of this data is coverage and the small number of observations, which restricts the statistical analysis that can be conducted.

### A.1.3 Data: HIV Prevalence

#### Data at Country Level

At the country level, data for HIV prevalence are obtained from UNAIDS (downloaded from the World Bank data platform https://data.worldbank.org/indicator/SH.DYN.AIDS.ZS). Since the exact number of individuals living with HIV cannot be determined in many countries, the best possible approximation is obtained through model estimates. For each country, UNAIDS composes teams of experts (epidemiologists, demographers, monitoring and evaluation specialists and technical partners) who collect and process data to produce estimates for HIV prevalence. UNAIDS collects all country estimates and reviews them through the work of the Strategic Information and Monitoring Division, in order to guarantee that estimates are comparable across countries over time. The construction of estimates differs across countries with high and low levels of HIV prevalence:

- For *high-level HIV epidemic countries* the estimates are based on data from surveillance among pregnant women and from nationally representative population-based surveys.
- For *low-level HIV epidemic countries* the estimates are based on data from sub-populations at high risk of HIV infections that are combined with nationally representative surveys to produce the estimates.

#### Construction of Data for Analysis – Country level

The analysis is based on HIV prevalence among the population group with age 15-49 years. Formally, this variable is defined as HIV Prevalence = number HIV infected (15-49 years) / total population (15-49 years). The data set covers 50 African countries over the period 1990-2017. The average HIV Prevalence (15-49) is 0.047, ranging from a minimum of 0.001 to a maximum of 0.285 (Swaziland in 2013 and 2014).

#### Data at Sub-national Level

HIV prevalence at the sub-national level (administrative level-1 units) is constructed using DHS survey data (DHS Program Spatial Data Repository, funded by USAID; spatialdata.dhsprogram.com [Last accessed August 3, 2020]). DHS data are, to the best of our knowledge, the most comprehensive source of sub-national information to map HIV prevalence. The data has been retrieved from https://www.statcompiler.com/en/. Since no information from DHS waves was available for some countries before 2001, the analysis makes use of information about HIV prevalence from waves up to 2006 for a subset of countries. Specifications of the empirical framework with country-year fixed effects address possible bias arising from this specific data limitation.

#### Construction of Data for Analysis – Sub-national level

The analysis is based on HIV prevalence among the population group with age 15-49 years. Formally, this variable is defined as HIV Prevalence = number HIV infected (15-49 years) / total population (15-49 years). The data set contains HIV prevalence for a total of 170 regions in 18 African countries. The average HIV Prevalence (15-49) is 0.068, ranging from a minimum of 0.001 to a maximum of 0.297 (Leribe region in Lesotho).

### A.1.4 Data: Latent HIV Exposure based on Geography

An alternative measure of cross-country exposure to HIV and scope for treatment is constructed based on historical geographic origin of the HIV epidemic and its subsequent spread. According to current knowledge the most infective strain of the HIV virus crossed from chimpanzees to humans probably before 1920 in Cameroon, while the beginning of the spread of HIV across Africa has been traced back to Kinshasa around 1920 (Gao, Bailes, Robertson, Chen, Rodenburg, Michael, Cummins, Arthur, Peeters, Shaw, Sharp and Hahn 1999, Faria, Rambaut, Suchard, Baele, Bedford, Ward, Tatem, Sousa, Arinaminpathy, Pépin, Posada, Peeters, Pybus and Lemey 2014). By 1980, about half of human infections in the Democratic Republic of Congo were observed outside of Kinshasa. The virus subsequently diffused out of DRC, first towards the great lakes area and then along the East of Africa, eventually reaching the Mediterranean basin and South Africa as well as the North-West towards Nigeria during the 1990’s (Kalipeni and Zulu 2012). In general, the spread and prevalence of HIV in a population depends on multiple factors, including population density, economic activities, and public health policies. The spread of HIV within Africa, however, was affected strongly by geographical features. While equatorial forests and deserts or semidesert areas constituted obstacles, rivers such as the Congo River played an important role by facilitating the spread of the virus out of Kinshasa. These geographic features are considered as prime determinants of the comparatively intense initial but uneven and non-concentric diffusion of HIV to the South-East of Africa (Faria et al. 2014).

**Construction of Latent Exposure Index (HIV*_geo_*).** An index of latent exposure to HIV is constructed based on the effective (non-geodesic) distance from the origin of the spread of the virus in Kinshasa. The measure exclusively uses information on first nature geography. A generalized version of the Dijkstra algorithm known as Fast Marching Method (Sethian 1996, Sethian 1999) is used to compute the shortest path to Kinshasa from any location, defined as centroids of grid-cells. These grid-cells span the entire African continent and are transformed that span the entire African continent and that are transformed to an equal area projection using the Africa Albers Equal Area Conic projection (ESRI:102022). The algorithm assigns a measure of effective distance that accounts for propagation costs for HIV transmission. These costs depend exclusively on first nature geographic features of the shortest travel path from each location to Kinshasa. This accounts for the iso-cost contour expanding more slowly through deep forests, very rugged terrains, or deserts and thereby reflects the fact that paths along such geographies involve higher effective distance, paralleling applications of similar procedures (Allen and Arkolakis 2014). Importantly, the measure reflects the relative propagation speed, or distance, from Kinshasa to a location relative to other locations.

Denoting by *c_i_* the instantaneous cost associated with passing any given cell *i*, a parameter *x_i_ ≥* 0 constitutes the inverse of costs, with *c_i_* = 1*/x_i_*. Accounting for the role of geography for the spreading of HIV, maximum instantaneous costs are associated with grid-cells *i* that exhibit inhospitable conditions as reflected by deserts or deep forests, or with monthly average maximum temperatures above 38◦C, such that *x_i_* = 0, rendering these cells virtually impenetrable and ignored by the fast marching method in the identification of the closest path from each location (grid-cell) to Kinshasa. For all other cells, the instantaneous costs are associated with a level *x_i_ >* 0. The minimum instantaneous cost is associated with cells that are penetrated by major rivers and water reservoirs. For all other cells, the instantaneous costs increase with the intensity of forest coverage and the roughness of the terrain. For any origin-destination pair, the algorithm delivers a (normalized) distance, which when summed up delivers a measure of latent HIV exposure of a location in terms of its spreading distance to Kinshasa.

Since the distribution of human populations in Africa is uneven with the vast majority of locations hosting no sizable human settlements and only a minority of locations (less than 20 percent) hosting comparatively large communities, only locations (grid-cells of 5*×*5km) that exhibit sufficiently large population density are associated with positive HIV potential and considered for the analysis. Population density data are as of year 2000 (Center for International Earth Science Information Network - CIESIN 2018, Tatem 2017). As baseline, the distances are computed for a population threshold for locations (grid-cells) of at least 16903 inhabitants, which corresponds to the 80th percentile (8th decile) of the distribution of population density. As alternative specifications, we computed the distances based on a population threshold of 20,000 inhabitants per cell (which is equivalent to around 1,000 inhabitants per square km), which corresponds to the 17.5% most densely populated cells (or the 82.5th percentile), as well as based on a population threshold of 10,000 L(egcenod rresponding to the top 2Organ8ized .Vio6lent Riotpercent of Ltehgened population density distribution). In a last step, cell-le% vScead Elventdata are aggregated toLimitted hS trikee country level.

The resulting measure of latent HIV exposure and scope for treatment in the year 2000 is a function of effective, purely geography-based distance from Kinshasa. Figure A1 displays the actual HIV prevalence in 2001 at the country level (left panel) and the corresponding measure of purely geography-based HIV exposure (right panel, both standardized).

**Figure A1:**
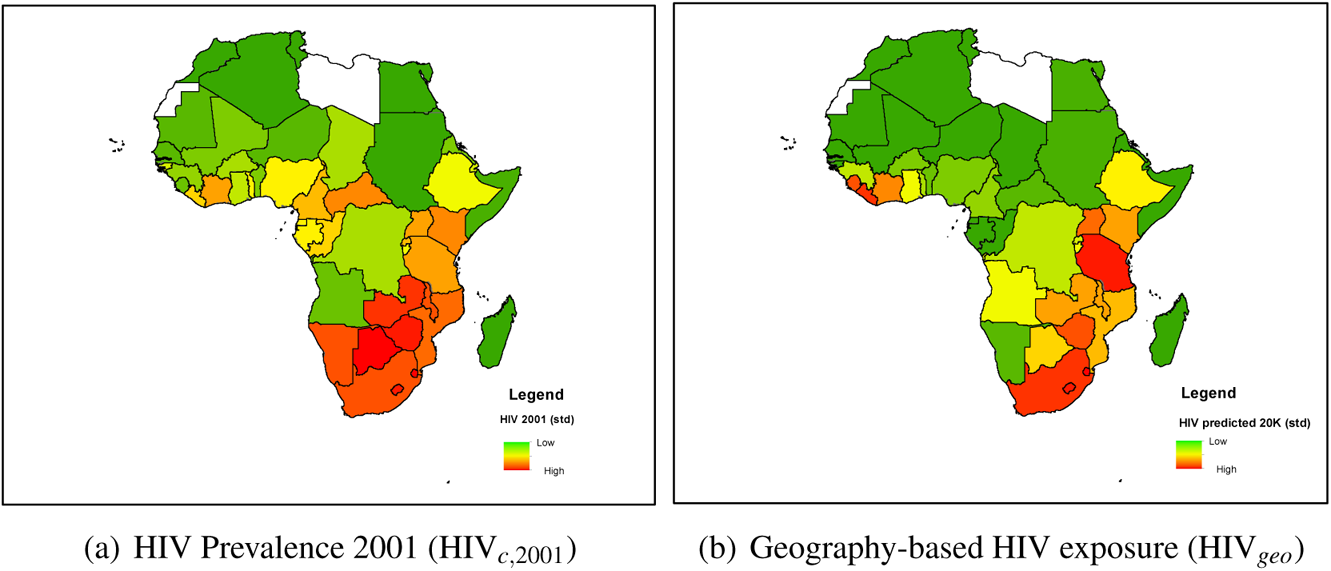
Actual HIV Prevalence and Latent HIV Exposure (Country Level) Note: Panel (a): HIV prevalence in 2001; data source: UNAIDS. Panel (b): Latent HIV exposure based on effective distance to Kinshasa, based on fast marching method algorithm; country-level averages. For greater comparability, measures in both panels have been standardized.

### A.1.5 Data: Antiretroviral Therapy (ART) Coverage

Data for Antiretroviral Therapy (ART) coverage at the country level is obtained from UNAIDS (downloaded from the World Bank data platform https://data.worldbank.org/indicator/SH.HIV.ARTC.ZS). ART coverage indicates the percentage of all individuals living with HIV who receive antiretroviral therapy. In each country, local facilities administering antiretroviral therapy hold registers that are compiled and sent to national authorities on a routine basis. UNAIDS requests countries to submit these data on the 31st of March every year through an on-line reporting tool. The tool features several quality checks in order to avoid reporting errors; UNAIDS further validates the data, comparing them with information from alternative sources.*

#### Construction of Data for Analysis (ART Cov)

The analysis makes use of ART coverage measured as the share of treated individuals among infected individuals. Formally, this variable is defined as ART Coverage (out of Infected Individuals) = number ART treated / number HIV-infected. As alternative measure for the robustness analysis, the results are cross-validated using the number of treated individuals out of the total population. Formally, this variable is defined as ART Coverage (percentage population) = number ART treated / total population. The data set covers 50 African countries over the period 1990-2017. The average ART Coverage (out of Infected Individuals) is 0.12, ranging from a minimum of 0 to a maximum of 0.85 (Swaziland in 2017). The identification strategy makes use of data for 53 low and middle income countries outside Africa for which ART coverage is available.

### A.1.6 Data: Antiretroviral Therapy (ART) Costs and Prices

Data on the time-series of ART Prices and Production Costs are based on the combination of information from two different sources: The WHO Global Price Reporting Mechanism (GPRM) and the Global Fund Pooled Procurement Mechanism Reference Pricing. The WHO Global Price Reporting Mechanism (GPRM) was created in 2003 to support transparency in the market of antiretroviral (ARV) drugs. Until 2013, GPRM reported prices and volumes of ARV drugs sold on international markets covering 132 countries (35 low-income countries, 44 lower-middle income, 36 upper-middle income and 17 high income countries). The main providers of data were the Global Fund, PEPFAR, UNITAID, and the procurement organizations working with them, such as the Clinton Foundation, Crown Agent, the Global Drug Facility (GDF), the International Dispensary Association (IDA HIV/AIDS), USAID/Deliver, Mission Pharma, Management Sciences for Health (MSH), the Partnership for Supply Chain Management (PFSCMS), the United Nations Development Programme (UNDP), the United Nations Children’s Fund (UNICEF), and the WHO/Contracting and Procurement Service (WHO/CPS). From 2004 to 2012 the GPRM covered between 70 and 80 percent of total volume of ARV drug transactions (Perriëns, Habiyambere, Dongmo-Nguimfack, Hirnschall et al. 2014). The analysis here makes use of price data for active pharmaceutical ingredient (API) from GRPM. These data contain the median price for the most relevant treatment regimens and their API over the period 2003-2012. Since 2015, the Global Fund Pooled Procurement Mechanism Reference Pricing has published quarterly reports containing reference prices for ARV medicines. Reports from the Global Fund were retrieved from https://www.theglobalfund.org/en/sourcing-management/health-products/antiretrovirals/ and https://www.who.int/hiv/amds/gprm/en/.

**Construction of Data for Analysis – Prices (ART Price).** The main analysis focuses on the median price of a specific and widely used first line treatment for adults, ZDV-3TC-EFV, which represents a combination of three reverse-transcriptase inhibitors: 300mg of Zidovudine (ZDV), 150mg of Lamivudine (3TC) and 600 mg of Efavirenz (EFV).^†^ Since data are available for the period between 2003 and 2012 and from 2015 to 2017, data for the years 2013 and 2014 have been linearly interpolated. The prices for the ZDV-3TC-EFV first line treatment ranges from 644$ US per patient per year in 2003 to 102$ US in 2017. For comparability with the results based on ART coverage in non-African countries, prices are converted into a reverse index.^‡^

#### Construction of Data for Analysis – Costs (ART Cost)

The production costs of the same first line treatment ZDV-3TC-EFV are available only for selected years, namely 2005, 2007, 2010 and 2012. The time-series of production cost were constructed as follows. First, exploiting prices and costs for available years, we computed the mark-up of pharmaceutical companies (ARV medicine prices/API costs). Second, this mark-up is assumed to be constant before 2005 and after 2012, when cost information is not available; in this way it is possible to compute, in a very conservative way, costs over the periods 2003-2005 and 2013-2017 by combining the predicted mark-ups with the complete time-series of price data. Third, for the missing years between 2005 and 2012, we interpolated the data to build a continuous estimate of the production costs. The API production cost for ZDV-3TC-EFV ranges from a maximum of 477$ in 2003 to a minimum of 86$ per patient per year in 2017. For comparability with the results based on ART coverage in non-African countries, costs are converted into a reverse index.^§^

#### Construction of Data for Analysis – Synthetic Price Index (Synth. Price)

As alternative, a synthetic index of global prices is constructed only exploiting information about the initial price prior to the expansion of ART demand (and thus potential influence of donors on the price development). This synthetic price index is therefore, by construction, unrelated to political interventions and to any other sort of demand-driven price decline. Concretely, a time series of synthetic prices for the first line treatment for adults, ZDV-3TC-EFV, is constructed following previous research on the evolution of drug prices after the end of patent exclusivity, and after the introduction of generic drugs (with the related increase in competition). Existing evidence shows that market prices typically display a sharp initial decline that is followed by more moderate reductions as market prices converge to the limit price, which ensures non-negative profits by generic producers (in terms of a minimum mark-up over production cost). The documented patterns are compatible with a proportional reduction of the mark-up in each year (Perriëns et al. 2014, Conti and Berndt 2014, Dave, Hartzema and Kesselheim 2017). The synthetic price index is obtained by computing the initial extra mark-up for the regimen ZDV-3TC-EFV in 2003 and then constructing the series of prices that would have been observed on the global markets if the mark-up were reduced each year after 2003 by a fixed proportion *x*.^¶^

### A.1.7 Data: Other Variables and Sources

#### A.1.7.1 Country-Level Data

- **Time Trends for African Regions**: The main specification accounts for differential trends in different regions of Africa, based on the classification of subregions according to the United Nations geoscheme for Africa. Countries are coded as follows: *Central Africa:* Burundi, Cameroon, Central African Republic, Chad, Congo, Dem. Rep. Congo, Rep. Equatorial Guinea, Gabon; *East Africa*: Comoros, Djibouti, Eritrea, Ethiopia, Kenya, Madagascar, Rwanda, Somalia, South Sudan, Sudan, Tanzania, Uganda; *North Africa*: Algeria, Egypt, Arab Rep. Mauritania, Morocco, Tunisia; *Southern Africa*: Angola, Botswana, Eswatini (Swaziland), Lesotho, Malawi, Mozambique, Namibia, South Africa, Zambia, Zimbabwe; *West Africa*: Benin, Burkina Faso, Cabo Verde, Cote d’Ivoire, Gambia, Ghana, Guinea, Guinea-Bissau, Liberia, Mali, Niger, Nigeria, Senegal, Sierra Leone, Togo.
- **GDELT Data**: Robustness was conducted for data on violent events collected by the GDELT project (Global Database of Events, Language, and Tone). GDELT contains global, machine-coded and georeferenced information of events of various types based on open source data from news media collected from numerous websites. GDELT data and documentation can be accessed at https://www.gdeltproject.org/data.html. Relevant events have been extracted based on the CAMEO classification for “protests” (classification 14, which also includes non-violent and violent protests, as well as riots). As result of the machine-based coding process, geolocalization of GDELT data is often unreliable or misleading.(Raleigh and Kishi 2019) Consequently, data are applied to country-level analysis only.
- **GDP**: GDP data, in constant 2010 US$, are taken from the World Bank data platform (https://data.worldbank.org/indicator/NY.GDP.MKTP.KD). GDP at purchaser’s prices is the sum of gross value added by all resident producers in the economy plus any product taxes and minus any subsidies not included in the value of the products. It is calculated without making deductions for depreciation of fabricated assets or for depletion and degradation of natural resources. Data are in constant 2010 U.S. dollars. Dollar figures for GDP are converted from domestic currencies using 2010 official exchange rates. For a few countries, where the official exchange rate does not reflect the rate effectively applied to actual foreign exchange transactions, an alternative conversion factor is used.
- **Population**: Population data are taken from United Nations Population Division, Census reports, Eurostat, United Nations Statistical Division, U.S. Census Bureau and Secretariat of the Pacific Community and downloaded from the World Bank data platform (https://data.worldbank.org/indicator/SP.POP.TOTL). Total population is based on the de facto definition of population, which counts all residents regardless of legal status or citizenship. The values represent mid-year estimates.
- **Life Expectancy**: Life expectancy data are taken from United Nations Population Division and Census and downloaded from World Bank data platform (https://data.worldbank.org/indicator/SP.DYN.LE00.IN). Life expectancy at birth indicates the number of years a newborn infant would live if prevailing patterns of mortality at the time of her birth were to stay the same throughout her life.
- **Malaria prevalence**: Malaria prevalence data are collected by WHO and downloaded from the World Bank data platform (https://data.worldbank.org/indicator/SH.MLR.INCD.P3). Malaria incidence is measured as the number of new cases per year per 1000 population at risk.
- **Malaria treatment prices**: Malaria treatment prices are collected by the Bill and Melinda Gates Foundation. The analysis focuses on the price of the most relevant drug against malaria: Artesiminin. Information about the price for artesiminin between 2004 and 2015 is obtained from the report “Novel Artemisimin Manufacturing Technologies: Request for Proposal” published by the Bill and Melinda Gates Foundation.^||^
- **Global Fund**: Global Fund data measure the amount of aid (in USD) disbursed by the Global Fund to a country in a given year. Data have been downloaded from the Global Fund data platform (https://data.theglobalfund.org/investments/home).
- **Democracy**: The Institutionalized Democracy score ranges between 0 and 10, with a higher score representing more democratic institutions. The combined Polity score is a composite of the Institutionalized Democracy score and a corresponding score of Institutionalized Autocracy, which is subtracted from the Institutionalized Democracy score. The Revised Combined Polity Score (Polity 2) represents a modified version of the Polity score and ranges from −10 to +10. Data have been downloaded from the Polity V Project dataset “Political Regime Characteristics and Transitions, 1800-2018” (http://www.systemicpeace.org/inscrdata.html).
- **Institutional Quality**: The quality of institutions is measured using indices in six broad dimensions of governance, based on data from the Worldwide Governance Indicators (2020 Update, data have been downloaded from http://www.govindicators.org). The dimensions include Voice and Accountability (va), Political Stability and Absence of Violence/Terrorism (pv), Government Effectiveness (ge), Regulatory Quality (rq), Rule of Law (rl), and Control of Corruption (cc). Each index represents an estimate of governance quality in standard normal units, ranging from approximately −2.5 (weak) to 2.5 (strong) governance performance.

#### A.1.7.2 Sub-national Level Data

**Malaria Prevalence**: Malaria prevalence at the sub-national level (administrative level-1 unit) has been constructed using DHS survey data (https://www.statcompiler.com/en/). DHS collects information about children who had malaria in the 5 years prior the interview. This measure is used as a proxy for malaria prevalence in a given region, using for each country the survey closest to 2001. The data set covers 103 regions across 10 countries.

- **Trust in Institutions**: Trust in institutions is constructed from the Afrobarometer project, using survey data from waves 1–6 (covering the period 1999-2016) (BenYishay, Rotberg, Wells, Lv, Goodman, Kovacevic and Runfola 2017). Afrobarometer surveys measure the social and political atmosphere in a country through face-to-face interviews with a random sample of 1200-2400 people per country. The surveys cover 12 countries during the first wave and up to 36 countries during the sixth wave. Survey responses are associated to administrative regions based on the latitude and longitude of survey respondents’ locations. The analysis focuses on trust in institutional representatives, relying on individual answers to questions on trust. For each question, respondents had four different answer categories, two related to positive and two related to negative responses, measured by a categorical variable ranging from 1 to 4 (or 0 to 3). The detailed wording of the questions can be found in the Afrobarometer codebooks (https://www.afrobarometer.org/data/merged-data). The questions refer to trust in

– Parliament (item Q43B in wave 2, Q55B in wave 3, Q49B in wave 4, Q59B in wave 5, Q52B in wave 6)
– Local Government (item Q43E in wave 2, Q43E in wave 2, Q55D in wave 3, Q49D in wave 4, Q59E in wave 5, Q52E in wave 6)
– Police (item *trspol* in wave 1, Q43I in wave 2, Q55H in wave 3, Q49G in wave 4, Q59H in wave 5, Q52H in wave 6)
- **Policy Approval**: Approval of policies is constructed from the Afrobarometer project, using survey data from waves 1–6 (covering the period 1999-2016) similar to trust in institutions (BenYishay et al. 2017). The analysis focuses responses to questions about how well the government handles different matters. For each question, respondents had four different answer categories, two related to positive and two related to negative responses. Approval is coded as binary variable, distinguishing between replies “very badly/fairly badly” and “fairly well/very well”. The detailed wording of the questions can be found in the Afrobarometer codebooks (https://www.afrobarometer.org/data/merged-data). The questions refer to approval in the following dimensions:

– Approval to the handling of combating HIV/AIDS (item *pfghiv* in wave 1, Q45L in wave 2, Q65K in wave 3, Q57L in wave 4, Q65M in wave 5).
– Approval to addressing educational needs (item *pfgedu* in wave 1, Q45g in wave 2, Q65g in wave 3, Q57h in wave 4, q65h in wave 5, Q66h in wave 6).
– Approval to economic policy (average of share of positive answers to questions referring to managing the economy, creating jobs, keeping prices stable, and reducing inequality):

* managing the economy (item Q45A in wave 2, Q65A in wave 3, Q57A in wave 4, Q65A in wave 5, Q66A in wave 6)
* job creation (item Q45B in wave 2, Q65B in wave 3, Q57C in wave 4, Q65C in wave 5, Q66C in wave 6)
* price stability (item Q45C in wave 2, Q65C in wave 3, Q57D in wave 4, Q65D in wave 5, Q66D in wave 6)
* reducing the income between rich and poor (item Q45D in wave 2, Q65D in wave 3, Q57E in wave 4, Q65E in wave 5, Q66E in wave 6)
– Approval to health policy (related to improving basic health services, item *pfghlt* in wave 1, Q45F in wave 2, Q65F in wave 3, Q57G in wave 4, Q65G in wave 5, Q66G in wave 6).
– Approval to policy aimed at reducing crime (item Q45E in wave 2, Q65E in wave 3, Q57F in wave 4, Q65F in wave 5, Q66F in wave 6).

## A.2 Identification

**Identification Assumptions.** The identification strategy is based on the interaction of cross-country heterogeneity in HIV exposure and thus scope for ART expansion and time-varying measures of ART expansion. For each specific combination of measures, the crucial assumption for identification is that this interaction is exogenous. While conceptually capturing the same underlying phenomenon, the different instruments of the variation in ART access over time differ in terms of data quality and potential concerns regarding the validity of the identifying assumptions.

**ART Price**: The median world price of ART treatment regimens represents a valid instrument, as it is exogenous to the intensity of social and political pressure or other country-time-specific factors that might affect ART coverage and conflict simultaneously. This implies that the instrument is only indirectly informative about the availability of ART treatments in a country but it has the advantage that the instrument does not respond, by construction, to the country-specific level of social violence or any policy intervention specifically targeted at a given country and given year. An advantage of this instrument is that, conceptually, it is more directly related to actual ART treatment in a country as compared to information on ART coverage outside Africa since the reduction in prices constitutes the ultimate driver of the increase in ART coverage. A potential concern for the quantitative interpretation is that a reduction in the prices paid by a government frees budget resources that can be used for alternative policies and, accordingly, lead to a reduction of e.g. protests or strikes. Limitations of this instrument are related to shorter time coverage of data compared to ART coverage outside Africa. Moreover, to the extent that prices for ART treatments might be determined by monopolistic pricing of pharmaceutical multinational corporations, international organizations might have had an indirect effect through their influence on price negotiations. This would show up in terms of a decline in prices due to lower mark-ups over costs, raising concerns about simultaneity.

**Figure A2:**
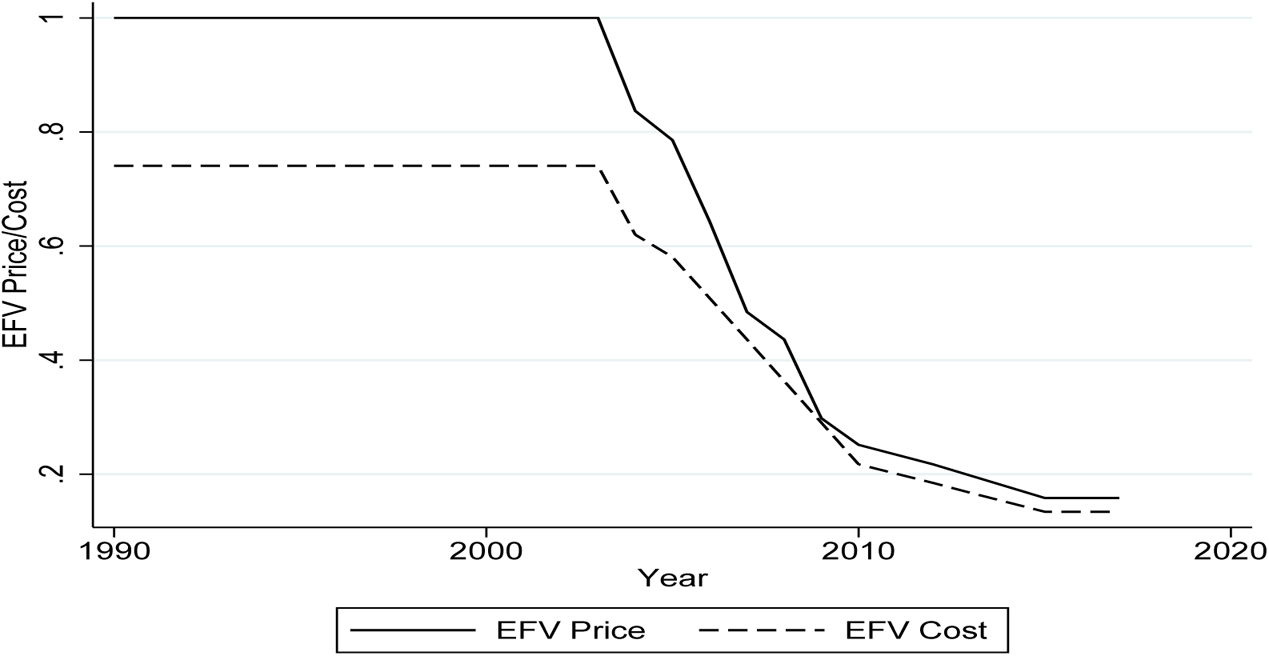
GLOBAL VARIATION IN PRICE AND COST OF ART (ART PRICE, ART COST) Note: The plot captures the global variation in the median world price of ART treatment regimens ZDV-3TC-EFV (ART Price) used in the baseline instrumentation in combination with cross-sectional heterogeneity in the scope for ART expansion, as well as the alternative construct based on the cost of the ZDV-3TC-EFV treatment regimens (ART Cost). Levels are normalized to highest median price in the sample.

**ART Cost**: The conceptual advantage of this instrument is that the decline in global production costs is largely related to the increase in the amount of treatments produced by international laboratories worldwide. The reduction in costs of production maps also into the reduction in prices, but the time variation of the two series differs because of variation in the markups charged by pharmaceutical companies, particularly due to increasing competition associated with the introduction of active principles produced as generic. The validity of the ART cost instrument is based on similar arguments as that of ART prices, but has the appealing feature of not relying on changes of markups of pharmaceutical companies, which might be influenced by political pressure. This makes the information about cost even less susceptible to pressures by international organizations since variation is related to an increase in research competition and patent expiration. Hence, conceptually, the use of global production costs is the preferred instrument. However, this instrument is subject to more severe data limitations in terms of availability and coverage, which requires interpolation of data for years with missing information. This reduces the variability of the measure and leads to a slightly lower strength of the first stage regression.

**Synthetic Price**: The synthetic price instrument is based on price data prior to the major expansion of the first line of ART treatment in 2003, and on price data after the major expansion (2015-2017). For the intermediate time period, the price index is constructed based on the assumption of a constant proportional decline of the mark-up in each year. This decline approximates the global dynamics of treatment costs and prices. The appealing feature of this price index is that only two data points are involved in its construction, alleviating potential concerns about a demand-driven price decline that violates the exclusion restriction due to systematic variation in prices in response to an ART expansion in particular countries.

**ART Cov**: The ART coverage in other low and middle income countries outside Africa captures effective variation in access, and thus comes closest to the variation captured by the instrumented variable (ART coverage in African countries). The use of global ART coverage outside Africa is conceptually not affected by the level of ART coverage in a specific country. Moreover, data quality and coverage is good and the inclusion of year fixed effects accounts for global shocks. However, a direct role of international actors or organizations in extending ART coverage worldwide could raise potential concerns about simultaneity.

**Instrument Selection.** A battery of statistical tests was performed to verify the first stage relevance and instrument validity. In light of the use of multiple instruments (ART Price, ART Cost, Synth. Price, ART Cov), Sargan-Hansen tests were performed on a series of 2SLS regressions which included every possible combination of the four instruments. The null hypothesis of the joint validity of all instruments was never rejected. Since for no possible combination of instruments the null hypothesis could be rejected, this suggests that at least one instrument per regression is valid. Likewise, tests of redundancy of instruments revealed that none of the instruments is redundant in the sense that asymptotic efficiency of the estimation is improved by using them. Since the instruments are highly correlated, the null of orthogonality is rejected for each combination of instruments. These results are reassuring even if (by construction) not definitive about the validity of the instruments in terms of exclusion restriction or about the dominance of one particular instrument. At the same time, they suggest the use of a single instrument at a time, rather than a combination of instruments.

**Alternative Instrument for Cross-Country Exposure and Treatment Scope: Geography-Based Exposure.** Another potential concern refers to the exogeneity of the cross-sectional variation in scope for ART expansion, as proxied by HIV prevalence in 2001. To investigate the robustness of the results with respect to the measure of cross-country heterogeneity in HIV prevalence in 2001 as measure of exposure and thus scope for ART expansion, the analysis has been repeated for the purely geography-based measure of HIV exposure as a function of effective, geography-based distance from Kinshasa, the historical starting point of the HIV epidemic in Africa. By construction, this measure is not driven by institutional, cultural or political features that could have affected the evolution of the HIV epidemic in a country and that might invalidate the exclusion restrictions.

**Figure A3:**
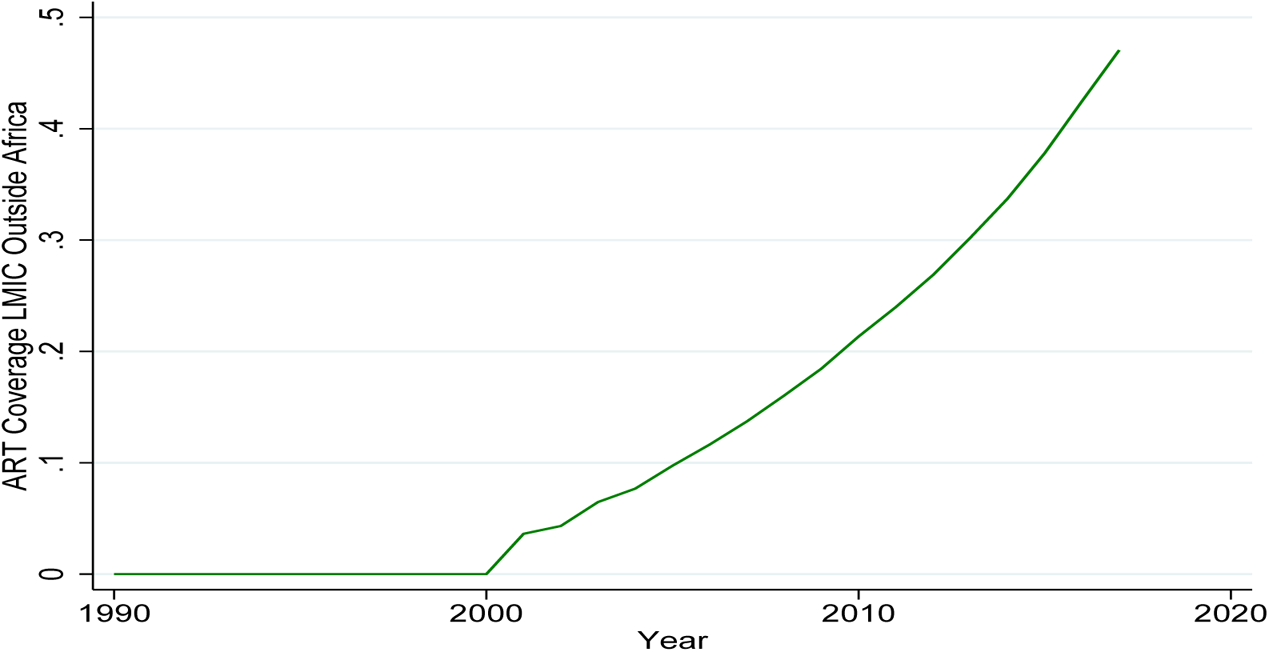
GLOBAL VARIATION IN ART COVERAGE (ART COV) Note: The plot captures the global variation in ART Coverage (ART Cov) in Low-/Middle-Income Countries that is used in the baseline instrumentation in combination with cross-sectional heterogeneity in the scope for ART expansion.

**Identification Using Sub-National Variation.** An alternative approach to identify the coefficient of interest is to use data at the level of sub-national regions. The advantage of this approach is that the estimation of the coefficient of interest can be conducted in an empirical specification that includes region fixed effects and country-specific time trends. This specification thereby implicitly accounts for many of the confounds for identification at the country level. In particular, health policies are mostly under the control of national governments; international aid by donors and international organizations as well as regulations of patents, procurements of treatments and agreements with pharmaceutical companies are typically organized at the country level; strikes, demonstrations and protests exerting pressure on health provision are likely to trigger responses by national governments and donors only if they are sufficiently visible and important at country levels; social disruptions of lower scale, or even intimate partner level violence, should not be expected to have a major impact on national or international policies. The identification of the effect from variation within countries thus absorbs many of these potential confounds in country effects and trends.

The disadvantage of this approach is that without reliable geo-referenced and time-varying data on ART coverage at the region level it is not possible to conduct an analysis based on a 2SLS approach as at the country level. Instead, the analysis is based on an intention-to-treat approach that relates social violence directly to the instrumental variables in terms of cross-region differences in the scope for treatment, *Z_r_*, interacted with time variation in global access to ART. The corresponding empirical model is given by,

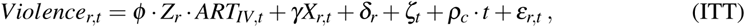

where violent events at the level of sub-national, administrative regions *r* in year *t* are the dependent variable; the term *Z_r_ · ART_IV,t_* represents the interaction between the scope for ART treatments in terms of HIV prevalence across regions prior to the ART expansion, and the time variation in ART access as described in the baseline analysis. Contrary to country-level data, HIV prevalence is not available for the same year (2001) in all regions; instead, *Z_r_* measures the HIV prevalence in the respective region in the year closest to 2001 for which data are available. *X_r,t_* are region-level controls, *δ_r_* and *ζ_t_* represent region and year fixed effects, respectively, and *ρ_c_ · t* represents a country-specific linear trend. Throughout, the main effect (linear term) of HIV prevalence, *Z_r_*, is absorbed by the (sub-national) regional fixed effects, and the time-varying instrument for ART coverage, *ART_IV,t_*, is absorbed by the year fixed effects. The validity of the region level analysis implies similar identification assumptions as at the country level.

## A.3 Additional Tables

### A.3.1 Descriptive Statistics

**Table A1:** Summary Statistics, Country Level

|  | (1) |  |  |  |  |  |
| --- | --- | --- | --- | --- | --- | --- |
|  | count | mean | min | max | p50 | sd |
| Social Violence, SCAD | 1394 | 11.879 | 0.000 | 617.000 | 4.000 | 31.316 |
| Social Violence(log), SCAD | 1394 | 1.610 | 0.000 | 6.426 | 1.609 | 1.281 |
| Social Violence (per 100K capita), SCAD | 1394 | 0.082 | 0.000 | 2.089 | 0.034 | 0.143 |
| Social Violence (log per 100K capita), SCAD | 1394 | 0.072 | 0.000 | 1.128 | 0.034 | 0.109 |
| Protests/Riots, ACLED | 1044 | 38.703 | 0.000 | 1782.000 | 5.000 | 123.419 |
| Protests/Riots (log), ACLED | 1044 | 2.063 | 0.000 | 7.486 | 1.792 | 1.668 |
| ART treatment | 1394 | 0.117 | 0.000 | 0.850 | 0.000 | 0.184 |

**Table A2:**
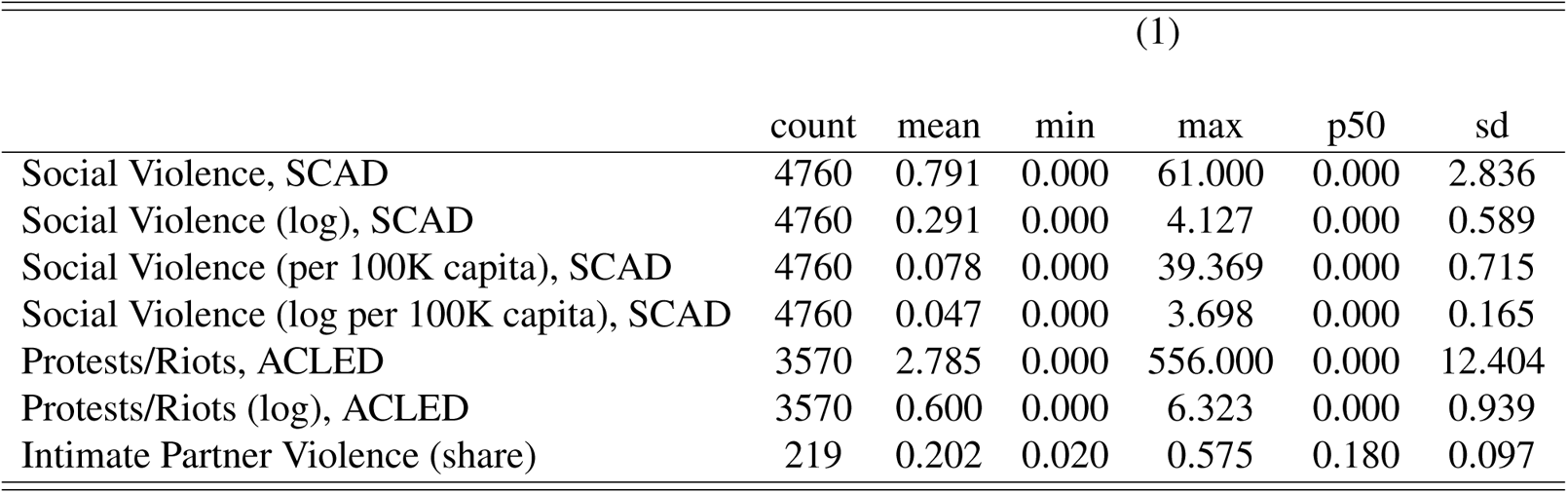
Summary Statistics, Subnational Level

## A.4 Supplementary Results

### A.4.1 2SLS Estimates: First Stage Results

**Table A3:** FIRST STAGE RESULTS: BASELINE SPECIFICATION

|  | ART TREATMENT |  |  |  |  |  |
| --- | --- | --- | --- | --- | --- | --- |
|  | 2SLS | ART TREATMENT (STANDARDISED) |  |  |  |  |
|  | (1) | (2) | (3) | (4) | (5) | (6) |
| $Z_{c,2001} \times ART_{IV,t}$ | 1.328***<br>(0.229) | 1.529***<br>(0.273) | 0.185***<br>(0.031) | 22.071***<br>(3.748) | 0.193***<br>(0.057) | 0.209***<br>(0.063) |
| $Z_{i,2001}$ | $HIV_{c,2001}$ | $HIV_{c,2001}$ | $HIV_{c,2001}$ | $HIV_{c,2001}$ | $HIV_{geo,16K}$ | $HIV_{geo,16K}$ |
| $ART_{IV,t}$ | ART Price | ART Cost | ART Synth. Price | ART Cov | ART Price | ART Synth. Price |
| Observations | 1,394 | 1,394 | 1,394 | 1,394 | 1,366 | 1,366 |
| Clusters | 50 | 50 | 50 | 50 | 49 | 49 |
| Kleibergen-Paap | 33.77 | 31.42 | 35.14 | 34.69 | 11.40 | 11.14 |
| *Country f.e.* | ✓ | ✓ | ✓ | ✓ | ✓ | ✓ |
| *Year f.e.* | ✓ | ✓ | ✓ | ✓ | ✓ | ✓ |
| *Time Trend* | ✓ | ✓ | ✓ | ✓ | ✓ | ✓ |
| *HIV Trend* | ✓ | ✓ | ✓ | ✓ | ✓ | ✓ |
| *Population* | ✓ | ✓ | ✓ | ✓ | ✓ | ✓ |

### A.4.2 2SLS Estimates: Raw vs. Standardized Coefficients

**Table A4:** RAW COEFFICIENTS

| SOCIAL VIOLENCE (LOG EVENTS) - SCAD DATA |  |  |  |  |  |  |  |  |  |
| --- | --- | --- | --- | --- | --- | --- | --- | --- | --- |
|  | OLS | 2SLS | 2SLS - ALTERNATIVE IV CONSTRUCTS |  |  |  |  | ITT |  |
|  | (1) | (2) | (3) | (4) | (5) | (6) | (7) | (8) | (9) |
| ART | -1.393**<br>(0.691) | -4.261**<br>(1.596) | -4.208**<br>(1.591) | -4.114**<br>(1.591) | -4.406**<br>(1.711) | -5.278**<br>(2.551) | -5.414**<br>(2.534) |  |  |
| $Z_{c,2001} \times ART_{IV,t}$ | | | | | | | | -1.283***<br>(0.473) | -0.764***<br>(0.156) |
| $Z_{i,2001}$ | | $HIV_{c,2001}$ | $HIV_{c,2001}$ | $HIV_{c,2001}$ | $HIV_{c,2001}$ | $HIV_{geo,16K}$ | $HIV_{geo,16K}$ | $HIV_{c,2001}$ | $HIV_{r,2001}$ |
| $ART_{IV,t}$ | | ART Price | ART Cost | ART Synth. Price | ART Cov | ART Price | ART Synth. Price | ART Price | ART Price |
| Observations | 1,394 | 1,394 | 1,394 | 1,394 | 1,394 | 1,366 | 1,366 | 1,394 | 4,760 |
| Clusters | 50 | 50 | 50 | 50 | 50 | 49 | 49 | 50 | 170 |
| Adj-R2 | 0.23 | 0.17 | 0.18 | 0.18 | 0.17 | 0.08 | 0.07 | 0.23 | 0.09 |
| Kleibergen-Paap |  | 33.77 | 31.42 | 35.14 | 34.69 | 11.40 | 11.14 |  |  |
| Country f.e. | ✓ | ✓ | ✓ | ✓ | ✓ | ✓ | ✓ | ✓ | × |
| Year f.e. | ✓ | ✓ | ✓ | ✓ | ✓ | ✓ | ✓ | ✓ | ✓ |
| Time Trend | ✓ | ✓ | ✓ | ✓ | ✓ | ✓ | ✓ | ✓ | ✓ |
| HIV Trend | ✓ | ✓ | ✓ | ✓ | ✓ | ✓ | ✓ | ✓ | ✓ |
| Population | ✓ | ✓ | ✓ | ✓ | ✓ | ✓ | ✓ | ✓ | ✓ |
| Region f.e. | × | × | × | × | × | × | × | × | ✓ |

### A.4.3 Robustness of Identification: Additional Results

#### A.4.3.1 Synthetic Control Approach: Alternative Specification Based on HIV Prevalence

**Figure A4:**
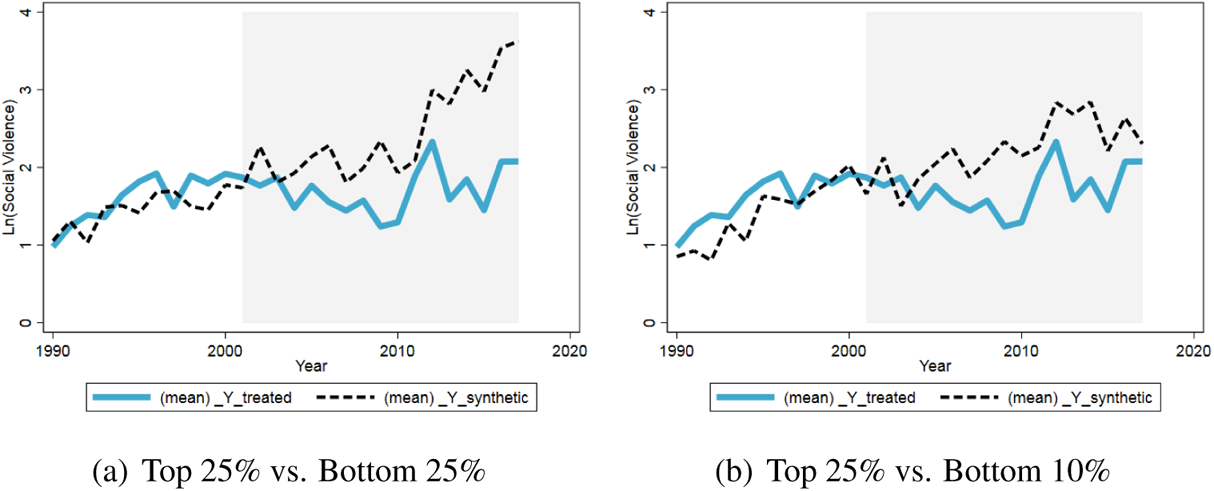
ART EXPANSION AND SOCIAL VIOLENCE: SYNTHETIC CONTROL APPROACH *Note:* Results based on the synthetic control method. For each treated unit, the incidence of social violence is computed under the average treatment and for the synthetic counterfactual. The graph plots averages across all treated units. With the intervention period beginning in 2001, the synthetic control is computed for each treated unit by minimizing the mean squared prediction error (MSPE) relative to the treated units during the pre-intervention period 1990 to 2000. As predictor variables for the construction of the weighted counterfactual of each treated unit, the procedure uses the average log number of conflict events, population and HIV prevalence (all measured between 1990 to 2000), the fraction of the country area within 100 km from the coast, the fraction of desert and of tropical forest, latitude and longitude.

#### A.4.3.2 Identification: Parallel Trends prior to ART Expansion

**Figure A5:**
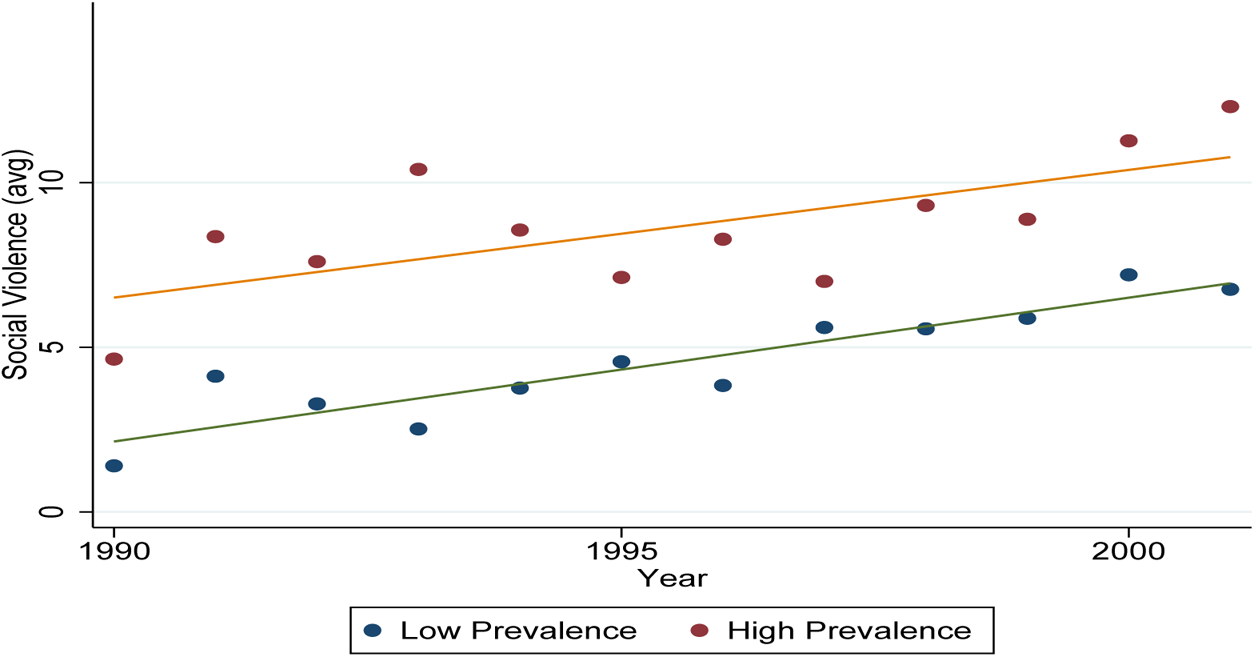
PARALLEL TRENDS: GROUP AVERAGES Note: Scatter plot of the average number of riots for countries with high and low HIV prevalence (threshold: median in 2001). The variable has been demeaned using the average number of events for each group over the period pre-2001 and post-2001, respectively, resembling the re-scaling when including group-specific fixed effects for each period. Linear fit for the two groups over the two periods, pre-2001 and post-2001.

**Figure A6:**
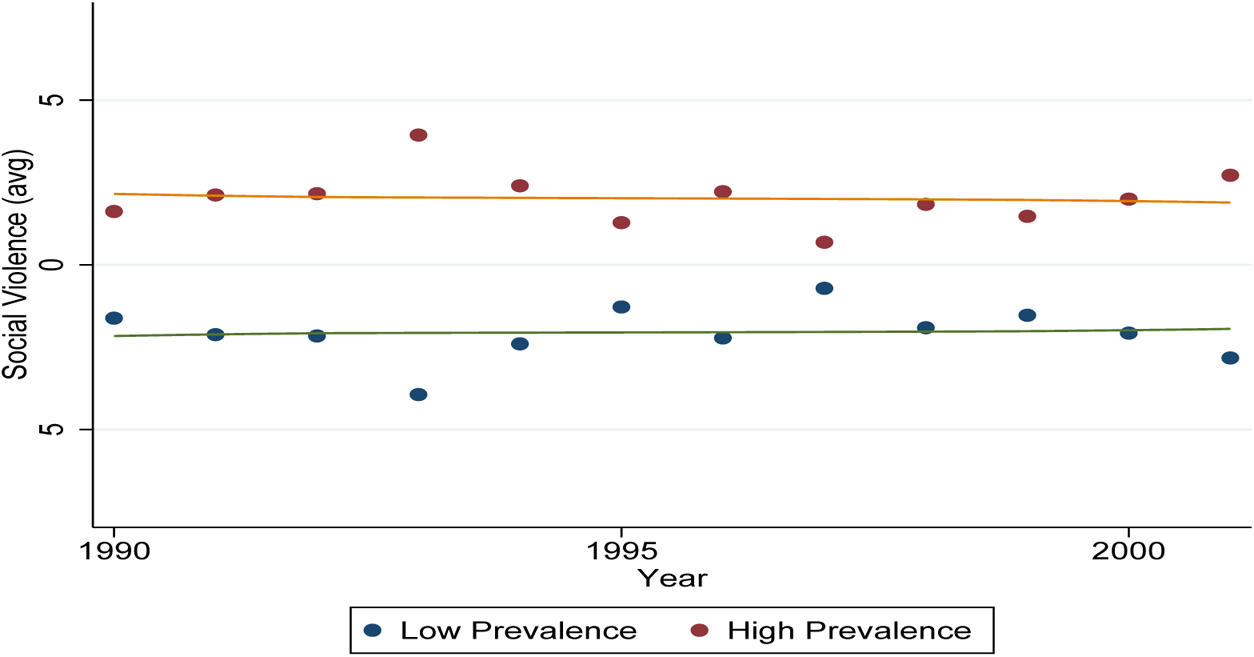
PARALLEL TRENDS: GROUP-YEAR AVERAGES Note: Scatter plot of the average number of riots for countries with high and low HIV prevalence (threshold: median in 2001). Variables have been demeaned using the average number of events for a given year, resembling the re-scaling when including year-fixed effects. Local polynomial fit (bandwidth=2) for the two groups over the two periods, pre-2001 and post-2001.

#### A.4.3.3 Robustness: Alternative Base Years

**Table A5:**
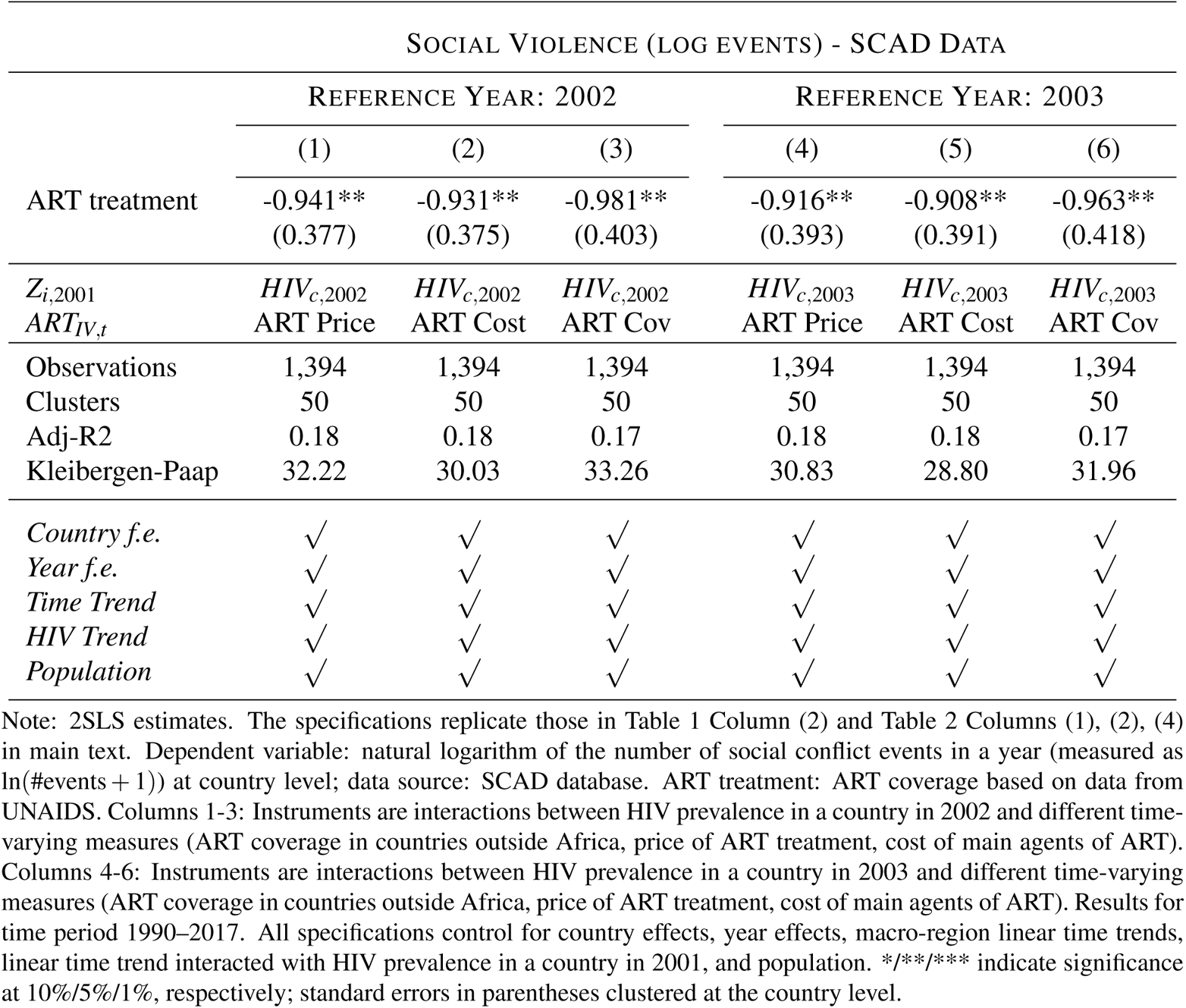
MAIN SPECIFICATION WITH ALTERNATIVE BASE YEARS (SAME TREND)

**Table A6.**
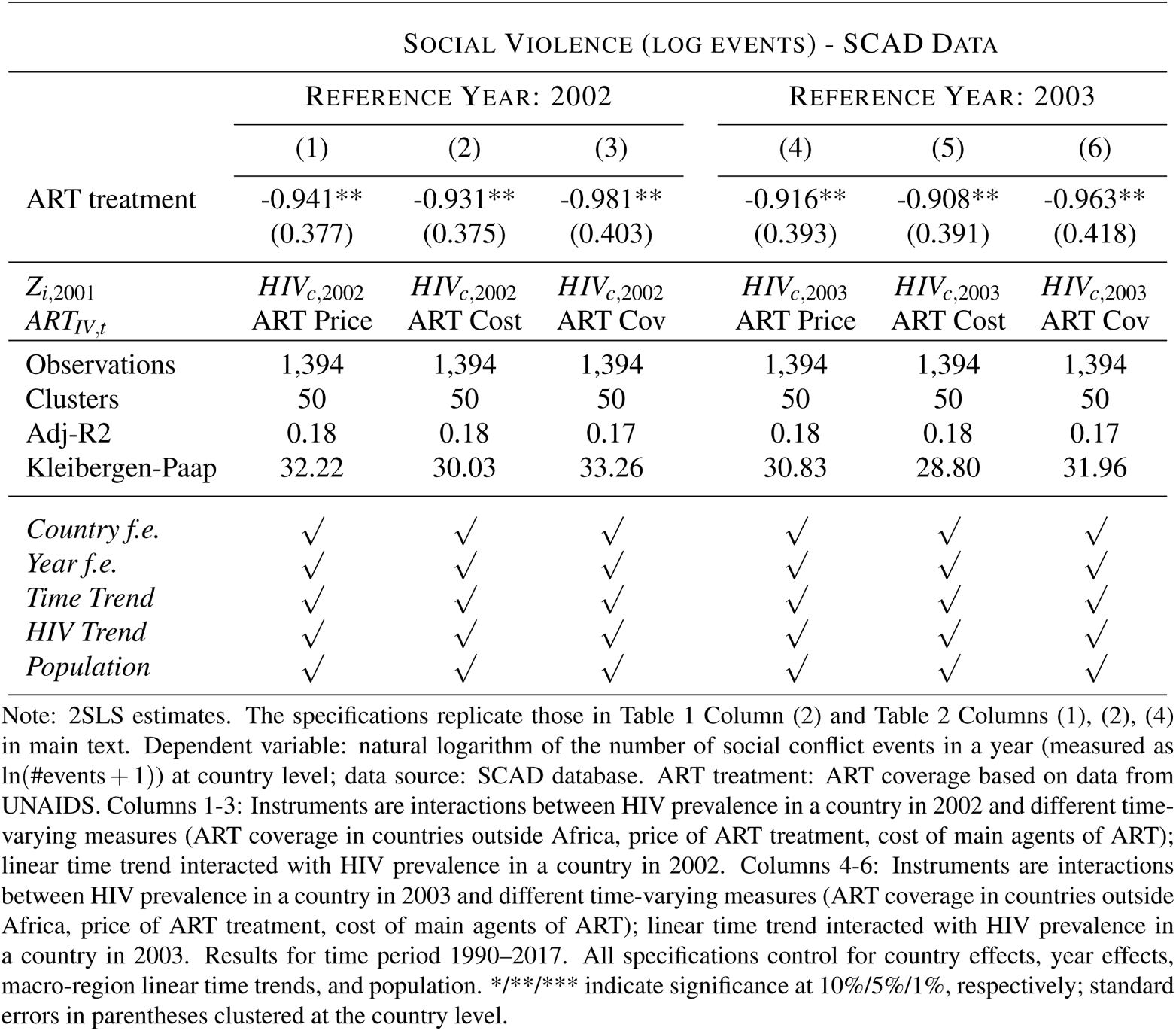
: MAIN SPECIFICATION WITH ALTERNATIVE BASE YEARS AND TRENDS

**Figure A7:**
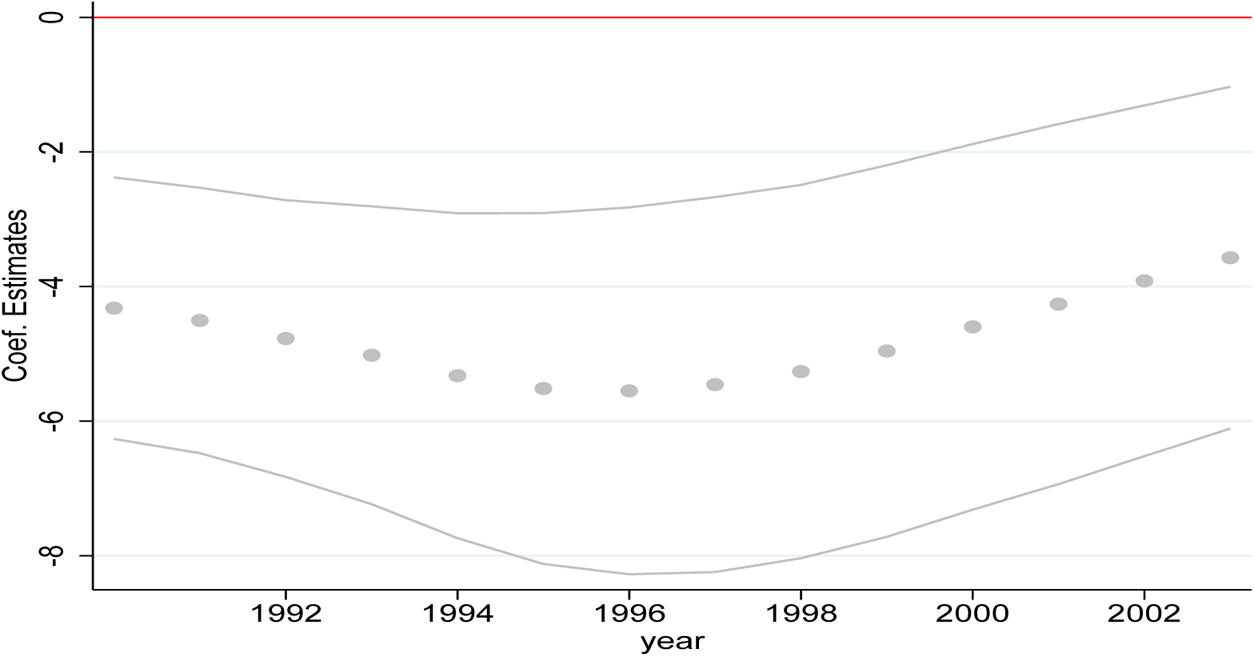
COEFFICIENT PLOTS FOR DIFFERENT BASE YEARS *T* IN *HIV_c,T_* Note: The figure plots the coefficient of interest (dots) from the 2SLS model, obtained for different reference years *T* in the construction of the instrument *HIV_c,T_ · ART_IV,t_* (Instrument: ART Coverage outside Africa). Lines show the corresponding 90-% confidence interval. The specification is the analogue of Table 1 Column (2) in the paper.

**Figure A8:**
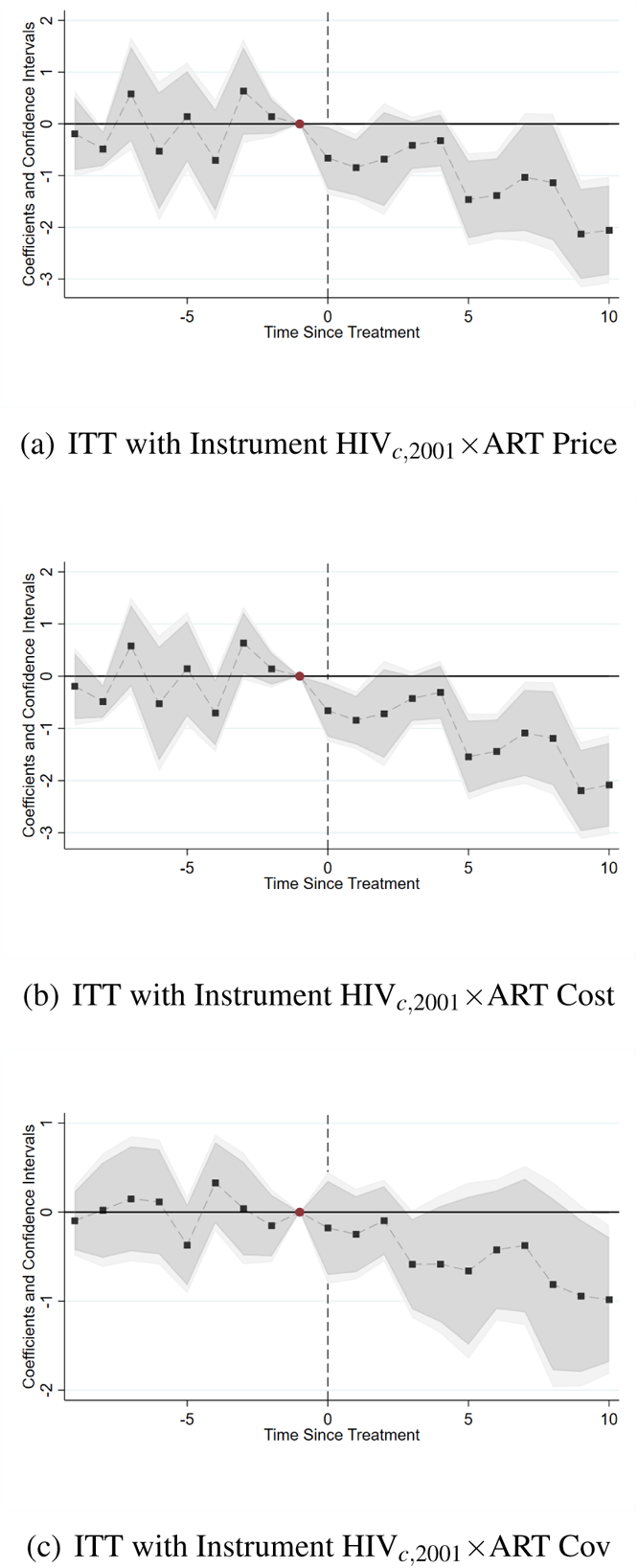
REDUCED FORM ESTIMATES – EVENT STUDY PLOTS Note: The figure plots event study graphs for the coefficient of interest from the ITT model. The empirical specification is as in Table 1 Column (2) of the paper. The estimation is conducted using the routine devised by Chaisemartin and Haultfoeuille (2020). Panel (a): Instruments are HIV*_c,_*_2001_*×*ART Price as in Table 1 Column (2). Panel (b): Instruments are HIV*_c,_*_2001_*×*ART Cost as in Table 2 Column (2). Panel (c): Instruments are HIV*_c,_*_2001_*×*ART Cost as in Table 2 Column (4). Dark shades show the corresponding 90-% confidence interval, light shades the corresponding 95% confidence interval.

#### A.4.3.4 Robustness of Instrument: Alternative Price Data and Production Costs Data

**Table A7:** ALTERNATIVE PRICE AND PRODUCTION COSTS DATA

| SOCIAL VIOLENCE (LOG EVENTS) - SCAD DATA |  |  |  |  |  |  |  |
| --- | --- | --- | --- | --- | --- | --- | --- |
|  | OLS | COUNTRY LEVEL<br>2SLS |  |  | SUBNATIONAL LEVEL<br>ITT |  |  |
|  | (1) | (2) | (3) | (4) | (5) | (6) | (7) |
| ART treatment | -0.316**<br>(0.157) | -0.910**<br>(0.363) | -0.953**<br>(0.368) | -0.999**<br>(0.388) |  |  |  |
| $Z_{c,2001} \times ART_{IV,t}$ | | | | | -0.182***<br>(0.036) | -0.179***<br>(0.035) | -0.215***<br>(0.046) |
| $Z_{i,2001}$ | | $HIV_{c,2001}$ | $HIV_{c,2001}$ | $HIV_{c,2001}$ | $HIV_{c,2001}$ | $HIV_{c,2001}$ | $HIV_{c,2001}$ |
| $ART_{IV,t}$ | | ART Price | ART Cost | ART Cov | ART Price | ART Cost | ART Cov |
| Observations | 1,394 | 1,394 | 1,394 | 1,394 | 4,760 | 4,760 | 4,760 |
| Clusters | 50 | 50 | 50 | 50 | 170 | 170 | 170 |
| Adj-R2 | 0.23 | 0.18 | 0.18 | 0.17 | 0.09 | 0.09 | 0.09 |
| Kleibergen-Paap |  | 40.97 | 38.66 | 34.69 |  |  |  |
| Country f.e. | ✓ | ✓ | ✓ | ✓ | × | × | × |
| Region f.e. | n.a. | n.a. | n.a. | n.a. | ✓ | ✓ | ✓ |
| Year f.e. | ✓ | ✓ | ✓ | ✓ | ✓ | ✓ | ✓ |
| Time Trend | ✓ | ✓ | ✓ | ✓ | ✓ | ✓ | ✓ |
| HIV Trend | ✓ | ✓ | ✓ | ✓ | ✓ | ✓ | ✓ |
| Population | ✓ | ✓ | ✓ | ✓ | ✓ | ✓ | ✓ |

#### A.4.3.5 Robustness of Instrument: Alternative Specifications for Treatment Scope and Intensity

**Table A8:** ALTERNATIVE SPECIFICATIONS OF INSTRUMENT:

| Panel A |  |  |  |  |  |
| --- | --- | --- | --- | --- | --- |
| SOCIAL VIOLENCE (LOG EVENTS) - SCAD DATA |  |  |  |  |  |
|  | (1) | (2) | (3) | (4) | (5) |
| ART treatment | -0.966**<br>(0.362) | -0.978*<br>(0.506) | -1.361**<br>(0.613) | -1.233**<br>(0.587) | -1.197**<br>(0.578) |
| $Z_{i,2001}$<br>$ART_{IV,t}$ | $HIV_{c,2001}$<br>ART Price | $HIV_{geo}$<br>ART Price | $HIV_{geo,10K}$<br>ART Price | $HIV_{geo,16K}$<br>ART Price | $HIV_{geo,20K}$<br>ART Price |
| Observations | 1,394 | 1,366 | 1,366 | 1,366 | 1,366 |
| Countries | 50 | 49 | 49 | 49 | 49 |
| Adj-R2 | 0.17 | 0.14 | 0.03 | 0.07 | 0.08 |
| Kleibergen-Paap | 33.77 | 7.24 | 11.61 | 11.76 | 11.40 |
| Panel B |  |  |  |  |  |
| SOCIAL VIOLENCE (LOG EVENTS) - SCAD DATA |  |  |  |  |  |
|  | (1) | (2) | (3) | (4) | (5) |
| ART | -0.966**<br>(0.362) | -0.938**<br>(0.366) | -0.933**<br>(0.361) | -0.922**<br>(0.356) | -1.227**<br>(0.575) |
| $Z_{i,2001}$<br>$ART_{IV,t}$ | $HIV_{c,2001}$<br>ART Price | $HIV_{c,2001}$<br>Synth. Price (15%) | $HIV_{c,2001}$<br>Synth. Price (20%) | $HIV_{c,2001}$<br>Synth. Price (25%) | $HIV_{geo,20K}$<br>Synth. Price (25%) |
| Mean |  |  |  |  |  |
| Observations | 1,394 | 1,394 | 1,394 | 1,394 | 1,366 |
| Regions | 50 | 50 | 50 | 50 | 49 |
| R2 | 0.22 | 0.23 | 0.23 | 0.23 | 0.13 |
| *Region f.e.* | ✓ | ✓ | ✓ | ✓ | ✓ |
| *Year f.e.* | ✓ | ✓ | ✓ | ✓ | ✓ |
| *Time Trend* | ✓ | ✓ | ✓ | ✓ | ✓ |
| *HIV Trend* | ✓ | ✓ | ✓ | ✓ | ✓ |
| *Population* | ✓ | ✓ | ✓ | ✓ | ✓ |

Note: 2SLS estimates. Dependent variable: natural logarithm of the number of social violence events in a year (measured as $\ln(\#events + 1)$ ) at country level; data source: SCAD database. ART coverage; Data source is UNAIDS. Results for time period 1990–2017. All specifications control for country effects, year effects, macro-region linear time trends, linear time trend interacted with HIV prevalence in a country in 2001, and population. \*/\*\*/\* indicate significance at 10%/5%/1%, respectively, standard errors in parentheses clustered at the country level.

#### A.4.3.6 Falsification of Instrument: Malaria

**Table A9:** FALSIFICATION OF INSTRUMENT: MALARIA PREVALENCE AND TREATMENT

|  | ART TREATMENT |  |  |  |
| --- | --- | --- | --- | --- |
|  | (1) | (2) | (3) | (4) |
| $X_{i,2001} \times ART_{IV,t}$ | 0.004<br>(0.003) | -0.046<br>(0.071) | -0.045<br>(0.070) | -0.049<br>(0.082) |
| $X_{i,2001}$ | $HIV_{c,2001}$ | $Malaria_{c,2001}$ | $Malaria_{c,2001}$ | $Malaria_{c,2001}$ |
| $ART_{IV,t}$ | Malaria Price | ART Price | ART Cost | ART Cov |
| Observations | 1,296 | 1,338 | 1,338 | 1,338 |
| Clusters | 50 | 48 | 48 | 48 |
| Kleibergen-Paap | 2.00 | 0.42 | 0.42 | 0.35 |
| *Country f.e* | ✓ | ✓ | ✓ | ✓ |
| *Year f.e* | ✓ | ✓ | ✓ | ✓ |
| *Time Trend* | ✓ | ✓ | ✓ | ✓ |
| *HIV Trend* | ✓ | ✓ | ✓ | ✓ |
| *Population* | ✓ | ✓ | ✓ | ✓ |

Note: Coefficient estimates (OLS). Dependent variable: ART coverage based on data from UNAIDS. Results for time period 1990–2017. All specifications control for country effects, year effects, macro-region linear time trends, linear time trend interacted with HIV prevalence in a country in 2001, and population. \*/\*\*/\*\* indicate significance at 10%/5%/1%, respectively, standard errors in parentheses clustered at the country level.

#### A.4.3.7 Placebo Instrument: Institutional Quality

**Table A10:** PLACEBO 1: REPLACING *Z_c,_*_2001_ *· ART_IV,t_* BY *X_c,_*_2001_ *· ART_IV,t_*–FIRST AND SECOND STAGE

| Panel A: First Stage |  | ART TREATMENT |  |  |  |  |  |
| --- | --- | --- | --- | --- | --- | --- | --- |
| $X_{c,2001}$ | $HIV_{c,2001}$<br>(1) | $Polity_{c,2001}$<br>(2) | $Polity2_{c,2001}$<br>(3) | $Democracyscore_{c,2001}$<br>(4) | $GlobalFund_{c,2001}$<br>(5) | $GDPpc_{c,2001}$<br>(6) | $SCADRiots_{c,2001}$<br>(7) |
| Instrument | 0.169***<br>(0.029) | -0.011<br>(0.047) | 0.045<br>(0.060) | -0.013<br>(0.046) | 0.153***<br>(0.050) | -0.001<br>(0.052) | 0.062<br>(0.078) |
| Kleibergen-Paap | 33.77 | 0.05 | 0.56 | 0.08 | 9.48 | 0.00 | 0.64 |
| Observations | 1,394 | 1,338 | 1,338 | 1,338 | 1,394 | 1,310 | 1,394 |
| Clusters | 50 | 48 | 48 | 48 | 50 | 47 | 50 |
| Panel B: Second Stage |  | SOCIAL VIOLENCE (LOG EVENTS) - SCAD DATA |  |  |  |  |  |
| $X_{c,2001}$ | $HIV_{c,2001}$<br>(1) | $Polity_{c,2001}$<br>(2) | $Polity2_{c,2001}$<br>(3) | $Democracyscore_{c,2001}$<br>(4) | $GlobalFund_{c,2001}$<br>(5) | $GDPpc_{c,2001}$<br>(6) | $SCADRiots_{c,2001}$<br>(7) |
| ART | -0.966**<br>(0.362) | -6.609<br>(32.467) | -0.755<br>(2.255) | -5.458<br>(22.679) | -0.941<br>(0.605) | -91.176<br>(6178.312) | -2.188<br>(2.516) |
| Observations | 1,394 | 1,338 | 1,338 | 1,338 | 1,394 | 1,310 | 1,394 |
| Clusters | 50 | 48 | 48 | 48 | 50 | 47 | 50 |
| Country f.e. | ✓ | ✓ | ✓ | ✓ | ✓ | ✓ | ✓ |
| Year f.e. | ✓ | ✓ | ✓ | ✓ | ✓ | ✓ | ✓ |
| Time Trend | ✓ | ✓ | ✓ | ✓ | ✓ | ✓ | ✓ |
| Share Trend | ✓ | ✓ | ✓ | ✓ | ✓ | ✓ | ✓ |
| Population | ✓ | ✓ | ✓ | ✓ | ✓ | ✓ | ✓ |

**Table A11:** PLACEBO 2: REPLACING *ZV_c,_*_2001_ *· ART_IV,t_* BY *X_c,_*_2001_ *· ART_IV,t_*–FIRST AND SECOND STAGE

| Panel A: First Stage |  | SOCIAL VIOLENCE (LOG EVENTS) - SCAD DATA |  |  |  |  |  |
| --- | --- | --- | --- | --- | --- | --- | --- |
| $X_{c,2001}$ | $HIV_{2001,c,2001}$<br>(8) | $Voice\&\;Accountability_{c,2001}$<br>(9) | $Political\;Stability_{c,2001}$<br>(10) | $Gov\;Effectiveness_{c,2001}$<br>(11) | $Regulatory\;Quality_{c,2001}$<br>(12) | $Rule\;of\;Law_{c,2001}$<br>(13) | $Corruption_{c,2001}$<br>(14) |
| Instrument | 0.169***<br>(0.029) | 0.100<br>(0.074) | 0.039<br>(0.062) | 0.098<br>(0.085) | 0.113<br>(0.081) | 0.135*<br>(0.070) | 0.091<br>(0.083) |
| Kleibergen-Paap | 33.77 | 1.85 | 0.40 | 1.35 | 1.94 | 3.70 | 1.20 |
| Observations | 1,394 | 1,366 | 1,366 | 1,366 | 1,366 | 1,366 | 1,366 |
| Clusters | 50 | 49 | 49 | 49 | 49 | 49 | 49 |
| Panel B: Second Stage |  | SOCIAL VIOLENCE (LOG EVENTS) - SCAD DATA |  |  |  |  |  |
| $X_{c,2001}$ | $HIV_{2001,c,2001}$<br>(8) | $Voice\&\;Accountability_{c,2001}$<br>(9) | $Political\;Stability_{c,2001}$<br>(10) | $Gov\;Effectiveness_{c,2001}$<br>(11) | $Regulatory\;Quality_{c,2001}$<br>(12) | $Rule\;of\;Law_{c,2001}$<br>(13) | $Corruption_{c,2001}$<br>(14) |
| ART | -0.966**<br>(0.362) | 0.217<br>(1.452) | 2.135<br>(4.379) | 1.316<br>(2.000) | 1.300<br>(1.614) | 1.274<br>(1.301) | 1.251<br>(2.144) |
| Observations | 1,394 | 1,366 | 1,366 | 1,366 | 1,366 | 1,366 | 1,366 |
| Clusters | 50 | 49 | 49 | 49 | 49 | 49 | 49 |
| Country f.e. | ✓ | ✓ | ✓ | ✓ | ✓ | ✓ | ✓ |
| Year f.e. | ✓ | ✓ | ✓ | ✓ | ✓ | ✓ | ✓ |
| Time Trend | ✓ | ✓ | ✓ | ✓ | ✓ | ✓ | ✓ |
| Share Trend | ✓ | ✓ | ✓ | ✓ | ✓ | ✓ | ✓ |
| Population | ✓ | ✓ | ✓ | ✓ | ✓ | ✓ | ✓ |

#### A.4.3.8 Overidentification: Controlling for Institutional Quality Interactions

**Table A12:**
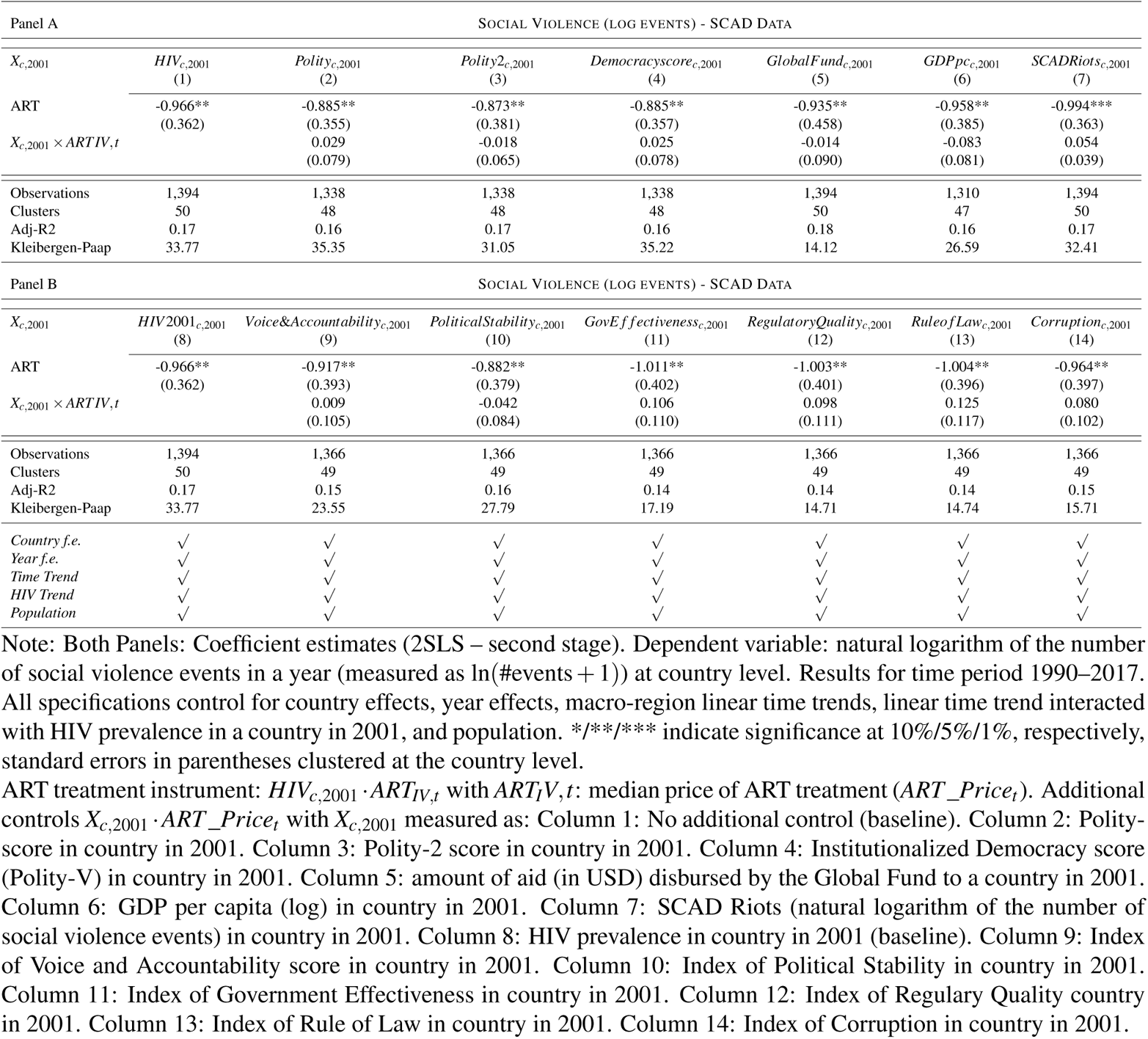
OVERIDENTIFICATION 1: CONTROLLING FOR PLACEBO-IV *X_c,_*_2001_ *· ART_IV,t_* –SECOND STAGE RESULTS

**Table A13:** OVERIDENTIFICATION 2: CONTROLLING FOR PLACEBO-IV *X_c,_*_2001_ *· ART_IV,t_* –ITT RESULTS

| Panel A |  |  |  |  |  |  |  |
| --- | --- | --- | --- | --- | --- | --- | --- |
| SOCIAL VIOLENCE (LOG EVENTS) - SCAD DATA |  |  |  |  |  |  |  |
| $X_{c,2001}$ | $HIV_{c,2001}$<br>(1) | $Polity_{c,2001}$<br>(2) | $Polity2_{c,2001}$<br>(3) | $Democracy_{c,2001}$<br>(4) | $GlobalFund_{c,2001}$<br>(5) | $GDPpc_{c,2001}$<br>(6) | $SCADRiot_{c,2001}$<br>(7) |
| ART treatment | -0.163***<br>(0.060) | -0.150**<br>(0.059) | -0.145**<br>(0.064) | -0.150**<br>(0.059) | -0.139**<br>(0.064) | -0.151**<br>(0.061) | -0.167***<br>(0.061) |
| $X_{c,2001} \times ART_{IV,t}$ | | 0.014<br>(0.056) | -0.043<br>(0.058) | 0.013<br>(0.054) | -0.061<br>(0.056) | -0.101*<br>(0.051) | 0.054<br>(0.059) |
| Observations | 1,394 | 1,338 | 1,338 | 1,338 | 1,394 | 1,310 | 1,394 |
| Clusters | 50 | 48 | 48 | 48 | 50 | 47 | 50 |
| Adj-R2 | 0.23 | 0.22 | 0.22 | 0.22 | 0.23 | 0.23 | 0.23 |
| Panel B |  |  |  |  |  |  |  |
| SOCIAL VIOLENCE (LOG EVENTS) - SCAD DATA |  |  |  |  |  |  |  |
| $X_{c,2001}$ | $HIV2001_{c,2001}$<br>(8) | $Voice\&Accountability_{c,2001}$<br>(9) | $PoliticalStability_{c,2001}$<br>(10) | $GovEffectiveness_{c,2001}$<br>(11) | $RegulatoryQuality_{c,2001}$<br>(12) | $RuleofLaw_{c,2001}$<br>(13) | $Corruption_{c,2001}$<br>(14) |
| ART treatment | -0.163***<br>(0.060) | -0.143**<br>(0.066) | -0.143**<br>(0.063) | -0.150**<br>(0.062) | -0.148**<br>(0.061) | -0.147**<br>(0.063) | -0.146**<br>(0.062) |
| $X_{c,2001} \times ART_{IV,t}$ | | -0.075<br>(0.086) | -0.066<br>(0.074) | -0.003<br>(0.089) | -0.021<br>(0.065) | -0.033<br>(0.082) | -0.047<br>(0.085) |
| Observations | 1,394 | 1,366 | 1,366 | 1,366 | 1,366 | 1,366 | 1,366 |
| Clusters | 50 | 49 | 49 | 49 | 49 | 49 | 49 |
| Adj-R2 | 0.23 | 0.22 | 0.22 | 0.22 | 0.22 | 0.22 | 0.22 |
| Country f.e. | ✓ | ✓ | ✓ | ✓ | ✓ | ✓ | ✓ |
| Year f.e. | ✓ | ✓ | ✓ | ✓ | ✓ | ✓ | ✓ |
| Time Trend | ✓ | ✓ | ✓ | ✓ | ✓ | ✓ | ✓ |
| HIV Trend | ✓ | ✓ | ✓ | ✓ | ✓ | ✓ | ✓ |
| Population | ✓ | ✓ | ✓ | ✓ | ✓ | ✓ | ✓ |

Note: Both Panels: Coefficient estimates (intention-to-treat), OLS. Dependent variable: natural logarithm of the number of social violence events in a year (measured as $\ln(\#events + 1)$ ) at country level. Results for time period 1990–2017. All specifications control for country effects, year effects, macro-region linear time trends, linear time trend interacted with HIV prevalence in a country in 2001, and population. \*/\*\*/\*\* indicate significance at 10%/5%/1%, respectively, standard errors in parentheses clustered at the country level.

#### A.4.3.9 Relaxing the Exclusion Restriction of Strict Exogeneity

**Figure A9:**
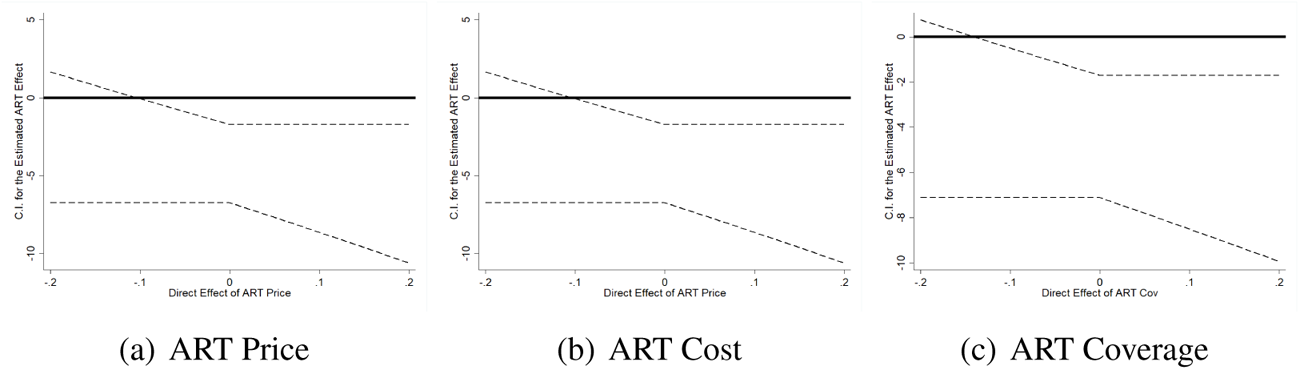
EFFECT OF INTEREST UNDER PARTIAL EXOGENEITY Note: Coefficient estimates (2SLS). Dependent variable: natural logarithm of the number of social conflict events in a year (measured as ln(#events + 1)) at country level; data source: SCAD database. ART treatment: ART coverage based on data from UNAIDS. Estimates for different instruments *HIV_c,_*_2001_ *· ART_IV,t_* if the instrument has a direct effect (depicted on the horizontal axis) on the outcome (Conley, Hansen and Rossi 2012).

### A.4.4 Robustness: Samples and Specification

#### A.4.4.1 Eliminating Single Countries

**Figure A10:**
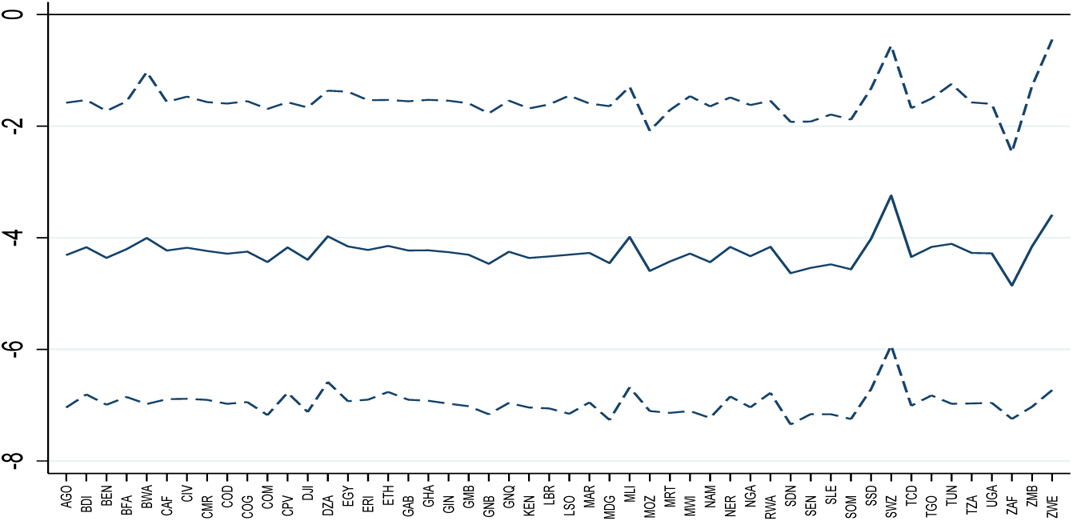
SENSITIVITY OF RESULTS WHEN EXCLUDING COUNTRIES – 2SLS RESULTS Note: The plot shows the coefficient of interest (and the corresponding 90% confidence band) obtained with the 2SLS estimation (OLS/2SLS–Stage 2) with instruments HIV prevalence in country *c* in year 2001 and ART Coverage outside Africa in year *t*, when the corresponding country shown at the bottom of the figure is excluded from the estimation sample.

**Figure A11:**
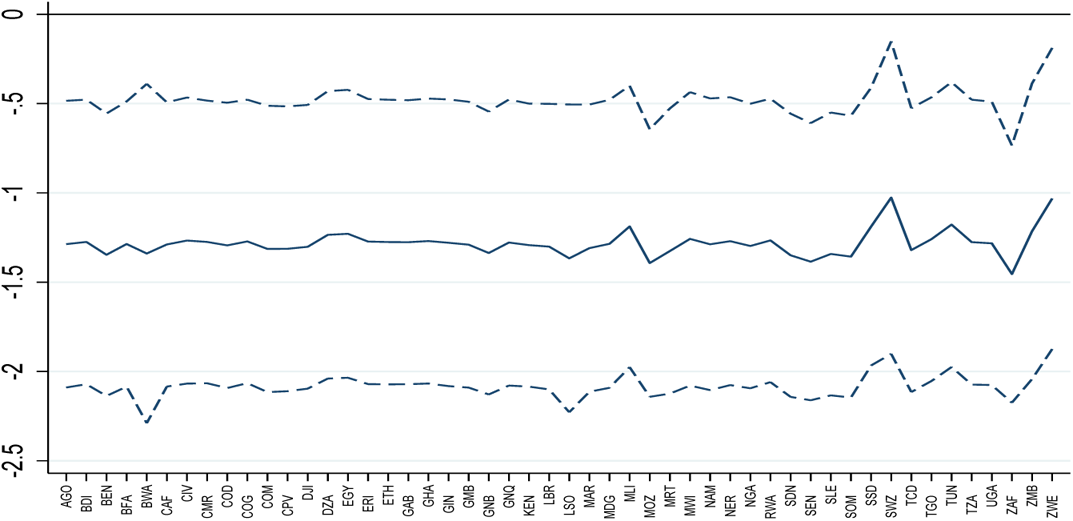
SENSITIVITY OF RESULTS WHEN EXCLUDING COUNTRIES – ITT RESUTS Note: The plot shows the coefficient of interest (and the corresponding 90% confidence band) obtained with the ITT estimation (ITT), iwth instruments HIV prevalence in country *c* in year 2001, ART Coverage outside Africa in year *t*, when the corresponding country shown at the bottom of the figure is excluded from the estimation sample.

#### A.4.4.2 Accounting for Non-Linear Trends in African Regions

**Table A14:**
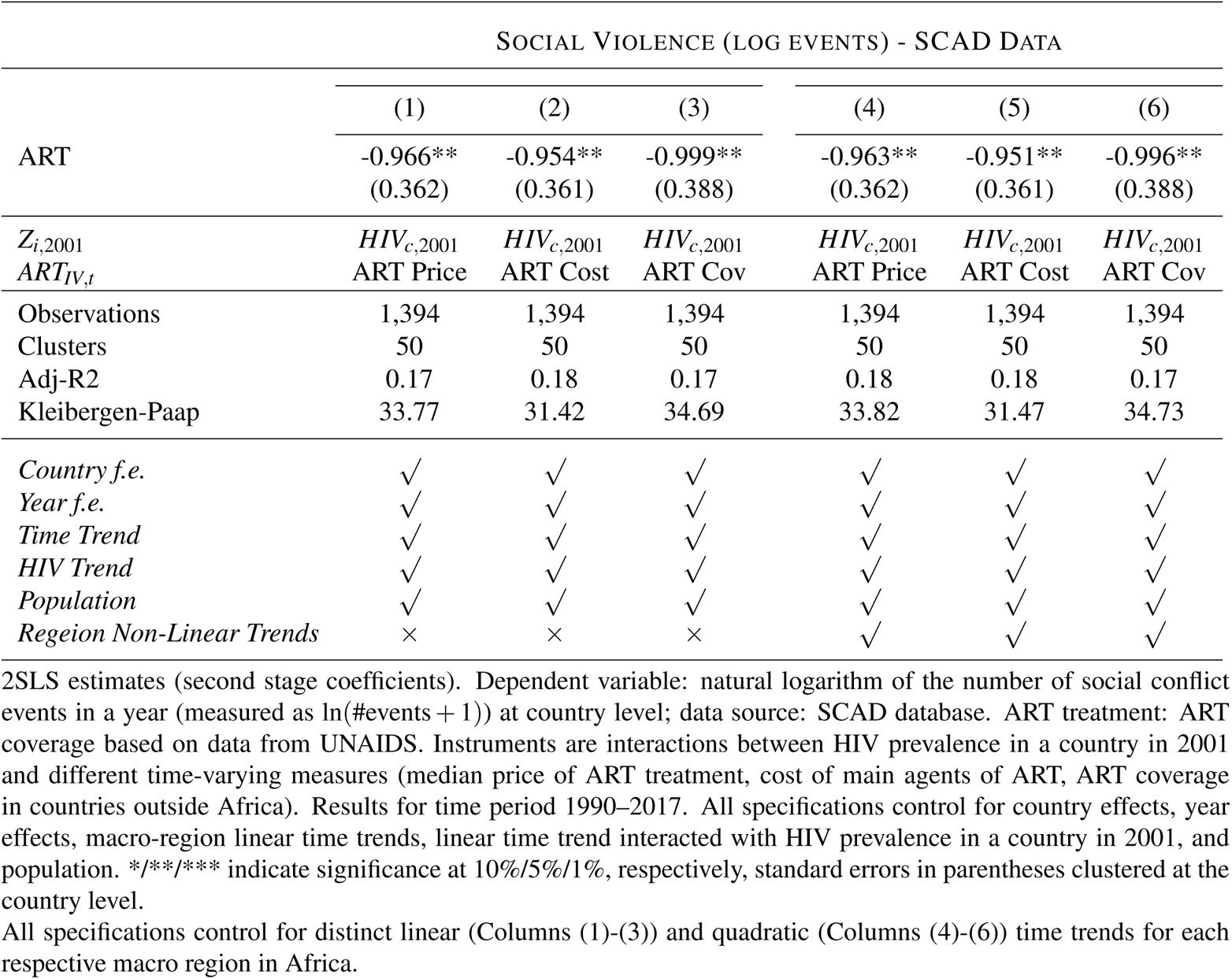
ACCOUNTING FOR NON-LINEAR REGION TRENDS

#### A.4.4.3 Alternative Samples

**Table A15:**
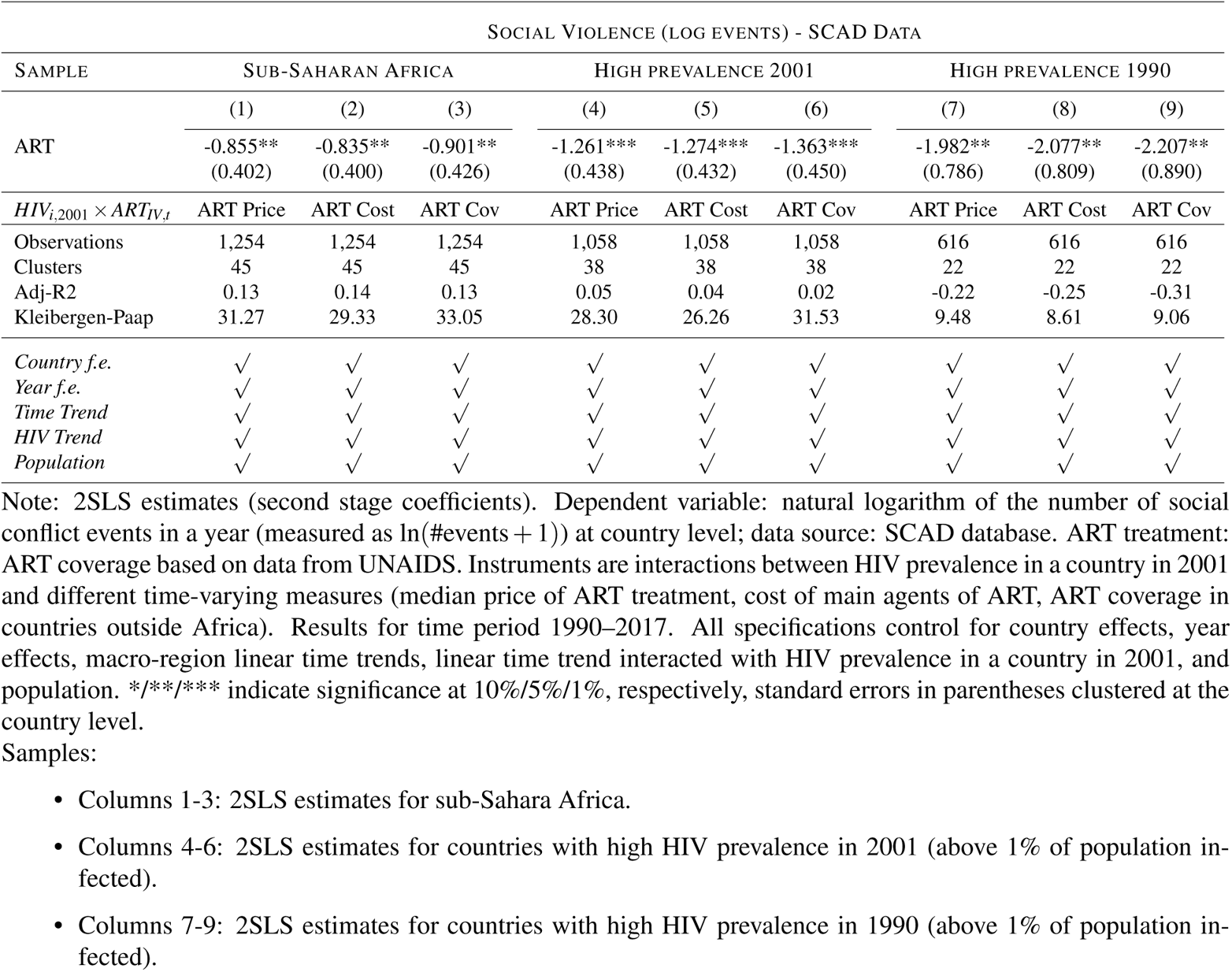
ALTERNATIVE SAMPLES

#### A.4.4.4 Health Aid, Income and Democracy

**Table A16:** AID HEALTH, INCOME AND DEMOCRACY

| SOCIAL VIOLENCE (LOG EVENTS) - SCAD DATA |  |  |  |  |  |  |  |  |  |  |  |  |
| --- | --- | --- | --- | --- | --- | --- | --- | --- | --- | --- | --- | --- |
|  | (1) | (2) | (3) | (4) | (5) | (6) | (7) | (8) | (9) | (10) | (11) | (12) |
| ART | -0.937**<br>(0.368) | -0.928**<br>(0.367) | -0.964**<br>(0.399) | -0.997**<br>(0.377) | -0.978**<br>(0.376) | -1.035**<br>(0.408) | -0.897**<br>(0.360) | -0.881**<br>(0.357) | -0.932**<br>(0.386) | -0.936**<br>(0.388) | -0.916**<br>(0.386) | -0.970**<br>(0.424) |
| $HIV_{i,2001} \times ART_{IV,t}$ | ART Price | ART Cost | ART Cov | ART Price | ART Cost | ART Cov | ART Price | ART Cost | ART Cov | ART Price | ART Cost | ART Cov |
| Observations | 1,394 | 1,394 | 1,394 | 1,299 | 1,299 | 1,299 | 1,340 | 1,340 | 1,340 | 1,268 | 1,268 | 1,268 |
| Clusters | 50 | 50 | 50 | 48 | 48 | 48 | 50 | 50 | 50 | 48 | 48 | 48 |
| Adj-R2 | 0.18 | 0.18 | 0.18 | 0.15 | 0.16 | 0.15 | 0.18 | 0.18 | 0.17 | 0.18 | 0.19 | 0.18 |
| Kleibergen-Paap | 36.87 | 34.20 | 36.16 | 30.65 | 28.54 | 31.22 | 33.32 | 30.83 | 34.40 | 32.04 | 29.56 | 30.44 |
| Country f.e | ✓ | ✓ | ✓ | ✓ | ✓ | ✓ | ✓ | ✓ | ✓ | ✓ | ✓ | ✓ |
| Year f.e | ✓ | ✓ | ✓ | ✓ | ✓ | ✓ | ✓ | ✓ | ✓ | ✓ | ✓ | ✓ |
| Time Trend | ✓ | ✓ | ✓ | ✓ | ✓ | ✓ | ✓ | ✓ | ✓ | ✓ | ✓ | ✓ |
| HIV Trend | ✓ | ✓ | ✓ | ✓ | ✓ | ✓ | ✓ | ✓ | ✓ | ✓ | ✓ | ✓ |
| Population | ✓ | ✓ | ✓ | ✓ | ✓ | ✓ | ✓ | ✓ | ✓ | ✓ | ✓ | ✓ |
| Health Aid | ✓ | ✓ | ✓ | × | × | × | ✓ | × | × | ✓ | ✓ | ✓ |
| Income | × | × | × | ✓ | ✓ | ✓ | × | × | × | ✓ | ✓ | ✓ |
| Democracy | × | × | × | × | × | × | ✓ | ✓ | ✓ | ✓ | ✓ | ✓ |

Note: 2SLS estimates (second stage coefficients). Dependent variable: natural logarithm of the number of social conflict events in a year (measured as $\ln(\#events + 1)$ ) at country level; data source: SCAD database. ART treatment: ART coverage based on data from UNAIDS. Instruments are interactions between HIV prevalence in a country in 2001 and different time-varying measures (median price of ART treatment, cost of main agents of ART, ART coverage in countries outside Africa). Results for time period 1990–2017. All specifications control for country effects, year effects, macro-region linear time trends, linear time trend interacted with HIV prevalence in a country in 2001, and population. \*/\*\*/\*\* indicate significance at 10%/5%/1%, respectively, standard errors in parentheses clustered at the country level.
Additional control variables:
- Columns 1-3: Amount of aid (in USD) disbursed by the Global Fund to a country in a given year. - Columns 4-6: GDP per capita. - Columns 7-9: Democracy (Institutionalized Democracy Index, Polity V). - Columns 10-12: Amount of aid (in USD) disbursed by the Global Fund to a country in a given year, GDP per capita, Democracy (Institutionalized Democracy Index, Polity V).

### A.4.5 Robustness: Alternative Coding for ART and Social Violence

#### A.4.5.1 ART Coverage: Alternative Measure

**Table A17:** ART COVERAGE: ALTERNATIVE MEASURE

| SOCIAL VIOLENCE (LOG EVENTS) - SCAD DATA |  |  |  |  |  |  |  |
| --- | --- | --- | --- | --- | --- | --- | --- |
|  | OLS | COUNTRY LEVEL<br>2SLS |  |  | SUBNATIONAL LEVEL<br>ITT |  |  |
|  | (1) | (2) | (3) | (4) | (5) | (6) | (7) |
| ART | -0.153***<br>(0.054) | -0.177***<br>(0.065) | -0.174***<br>(0.064) | -0.178***<br>(0.064) |  |  |  |
| $Z_{c,2001} \times ART_{IV,t}$ | | | | | -0.166***<br>(0.034) | -0.157***<br>(0.032) | -0.215***<br>(0.046) |
| $HIV_{i,2001} \times ART_{IV,t}$ | | ART Price | ART Cost | ART Cov | ART Price | ART Cost | ART Cov |
| Observations | 1,394 | 1,394 | 1,394 | 1,394 | 4,760 | 4,760 | 4,760 |
| Clusters | 50 | 50 | 50 | 50 | 170 | 170 | 170 |
| Adj-R2 | 0.23 | 0.23 | 0.23 | 0.23 | 0.09 | 0.09 | 0.09 |
| Kleibergen-Paap |  | 134.17 | 126.83 | 112.04 |  |  |  |
| Country f.e. | ✓ | ✓ | ✓ | ✓ | × | × | × |
| Region f.e. | n.a. | n.a. | n.a. | n.a. | ✓ | ✓ | ✓ |
| Year f.e. | ✓ | ✓ | ✓ | ✓ | ✓ | ✓ | ✓ |
| Time Trend | ✓ | ✓ | ✓ | ✓ | ✓ | ✓ | ✓ |
| HIV Trend | ✓ | ✓ | ✓ | ✓ | ✓ | ✓ | ✓ |
| Population | ✓ | ✓ | ✓ | ✓ | ✓ | ✓ | ✓ |

#### A.4.5.2 Social Violence, SCAD (Log Events Per Population)

**Table A18:**
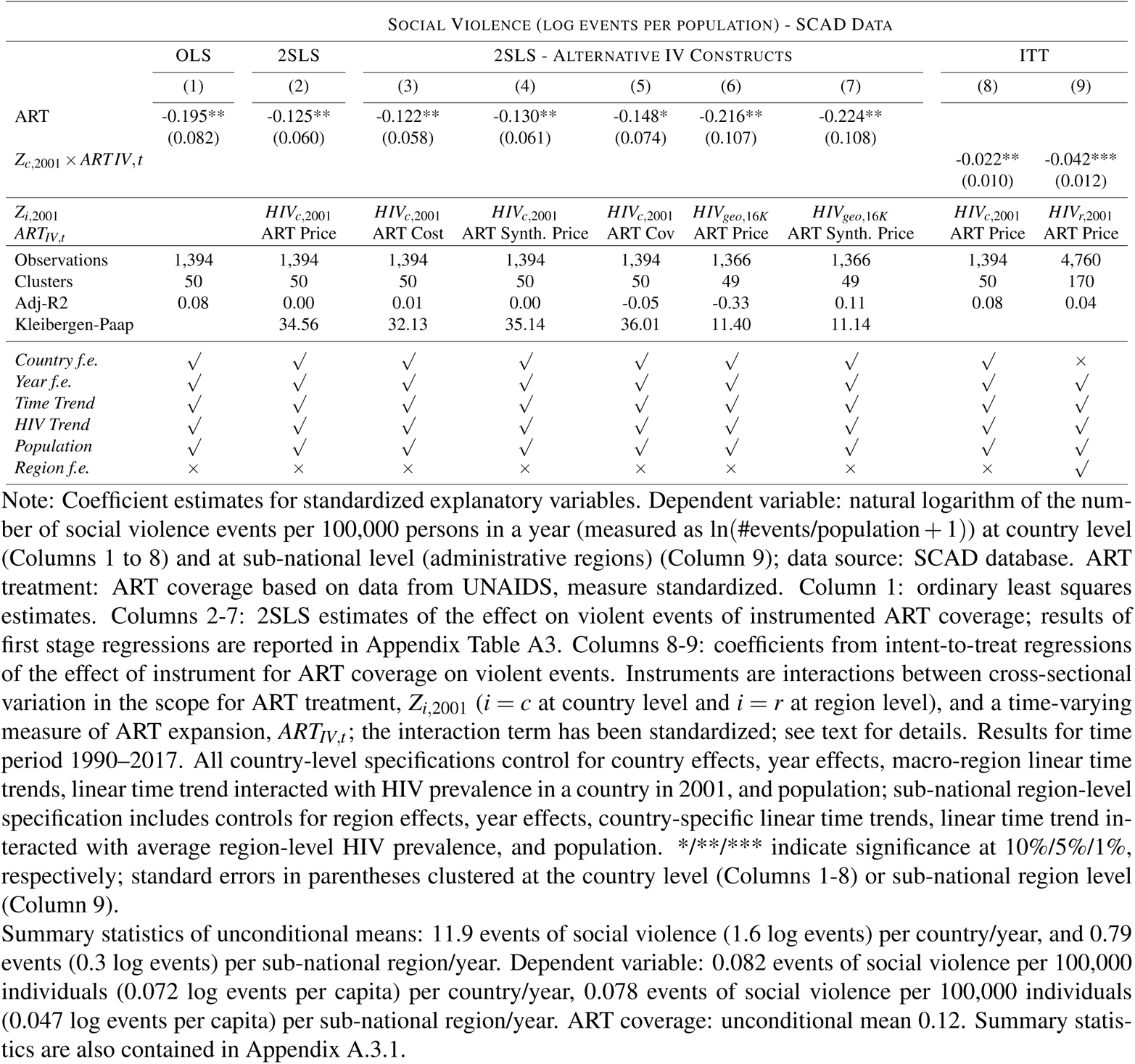
EFFECT OF ART EXPANSION ON VIOLENCE IN AFRICA: SCAD EVENTS PER CAPITA

#### A.4.5.3 Different Events and Participants

**Table A19:** DIFFERENT EVENTS AND PARTICIPANTS, COUNTRY ANALYSIS

| SOCIAL VIOLENCE (LOG EVENTS) - SCAD DATA |  |  |  |  |  |  |  |  |
| --- | --- | --- | --- | --- | --- | --- | --- | --- |
| Panel A |  | ART PRICE |  |  |  |  |  |  |
|  | (1)<br>No Strikes | (2)<br>Only Strikes | (3)<br>Health Workers | (4)<br>No Health Workers | (5)<br>Civil Servants | (6)<br>No Civil Servants | (7)<br>NGO | (8)<br>No NGO |
| ART | -0.899**<br>(0.401) | -0.504*<br>(0.266) | -0.121<br>(0.095) | -0.963***<br>(0.354) | -0.226<br>(0.178) | -0.960***<br>(0.350) | -0.160<br>(0.143) | -0.935**<br>(0.369) |
| Mean | 10.70 | 1.18 | 0.19 | 11.69 | 0.54 | 11.34 | 0.34 | 11.54 |
| Observations | 1,394 | 1,394 | 1,394 | 1,394 | 1,394 | 1,394 | 1,394 | 1,394 |
| Clusters | 50 | 50 | 50 | 50 | 50 | 50 | 50 | 50 |
| Adj-R2 | 0.20 | -0.03 | -0.02 | 0.17 | -0.00 | 0.18 | 0.12 | 0.17 |
| Kleibergen-Paap | 33.77 | 33.77 | 33.77 | 33.77 | 33.77 | 33.77 | 33.77 | 33.77 |
| Panel B |  | ART COST |  |  |  |  |  |  |
|  | (1) | (2) | (3) | (4) | (5) | (6) | (7) | (8) |
| ART | -0.869**<br>(0.395) | -0.515*<br>(0.264) | -0.137<br>(0.096) | -0.946**<br>(0.354) | -0.253<br>(0.182) | -0.934***<br>(0.348) | -0.171<br>(0.151) | -0.920**<br>(0.368) |
| Mean | 10.70 | 1.18 | 0.19 | 11.69 | 0.54 | 11.34 | 0.34 | 11.54 |
| Observations | 1,394 | 1,394 | 1,394 | 1,394 | 1,394 | 1,394 | 1,394 | 1,394 |
| Clusters | 50 | 50 | 50 | 50 | 50 | 50 | 50 | 50 |
| Adj-R2 | 0.21 | -0.03 | -0.02 | 0.18 | -0.01 | 0.18 | 0.12 | 0.17 |
| Kleibergen-Paap | 31.42 | 31.42 | 31.42 | 31.42 | 31.42 | 31.42 | 31.42 | 31.42 |
| Panel C |  | ART COVERAGE |  |  |  |  |  |  |
|  | (1) | (2) | (3) | (4) | (5) | (6) | (7) | (8) |
| ART | -0.924**<br>(0.410) | -0.581**<br>(0.254) | -0.114<br>(0.093) | -0.985**<br>(0.378) | -0.368*<br>(0.193) | -0.945**<br>(0.365) | -0.168<br>(0.174) | -0.977**<br>(0.392) |
| Mean | 10.70 | 1.18 | 0.19 | 11.69 | 0.54 | 11.34 | 0.34 | 11.54 |
| Observations | 1,394 | 1,394 | 1,394 | 1,394 | 1,394 | 1,394 | 1,394 | 1,394 |
| Clusters | 50 | 50 | 50 | 50 | 50 | 50 | 50 | 50 |
| Adj-R2 | 0.20 | -0.05 | -0.02 | 0.17 | -0.03 | 0.18 | 0.12 | 0.17 |
| Kleibergen-Paap | 34.69 | 34.69 | 34.69 | 34.69 | 34.69 | 34.69 | 34.69 | 34.69 |
| Country f.e | ✓ | ✓ | ✓ | ✓ | ✓ | ✓ | ✓ | ✓ |
| Year f.e | ✓ | ✓ | ✓ | ✓ | ✓ | ✓ | ✓ | ✓ |
| Time Trend | ✓ | ✓ | ✓ | ✓ | ✓ | ✓ | ✓ | ✓ |
| HIV Trend | ✓ | ✓ | ✓ | ✓ | ✓ | ✓ | ✓ | ✓ |
| Population | ✓ | ✓ | ✓ | ✓ | ✓ | ✓ | ✓ | ✓ |

Note: 2SLS estimates (second stage coefficients). Dependent variable: natural logarithm of the number of social conflict events of a particular type in a year (measured as $\ln(\#events + 1)$ ) at country level; data source: SCAD database. ART treatment: ART coverage based on data from UNAIDS. Instruments are interactions between HIV prevalence in a country in 2001 and different time-varying measures (Panel A: price of ART treatment; Panel B: cost of main agents of ART; Panel C: ART coverage in countries outside Africa). Results for time period 1990–2017. All specifications control for country effects, year effects, macro-region linear time trends, linear time trend interacted with HIV prevalence in a country in 2001, and population. \*/\*\*/\*\* indicate significance at 10%/5%/1%, respectively, standard errors in parentheses clustered at the country level.
Different event types and participants based on narratives in SCAD documentation. Only events with the respective words in narratives are included (or excluded) in the respective columns.

**Table A20:** EVENTS AND PARTICIPANTS, SUB-NATIONAL ANALYSIS

| SOCIAL VIOLENCE (LOG EVENTS) - SCAD DATA |  |  |  |  |  |  |  |  |
| --- | --- | --- | --- | --- | --- | --- | --- | --- |
| Panel A |  | ART PRICE |  |  |  |  |  |  |
|  | (1) | (2) | (3) | (4) | (5) | (6) | (7) | (8) |
|  | No Strikes | Only Strikes | Health Workers | No Health Workers | Civil Servants | No Civil Servants | NGO | No NGO |
| $HIV_{r,2001} \times ART_{IV,t}$ | -0.103***<br>(0.028) | -0.037**<br>(0.014) | -0.006<br>(0.004) | -0.165***<br>(0.034) | -0.016**<br>(0.007) | -0.159***<br>(0.033) | 0.007<br>(0.008) | -0.166***<br>(0.034) |
| Mean | 0.74 | 0.06 | 0.01 | 0.78 | 0.03 | 0.76 | 0.02 | 0.77 |
| Observations | 4,760 | 4,760 | 4,760 | 4,760 | 4,760 | 4,760 | 4,760 | 4,760 |
| Clusters | 170 | 170 | 170 | 170 | 170 | 170 | 170 | 170 |
| Adj-R2 | 0.07 | -0.01 | -0.01 | 0.09 | -0.01 | 0.08 | -0.00 | 0.09 |
| Panel B |  | ART COST |  |  |  |  |  |  |
|  | (1) | (2) | (3) | (4) | (5) | (6) | (7) | (8) |
| $Z_{r,2001} \times ART_{IV,t}$ | -0.097***<br>(0.027) | -0.036***<br>(0.014) | -0.006<br>(0.004) | -0.156***<br>(0.032) | -0.016**<br>(0.007) | -0.150***<br>(0.031) | 0.007<br>(0.008) | -0.157***<br>(0.033) |
| Mean | 0.74 | 0.06 | 0.01 | 0.78 | 0.03 | 0.76 | 0.02 | 0.77 |
| Observations | 4,760 | 4,760 | 4,760 | 4,760 | 4,760 | 4,760 | 4,760 | 4,760 |
| Clusters | 170 | 170 | 170 | 170 | 170 | 170 | 170 | 170 |
| Adj-R2 | 0.07 | -0.01 | -0.01 | 0.08 | -0.01 | 0.08 | -0.00 | 0.08 |
| Panel C |  | ART Cov |  |  |  |  |  |  |
|  | (1) | (2) | (3) | (4) | (5) | (6) | (7) | (8) |
| $Z_{r,2001} \times ART_{IV,t}$ | -0.138***<br>(0.037) | -0.045**<br>(0.018) | -0.006<br>(0.007) | -0.215***<br>(0.046) | -0.033***<br>(0.012) | -0.198***<br>(0.044) | 0.010<br>(0.012) | -0.216***<br>(0.046) |
| Mean | 0.74 | 0.06 | 0.01 | 0.78 | 0.03 | 0.76 | 0.02 | 0.77 |
| Observations | 4,760 | 4,760 | 4,760 | 4,760 | 4,760 | 4,760 | 4,760 | 4,760 |
| Clusters | 170 | 170 | 170 | 170 | 170 | 170 | 170 | 170 |
| Adj-R2 | 0.07 | -0.01 | -0.01 | 0.08 | -0.01 | 0.08 | -0.00 | 0.08 |
| *Region f.e* | ✓ | ✓ | ✓ | ✓ | ✓ | ✓ | ✓ | ✓ |
| *Year f.e* | ✓ | ✓ | ✓ | ✓ | ✓ | ✓ | ✓ | ✓ |
| *Time Trend* | ✓ | ✓ | ✓ | ✓ | ✓ | ✓ | ✓ | ✓ |
| *HIV Trend* | ✓ | ✓ | ✓ | ✓ | ✓ | ✓ | ✓ | ✓ |
| *Population* | ✓ | ✓ | ✓ | ✓ | ✓ | ✓ | ✓ | ✓ |

Coefficient estimates (ITT). Dependent variable: natural logarithm of the number of social conflict events of a particular type in a year (measured as $\ln(\#events + 1)$ ) at sub-national region level; data source: SCAD database. Instruments are interactions between HIV prevalence in a country in 2001 and different time-varying measures (Panel A: price of ART treatment; Panel B: cost of main agents of ART; Panel C: ART coverage in countries outside Africa). Results for time period 1990–2017. All specifications control for region effects, year effects, country-specific linear time trends, linear time trend interacted with average region-level HIV prevalence, and population. \*/\*\*/\*\* indicate significance at 10%/5%/1%, respectively, standard errors in parentheses clustered at the sub-national region level.
Different event types and participants based on narratives in SCAD documentation. Only events with the respective words in narratives are included (or excluded) in the respective columns.

### A.4.6 Alternative Outcomes: Life Expectancy and GDP growth

**Table A21:** ALTERNATIVE OUTCOMES: GDP PER CAPITA (LOG)

|  | GDP P.C. (LOG) |  |  |  |
| --- | --- | --- | --- | --- |
|  | OLS | 2SLS |  |  |
|  | (1) | (2) | (3) | (4) |
| ART | 0.088<br>(0.063) | 0.088<br>(0.080) | 0.093<br>(0.082) | 0.069<br>(0.084) |
| $Z_{i,2001}$ | | $HIV2001_{c,2001}$ | $HIV2001_{c,2001}$ | $HIV2001_{c,2001}$ |
| $ART_{IV,t}$ | | ART Price | ART Cost | ART Cov |
| Observations | 1,299 | 1,299 | 1,299 | 1,299 |
| Clusters | 48 | 48 | 48 | 48 |
| Adj-R2 | 0.33 | 0.33 | 0.33 | 0.33 |
| Kleibergen-Paap |  | 30.30 | 28.20 | 31.01 |
| *Country f.e.* | ✓ | ✓ | ✓ | ✓ |
| *Year f.e.* | ✓ | ✓ | ✓ | ✓ |
| *Time Trend* | ✓ | ✓ | ✓ | ✓ |
| *HIV Trend* | ✓ | ✓ | ✓ | ✓ |
| *Population* | ✓ | ✓ | ✓ | ✓ |
| *Region f.e.* | ✓ | ✓ | ✓ | ✓ |

**Table A22:** ALTERNATIVE OUTCOMES: GROWTH OF GDP PER CAPITA (LOG)

| GROWTH OF GDP P.C. (LOG) |  |  |  |  |
| --- | --- | --- | --- | --- |
|  | OLS | 2SLS |  |  |
|  | (1) | (2) | (3) | (4) |
| ART | 0.011<br>(0.009) | 0.039<br>(0.030) | 0.036<br>(0.028) | 0.026<br>(0.021) |
| $Z_{i,2001}$ | | $HIV2001_{c,2001}$ | $HIV2001_{c,2001}$ | $HIV2001_{c,2001}$ |
| $ART_{IV,t}$ | | ART Price | ART Cost | ART Cov |
| Observations | 1,251 | 1,251 | 1,251 | 1,251 |
| Clusters | 47 | 47 | 47 | 47 |
| Adj-R2 | 0.02 | -0.00 | 0.00 | 0.01 |
| Kleibergen-Paap |  | 28.22 | 26.06 | 28.21 |
| *Country f.e.* | ✓ | ✓ | ✓ | ✓ |
| *Year f.e.* | ✓ | ✓ | ✓ | ✓ |
| *Time Trend* | ✓ | ✓ | ✓ | ✓ |
| *HIV Trend* | ✓ | ✓ | ✓ | ✓ |
| *Population* | ✓ | ✓ | ✓ | ✓ |
| *Region f.e.* | ✓ | ✓ | ✓ | ✓ |

**Table A23:** ALTERNATIVE OUTCOMES: LIFE EXPECTANCY

| LIFE EXPECTANCY (LOG) |  |  |  |  |
| --- | --- | --- | --- | --- |
|  | OLS | 2SLS |  |  |
|  | (1) | (2) | (3) | (4) |
| ART | 0.075***<br>(0.022) | 0.323***<br>(0.050) | 0.325***<br>(0.052) | 0.328***<br>(0.049) |
| $Z_{i,2001}$ | | $HIV2001_{c,2001}$ | $HIV2001_{c,2001}$ | $HIV2001_{c,2001}$ |
| $ART_{IV,t}$ | | ART Price | ART Cost | ART Cov |
| Observations | 1,394 | 1,394 | 1,394 | 1,394 |
| Clusters | 50 | 50 | 50 | 50 |
| Adj-R2 | 0.64 | -0.07 | -0.08 | -0.10 |
| Kleibergen-Paap |  | 33.77 | 31.42 | 34.69 |
| *Country f.e.* | ✓ | ✓ | ✓ | ✓ |
| *Year f.e.* | ✓ | ✓ | ✓ | ✓ |
| *Time Trend* | ✓ | ✓ | ✓ | ✓ |
| *HIV Trend* | ✓ | ✓ | ✓ | ✓ |
| *Population* | ✓ | ✓ | ✓ | ✓ |
| *Region f.e.* | ✓ | ✓ | ✓ | ✓ |

### A.4.7 Mediation Analysis: Economic Prosperity and Health

**Table A24:** MEDIATION: ART EXPANSION, INCOME, AND SOCIAL VIOLENCE

| SOCIAL VIOLENCE (LOG EVENTS) - SCAD DATA |  |  |  |  |
| --- | --- | --- | --- | --- |
|  | OLS | 2SLS |  |  |
|  | (1) | (2) | (3) | (4) |
| GDP p.c. (log) | -0.205*<br>(0.114) | -11.379<br>(11.104) | -10.625<br>(10.056) | -15.013<br>(19.318) |
| $Z_{i,2001}$ | | $HIV2001_{c,2001}$ | $HIV2001_{c,2001}$ | $HIV2001_{c,2001}$ |
| $ART_{IV,t}$ | | ART Price | ART Cost | ART Cov |
| Observations | 1,299 | 1,299 | 1,299 | 1,299 |
| Clusters | 48 | 48 | 48 | 48 |
| Adj-R2 | 0.21 | -8.81 | -7.63 | -15.63 |
| Kleibergen-Paap |  | 1.09 | 1.16 | 0.64 |
| *Country f.e.* | ✓ | ✓ | ✓ | ✓ |
| *Year f.e.* | ✓ | ✓ | ✓ | ✓ |
| *Time Trend* | ✓ | ✓ | ✓ | ✓ |
| *HIV Trend* | ✓ | ✓ | ✓ | ✓ |
| *Population* | ✓ | ✓ | ✓ | ✓ |
| *Region f.e.* | ✓ | ✓ | ✓ | ✓ |

**Table A25:** MEDIATION: ART EXPANSION, INCOME, AND SOCIAL VIOLENCE (UCDP)

| ONGOING ARMED CONFLICTS (LOG EVENTS) - UCDP DATA |  |  |  |  |
| --- | --- | --- | --- | --- |
|  | OLS | 2SLS |  |  |
|  | (1) | (2) | (3) | (4) |
| GDP p.c. (log) | -0.247<br>(0.224) | 0.857<br>(5.911) | 0.635<br>(5.637) | 2.848<br>(8.921) |
| $Z_{i,2001}$ | | $HIV2001_{c,2001}$ | $HIV2001_{c,2001}$ | $HIV2001_{c,2001}$ |
| $ART_{IV,t}$ | | ART Price | ART Cost | ART Cov |
| Observations | 1,299 | 1,299 | 1,299 | 1,299 |
| Clusters | 48 | 48 | 48 | 48 |
| Adj-R2 | 0.03 | -0.01 | 0.00 | -0.34 |
| Kleibergen-Paap |  | 1.09 | 1.16 | 0.64 |
| *Country f.e.* | ✓ | ✓ | ✓ | ✓ |
| *Year f.e.* | ✓ | ✓ | ✓ | ✓ |
| *Time Trend* | ✓ | ✓ | ✓ | ✓ |
| *HIV Trend* | ✓ | ✓ | ✓ | ✓ |
| *Population* | ✓ | ✓ | ✓ | ✓ |
| *Region f.e.* | ✓ | ✓ | ✓ | ✓ |

**Table A26:** MEDIATION: ART EXPANSION, GDP PER CAPITA GROWTH, AND SOCIAL VIOLENCE

| SOCIAL VIOLENCE (LOG EVENTS) - SCAD DATA |  |  |  |  |
| --- | --- | --- | --- | --- |
|  | OLS | 2SLS |  |  |
|  | (1) | (2) | (3) | (4) |
| Growth of GDP p.c. (log) | -0.826<br>(0.587) | -26.771<br>(21.275) | -28.066<br>(22.254) | -41.486<br>(34.714) |
| $Z_{i,2001}$ | | $HIV2001_{c,2001}$ | $HIV2001_{c,2001}$ | $HIV2001_{c,2001}$ |
| $ART_{IV,t}$ | | ART Price | ART Cost | ART Cov |
| Observations | 1,251 | 1,251 | 1,251 | 1,251 |
| Clusters | 47 | 47 | 47 | 47 |
| Adj-R2 | 0.19 | -3.29 | -3.64 | -8.34 |
| Kleibergen-Paap |  | 1.47 | 1.49 | 1.42 |
| *Country f.e.* | ✓ | ✓ | ✓ | ✓ |
| *Year f.e.* | ✓ | ✓ | ✓ | ✓ |
| *Time Trend* | ✓ | ✓ | ✓ | ✓ |
| *HIV Trend* | ✓ | ✓ | ✓ | ✓ |
| *Population* | ✓ | ✓ | ✓ | ✓ |
| *Region f.e.* | ✓ | ✓ | ✓ | ✓ |

**Table A27:** MEDIATION: ART EXPANSION, GDP PER CAPITA GROWTH, AND SOCIAL VIOLENCE (UCDP)

| ONGOING ARMED CONFLICTS (LOG EVENTS) - UCDP DATA |  |  |  |  |
| --- | --- | --- | --- | --- |
|  | OLS | 2SLS |  |  |
|  | (1) | (2) | (3) | (4) |
| Growth of GDP p.c. (log) | -2.614**<br>(1.009) | -3.940<br>(13.087) | -4.606<br>(14.115) | -2.926<br>(21.203) |
| $Z_{i,2001}$<br>$ART_{IV,t}$ | | $HIV2001_{c,2001}$<br>ART Price | $HIV2001_{c,2001}$<br>ART Cost | $HIV2001_{c,2001}$<br>ART Cov |
| Observations | 1,251 | 1,251 | 1,251 | 1,251 |
| Clusters | 47 | 47 | 47 | 47 |
| Adj-R2 | 0.04 | 0.03 | 0.03 | 0.04 |
| Kleibergen-Paap |  | 1.47 | 1.49 | 1.42 |
| *Country f.e.* | ✓ | ✓ | ✓ | ✓ |
| *Year f.e.* | ✓ | ✓ | ✓ | ✓ |
| *Time Trend* | ✓ | ✓ | ✓ | ✓ |
| *HIV Trend* | ✓ | ✓ | ✓ | ✓ |
| *Population* | ✓ | ✓ | ✓ | ✓ |
| *Region f.e.* | ✓ | ✓ | ✓ | ✓ |

**Table A28:** MEDIATION: ART EXPANSION, LIFE EXPECTANCY, AND SOCIAL VIOLENCE

| SOCIAL VIOLENCE (LOG EVENTS) - SCAD DATA |  |  |  |  |
| --- | --- | --- | --- | --- |
|  | OLS | 2SLS |  |  |
|  | (1) | (2) | (3) | (4) |
| Life Expectancy (log) | -1.555*<br>(0.847) | -2.989***<br>(1.026) | -2.938***<br>(1.016) | -3.046***<br>(1.049) |
| $Z_{i,2001}$ | | $HIV2001_{c,2001}$ | $HIV2001_{c,2001}$ | $HIV2001_{c,2001}$ |
| $ART_{IV,t}$ | | ART Price | ART Cost | ART Cov |
| Observations | 1,394 | 1,394 | 1,394 | 1,394 |
| Clusters | 50 | 50 | 50 | 50 |
| Adj-R2 | 0.23 | 0.22 | 0.22 | 0.22 |
| Kleibergen-Paap |  | 140.01 | 133.44 | 143.01 |
| *Country f.e.* | ✓ | ✓ | ✓ | ✓ |
| *Year f.e.* | ✓ | ✓ | ✓ | ✓ |
| *Time Trend* | ✓ | ✓ | ✓ | ✓ |
| *HIV Trend* | ✓ | ✓ | ✓ | ✓ |
| *Population* | ✓ | ✓ | ✓ | ✓ |
| *Region f.e.* | ✓ | ✓ | ✓ | ✓ |

**Table A29:** MEDIATION: ART EXPANSION, LIFE EXPECTANCY, AND SOCIAL VIOLENCE (UCDP)

| ONGOING ARMED CONFLICTS (LOG EVENTS) - UCDP DATA |  |  |  |  |
| --- | --- | --- | --- | --- |
|  | OLS | 2SLS |  |  |
|  | (1) | (2) | (3) | (4) |
| Life Expectancy (log) | -3.638**<br>(1.636) | -0.357<br>(1.485) | -0.396<br>(1.491) | 0.038<br>(1.594) |
| $Z_{i,2001}$ | | $HIV2001_{c,2001}$ | $HIV2001_{c,2001}$ | $HIV2001_{c,2001}$ |
| $ART_{IV,t}$ | | ART Price | ART Cost | ART Cov |
| Observations | 1,394 | 1,394 | 1,394 | 1,394 |
| Clusters | 50 | 50 | 50 | 50 |
| Adj-R2 | 0.05 | 0.02 | 0.03 | 0.02 |
| Kleibergen-Paap |  | 140.01 | 133.44 | 143.01 |
| *Country f.e.* | ✓ | ✓ | ✓ | ✓ |
| *Year f.e.* | ✓ | ✓ | ✓ | ✓ |
| *Time Trend* | ✓ | ✓ | ✓ | ✓ |
| *HIV Trend* | ✓ | ✓ | ✓ | ✓ |
| *Population* | ✓ | ✓ | ✓ | ✓ |
| *Region f.e.* | ✓ | ✓ | ✓ | ✓ |

#### A.4.7.1 Social Violence, ACLED

**Figure A12:**
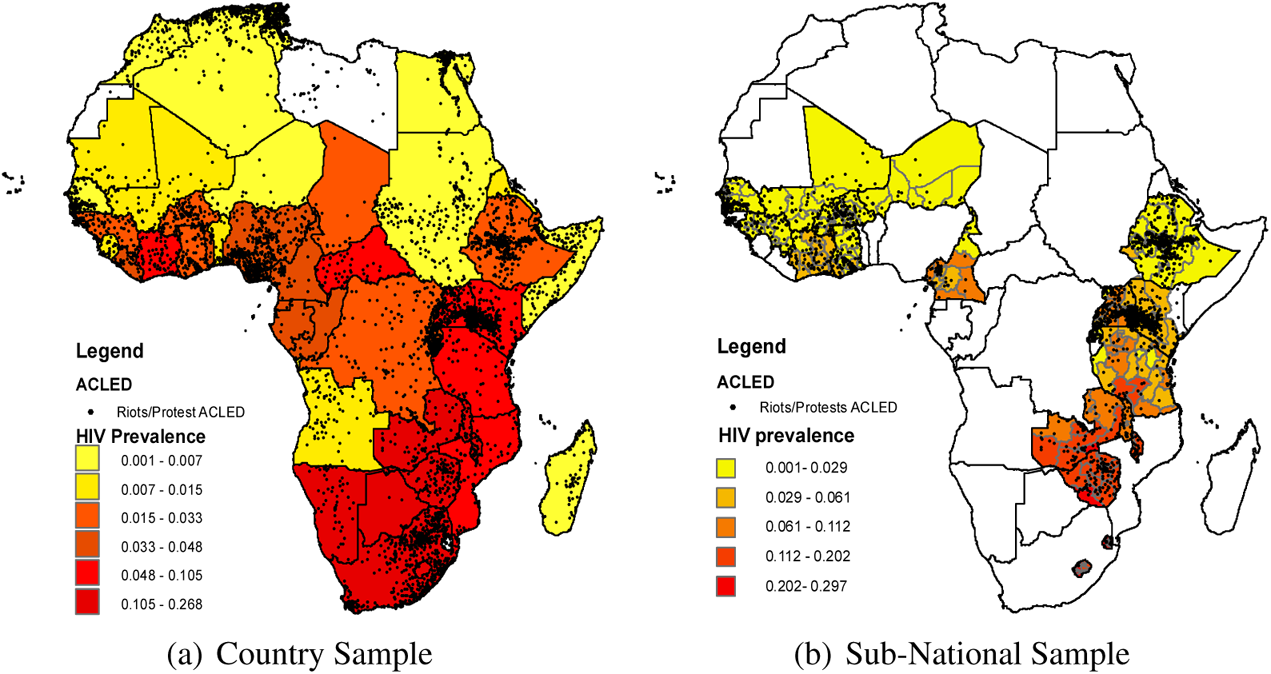
HIV PREVALENCE AND SOCIAL VIOLENCE IN AFRICA: ACLED Note: See Appendix A.3.1 for summary statistics.

**Table A30:** EFFECT OF ART EXPANSION ON VIOLENCE IN AFRICA: ACLED

| SOCIAL VIOLENCE (LOG EVENTS PER POPULATION) - ACLED DATA |  |  |  |  |  |  |  |  |  |
| --- | --- | --- | --- | --- | --- | --- | --- | --- | --- |
|  | OLS | 2SLS | 2SLS - ALTERNATIVE IV CONSTRUCTS |  |  |  |  | ITT |  |
|  | (1) | (2) | (3) | (4) | (5) | (6) | (7) | (8) | (9) |
| ART | -0.483<br>(0.920) | -1.512*<br>(0.778) | -1.611*<br>(0.809) | -1.430*<br>(0.738) | -1.809*<br>(0.912) | -1.556**<br>(0.715) | -1.530*<br>(0.763) |  |  |
| $Z_{c,2001} \times ART_{IV,t}$ | | | | | | | | -0.221**<br>(0.091) | -0.182***<br>(0.049) |
| $Z_{i,2001}$ | | $HIV_{c,2001}$ | $HIV_{c,2001}$ | $HIV_{c,2001}$ | $HIV_{c,2001}$ | $HIV_{geo,16K}$ | $HIV_{geo,16K}$ | $HIV_{c,2001}$ | $HIV_{r,2001}$ |
| $ART_{IV,t}$ | | ART Price | ART Cost | ART Synth. Price | ART Cov | ART Price | ART Synth. Price | ART Price | ART Price |
| Observations | 1,044 | 1,044 | 1,044 | 1,044 | 1,044 | 1,023 | 1,023 | 1,044 | 3,570 |
| Clusters | 50 | 50 | 50 | 50 | 50 | 49 | 49 | 50 | 170 |
| Adj-R2 | 0.45 | 0.32 | 0.30 | 0.33 | 0.26 | 0.29 | 0.29 | 0.45 | 0.33 |
| Kleibergen-Paap |  | 13.25 | 11.33 | 13.52 | 7.66 | 9.91 | 9.06 |  |  |
| Country f.e. | ✓ | ✓ | ✓ | ✓ | ✓ | ✓ | ✓ | ✓ | × |
| Year f.e. | ✓ | ✓ | ✓ | ✓ | ✓ | ✓ | ✓ | ✓ | ✓ |
| Time Trend | ✓ | ✓ | ✓ | ✓ | ✓ | ✓ | ✓ | ✓ | ✓ |
| HIV Trend | ✓ | ✓ | ✓ | ✓ | ✓ | ✓ | ✓ | ✓ | ✓ |
| Population | ✓ | ✓ | ✓ | ✓ | ✓ | ✓ | ✓ | ✓ | ✓ |
| Region f.e. | × | × | × | × | × | × | × | × | ✓ |

**Table A31:** FIRST STAGE RESULTS: Table A30

| ART TREATMENT (STANDARDISED) |  |  |  |  |  |  |
| --- | --- | --- | --- | --- | --- | --- |
|  | 2SLS | 2SLS - ALTERNATIVE IV CONSTRUCTS |  |  |  |  |
|  | (1) | (2) | (3) | (4) | (5) | (6) |
| $Z_{c,2001} \times ART_{IV,t}$ | 1.154***<br>(0.317) | 1.289***<br>(0.383) | 0.180***<br>(0.049) | 21.032***<br>(7.601) | 0.196***<br>(0.062) | 0.229***<br>(0.076) |
| $Z_{i,2001}$ | $HIV_{c,2001}$ | $HIV_{c,2001}$ | $HIV_{c,2001}$ | $HIV_{c,2001}$ | $HIV_{geo,16K}$ | $HIV_{geo,16K}$ |
| $ART_{IV,t}$ | ART Price | ART Cost | ART Synth. Price | ART Cov | ART Price | ART Synth. Price |
| Observations | 1,044 | 1,044 | 1,044 | 1,044 | 1,023 | 1,023 |
| Clusters | 50 | 50 | 50 | 50 | 49 | 49 |
| Kleibergen-Paap | 13.25 | 11.33 | 13.52 | 7.66 | 9.91 | 9.06 |
| Country f.e. | ✓ | ✓ | ✓ | ✓ | ✓ | ✓ |
| Year f.e. | ✓ | ✓ | ✓ | ✓ | ✓ | ✓ |
| Time Trend | ✓ | ✓ | ✓ | ✓ | ✓ | ✓ |
| HIV Trend | ✓ | ✓ | ✓ | ✓ | ✓ | ✓ |
| Population | ✓ | ✓ | ✓ | ✓ | ✓ | ✓ |

Note: Coefficient estimates (OLS). Estimates represent first stage results for the corresponding outcome equations (columns) in Figure ED2. Dependent variable: ART coverage (relative to infected) in a year at country level, based on data from UNAIDS. Instruments are interactions between cross-sectional variation in the scope for ART treatment, $Z_{i,2001}$ ( $i = c$ at country level and $i = r$ at region level), and a time-varying measure of ART expansion, $ART_{IV,t}$ , see text for details. Results for time period 1997–2017. All country-level specifications control for country effects, year effects, macro-region linear time trends, linear time trend interacted with HIV prevalence in a country in 2001, and population. \*/\*\*/\*\* indicate significance at 10%/5%/1%, respectively; standard errors in parentheses clustered at the country level.

#### A.4.7.2 Social Violence, GDELT

**Table A32:** SOCIAL VIOLENCE, GDELT

|  | SOCIAL VIOLENCE (LOG EVENTS) - GDELT |  |  |  |
| --- | --- | --- | --- | --- |
|  | (1) | (2) | (3) | (4) |
| ART | -0.284<br>(0.189) | -0.800**<br>(0.371) | -0.761*<br>(0.382) | -0.719*<br>(0.389) |
| $HIV_{c,2001} \times ART_{IV,t}$ | | ART Price | ART Cost | ART Cov |
| Observations | 1,394 | 1,394 | 1,394 | 1,394 |
| Clusters | 50 | 50 | 50 | 50 |
| Adj-R2 | 0.72 | 0.71 | 0.71 | 0.71 |
| Kleibergen-Paap |  | 33.77 | 31.42 | 34.69 |
| *Country f.e* | ✓ | ✓ | ✓ | ✓ |
| *Year f.e* | ✓ | ✓ | ✓ | ✓ |
| *Time Trend* | ✓ | ✓ | ✓ | ✓ |
| *HIV Trend* | ✓ | ✓ | ✓ | ✓ |
| *Population* | ✓ | ✓ | ✓ | ✓ |

#### A.4.7.3 Large-Scale Armed Conflicts

**Table A33:**
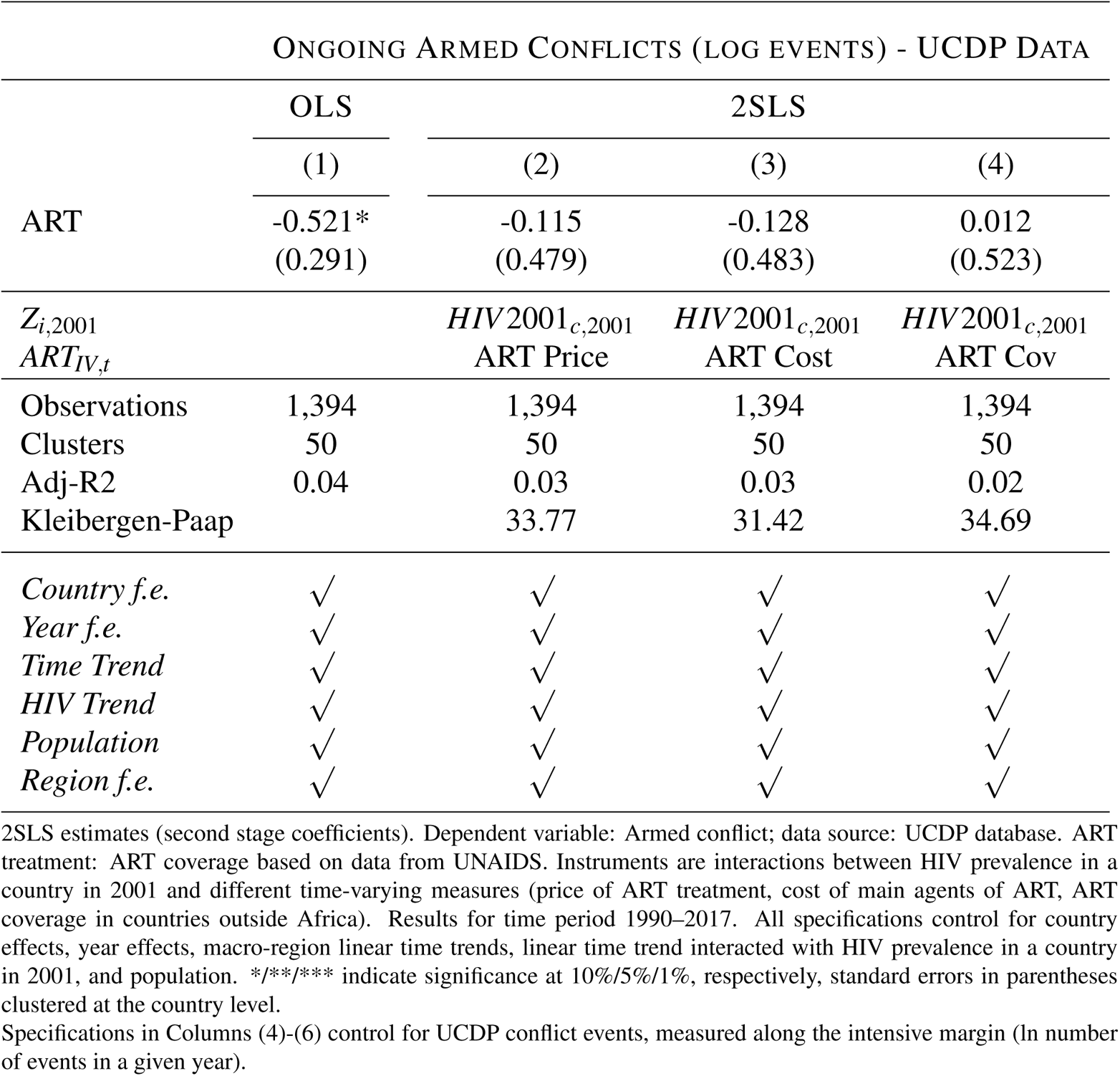
ART EXPANSION AND ARMED CONFLICT (UCDP)

| ONGOING ARMED CONFLICTS (LOG EVENTS) - UCDP DATA |  |  |  |  |
| --- | --- | --- | --- | --- |
|  | OLS | 2SLS |  |  |
|  | (1) | (2) | (3) | (4) |
| ART | -0.521*<br>(0.291) | -0.115<br>(0.479) | -0.128<br>(0.483) | 0.012<br>(0.523) |
| $Z_{i,2001}$ | | $HIV2001_{c,2001}$ | $HIV2001_{c,2001}$ | $HIV2001_{c,2001}$ |
| $ART_{IV,t}$ | | ART Price | ART Cost | ART Cov |
| Observations | 1,394 | 1,394 | 1,394 | 1,394 |
| Clusters | 50 | 50 | 50 | 50 |
| Adj-R2 | 0.04 | 0.03 | 0.03 | 0.02 |
| Kleibergen-Paap |  | 33.77 | 31.42 | 34.69 |
| *Country f.e.* | ✓ | ✓ | ✓ | ✓ |
| *Year f.e.* | ✓ | ✓ | ✓ | ✓ |
| *Time Trend* | ✓ | ✓ | ✓ | ✓ |
| *HIV Trend* | ✓ | ✓ | ✓ | ✓ |
| *Population* | ✓ | ✓ | ✓ | ✓ |
| *Region f.e.* | ✓ | ✓ | ✓ | ✓ |

#### A.4.7.4 Casualties and Size of Events

**Table A34:**
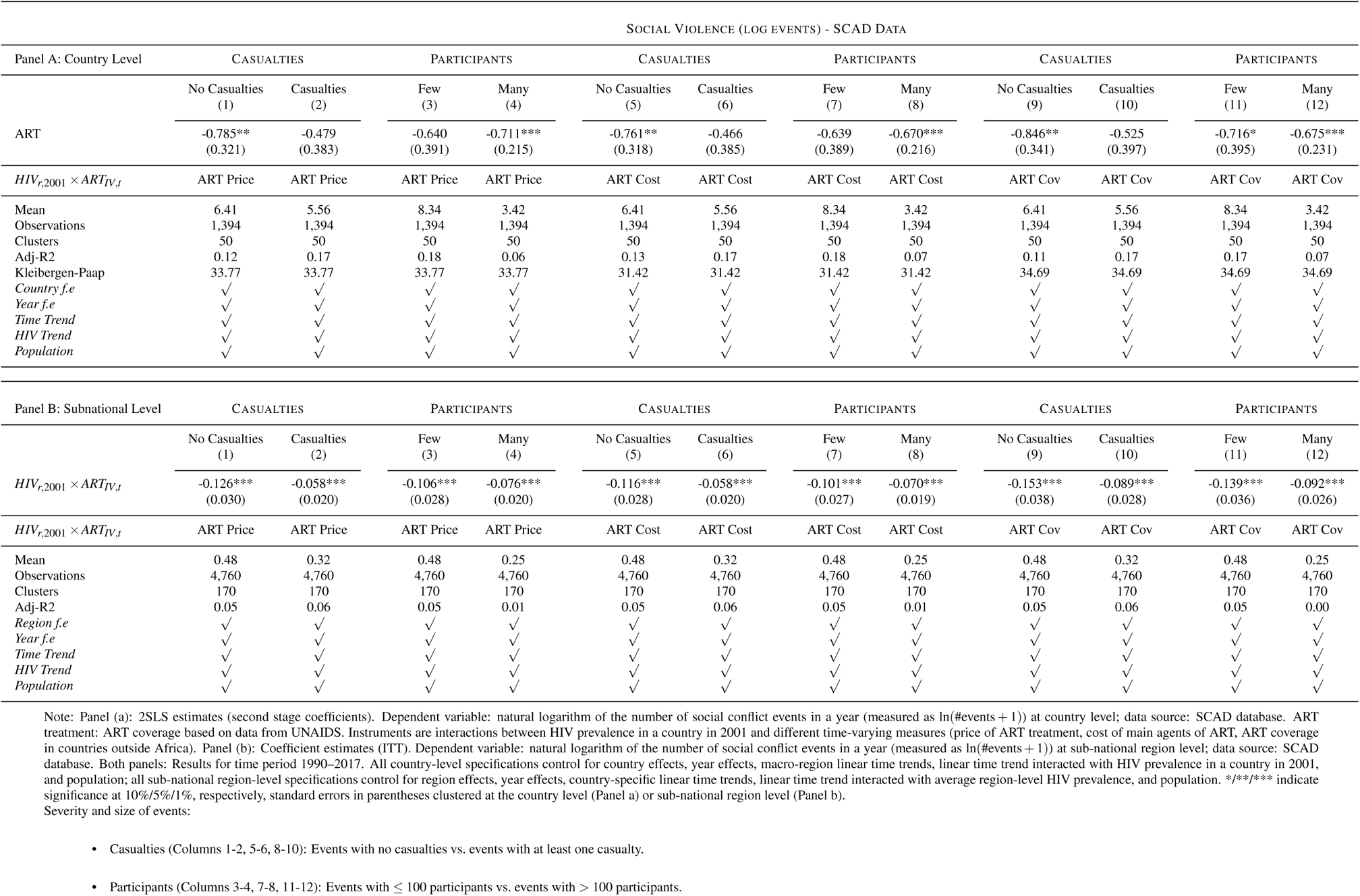
CASUALTIES AND SIZE OF EVENTS

| SOCIAL VIOLENCE (LOG EVENTS) - SCAD DATA |  |  |  |  |  |  |  |  |  |  |  |  |
| --- | --- | --- | --- | --- | --- | --- | --- | --- | --- | --- | --- | --- |
| Panel A: Country Level | CASUALTIES |  | PARTICIPANTS |  | CASUALTIES |  | PARTICIPANTS |  | CASUALTIES |  | PARTICIPANTS |  |
|  | No Casualties<br>(1) | Casualties<br>(2) | Few<br>(3) | Many<br>(4) | No Casualties<br>(5) | Casualties<br>(6) | Few<br>(7) | Many<br>(8) | No Casualties<br>(9) | Casualties<br>(10) | Few<br>(11) | Many<br>(12) |
| ART | -0.785**<br>(0.321) | -0.479<br>(0.383) | -0.640<br>(0.391) | -0.711***<br>(0.215) | -0.761**<br>(0.318) | -0.466<br>(0.385) | -0.639<br>(0.389) | -0.670***<br>(0.216) | -0.846**<br>(0.341) | -0.525<br>(0.397) | -0.716*<br>(0.395) | -0.675***<br>(0.231) |
| $HIV_{r,2001} \times ART_{IV,t}$ | ART Price | ART Price | ART Price | ART Price | ART Cost | ART Cost | ART Cost | ART Cost | ART Cov | ART Cov | ART Cov | ART Cov |
| Mean | 6.41 | 5.56 | 8.34 | 3.42 | 6.41 | 5.56 | 8.34 | 3.42 | 6.41 | 5.56 | 8.34 | 3.42 |
| Observations | 1,394 | 1,394 | 1,394 | 1,394 | 1,394 | 1,394 | 1,394 | 1,394 | 1,394 | 1,394 | 1,394 | 1,394 |
| Clusters | 50 | 50 | 50 | 50 | 50 | 50 | 50 | 50 | 50 | 50 | 50 | 50 |
| Adj-R2 | 0.12 | 0.17 | 0.18 | 0.06 | 0.13 | 0.17 | 0.18 | 0.07 | 0.11 | 0.17 | 0.17 | 0.07 |
| Kleibergen-Paap | 33.77 | 33.77 | 33.77 | 33.77 | 31.42 | 31.42 | 31.42 | 31.42 | 34.69 | 34.69 | 34.69 | 34.69 |
| Country *f.e* | ✓ | ✓ | ✓ | ✓ | ✓ | ✓ | ✓ | ✓ | ✓ | ✓ | ✓ | ✓ |
| Year *f.e* | ✓ | ✓ | ✓ | ✓ | ✓ | ✓ | ✓ | ✓ | ✓ | ✓ | ✓ | ✓ |
| Time Trend | ✓ | ✓ | ✓ | ✓ | ✓ | ✓ | ✓ | ✓ | ✓ | ✓ | ✓ | ✓ |
| HIV Trend | ✓ | ✓ | ✓ | ✓ | ✓ | ✓ | ✓ | ✓ | ✓ | ✓ | ✓ | ✓ |
| Population | ✓ | ✓ | ✓ | ✓ | ✓ | ✓ | ✓ | ✓ | ✓ | ✓ | ✓ | ✓ |
| Panel B: Subnational Level | CASUALTIES |  | PARTICIPANTS |  | CASUALTIES |  | PARTICIPANTS |  | CASUALTIES |  | PARTICIPANTS |  |
|  | No Casualties<br>(1) | Casualties<br>(2) | Few<br>(3) | Many<br>(4) | No Casualties<br>(5) | Casualties<br>(6) | Few<br>(7) | Many<br>(8) | No Casualties<br>(9) | Casualties<br>(10) | Few<br>(11) | Many<br>(12) |
| $HIV_{r,2001} \times ART_{IV,t}$ | -0.126***<br>(0.030) | -0.058***<br>(0.020) | -0.106***<br>(0.028) | -0.076***<br>(0.020) | -0.116***<br>(0.028) | -0.058***<br>(0.020) | -0.101***<br>(0.027) | -0.070***<br>(0.019) | -0.153***<br>(0.038) | -0.089***<br>(0.028) | -0.139***<br>(0.036) | -0.092***<br>(0.026) |
| $HIV_{r,2001} \times ART_{IV,t}$ | ART Price | ART Price | ART Price | ART Price | ART Cost | ART Cost | ART Cost | ART Cost | ART Cov | ART Cov | ART Cov | ART Cov |
| Mean | 0.48 | 0.32 | 0.48 | 0.25 | 0.48 | 0.32 | 0.48 | 0.25 | 0.48 | 0.32 | 0.48 | 0.25 |
| Observations | 4,760 | 4,760 | 4,760 | 4,760 | 4,760 | 4,760 | 4,760 | 4,760 | 4,760 | 4,760 | 4,760 | 4,760 |
| Clusters | 170 | 170 | 170 | 170 | 170 | 170 | 170 | 170 | 170 | 170 | 170 | 170 |
| Adj-R2 | 0.05 | 0.06 | 0.05 | 0.01 | 0.05 | 0.06 | 0.05 | 0.01 | 0.05 | 0.06 | 0.05 | 0.00 |
| Region *f.e* | ✓ | ✓ | ✓ | ✓ | ✓ | ✓ | ✓ | ✓ | ✓ | ✓ | ✓ | ✓ |
| Year *f.e* | ✓ | ✓ | ✓ | ✓ | ✓ | ✓ | ✓ | ✓ | ✓ | ✓ | ✓ | ✓ |
| Time Trend | ✓ | ✓ | ✓ | ✓ | ✓ | ✓ | ✓ | ✓ | ✓ | ✓ | ✓ | ✓ |
| HIV Trend | ✓ | ✓ | ✓ | ✓ | ✓ | ✓ | ✓ | ✓ | ✓ | ✓ | ✓ | ✓ |
| Population | ✓ | ✓ | ✓ | ✓ | ✓ | ✓ | ✓ | ✓ | ✓ | ✓ | ✓ | ✓ |

#### A.4.7.5 Ongoing Armed Conflicts

**Table A35:** ARMED CONFLICT (UCDP) AS ADDITIONAL CONTROL VARIABLE

| SOCIAL VIOLENCE (LOG EVENTS) - SCAD DATA |  |  |  |  |  |  |
| --- | --- | --- | --- | --- | --- | --- |
|  | (1) | (2) | (3) | (4) | (5) | (6) |
| ART | -0.966**<br>(0.362) | -0.954**<br>(0.361) | -0.999**<br>(0.388) | -0.946***<br>(0.333) | -0.931***<br>(0.331) | -1.001***<br>(0.357) |
| $HIV_{c,2001} \times ART_{IV,t}$ | ART Price | ART Cost | ART Cov | ART Price | ART Cost | ART Cov |
| Observations | 1,394 | 1,394 | 1,394 | 1,394 | 1,394 | 1,394 |
| Clusters | 50 | 50 | 50 | 50 | 50 | 50 |
| Adj-R2 | 0.17 | 0.18 | 0.17 | 0.24 | 0.24 | 0.22 |
| Kleibergen-Paap | 33.77 | 31.42 | 34.69 | 33.87 | 31.46 | 34.88 |
| *Country f.e.* | ✓ | ✓ | ✓ | ✓ | ✓ | ✓ |
| *Year f.e.* | ✓ | ✓ | ✓ | ✓ | ✓ | ✓ |
| *Time Trend* | ✓ | ✓ | ✓ | ✓ | ✓ | ✓ |
| *HIV Trend* | ✓ | ✓ | ✓ | ✓ | ✓ | ✓ |
| *Population* | ✓ | ✓ | ✓ | ✓ | ✓ | ✓ |
| *UCDP Events* | × | × | × | ✓ | ✓ | ✓ |

2SLS estimates (second stage coefficients). Dependent variable: natural logarithm of the number of social conflict events in a year (measured as $\ln(\#events + 1)$ ) at country level; data source: SCAD database. ART treatment: ART coverage based on data from UNAIDS. Instruments are interactions between HIV prevalence in a country in 2001 and different time-varying measures (price of ART treatment, cost of main agents of ART, ART coverage in countries outside Africa). Results for time period 1990–2017. All specifications control for country effects, year effects, macro-region linear time trends, linear time trend interacted with HIV prevalence in a country in 2001, and population. \*\*\*/\*\*\* indicate significance at 10%/5%/1%, respectively, standard errors in parentheses clustered at the country level.
Specifications in Columns (4)-(6) control for UCDP conflict events, measured along the intensive margin ( $\ln$ number of events in a given year).

#### A.4.7.6 Intimate Partner Violence, DHS

The DHS survey data for intimate partner violence only contain information for 124 regions with 219 observations in total. This requires the estimation of a modified empirical framework, because the specification with region fixed effects and year fixed effect would be too demanding for estimation. Consequently, different from the main analysis, the empirical specification contains a time trend and country fixed effects, which capture unobservable variation at the country level related to institutions or variation in data collection. The empirical framework is given by

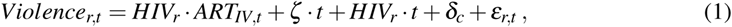

where

- *Violence_r,t_* is incidence of social violence in subnational region *r* in year *t*;
- *HIV_r_ · ART_IV,t_* represents the instruments as described in the baseline analysis;
- *ζ · t* is a linear time trend;
- *HIV_r_ ·t* is a trend of HIV prevalence, allowing for different trends according to pre-expansion HIV prevalence;
- *δ_c_* represents a country fixed effect.

**Figure A13:**
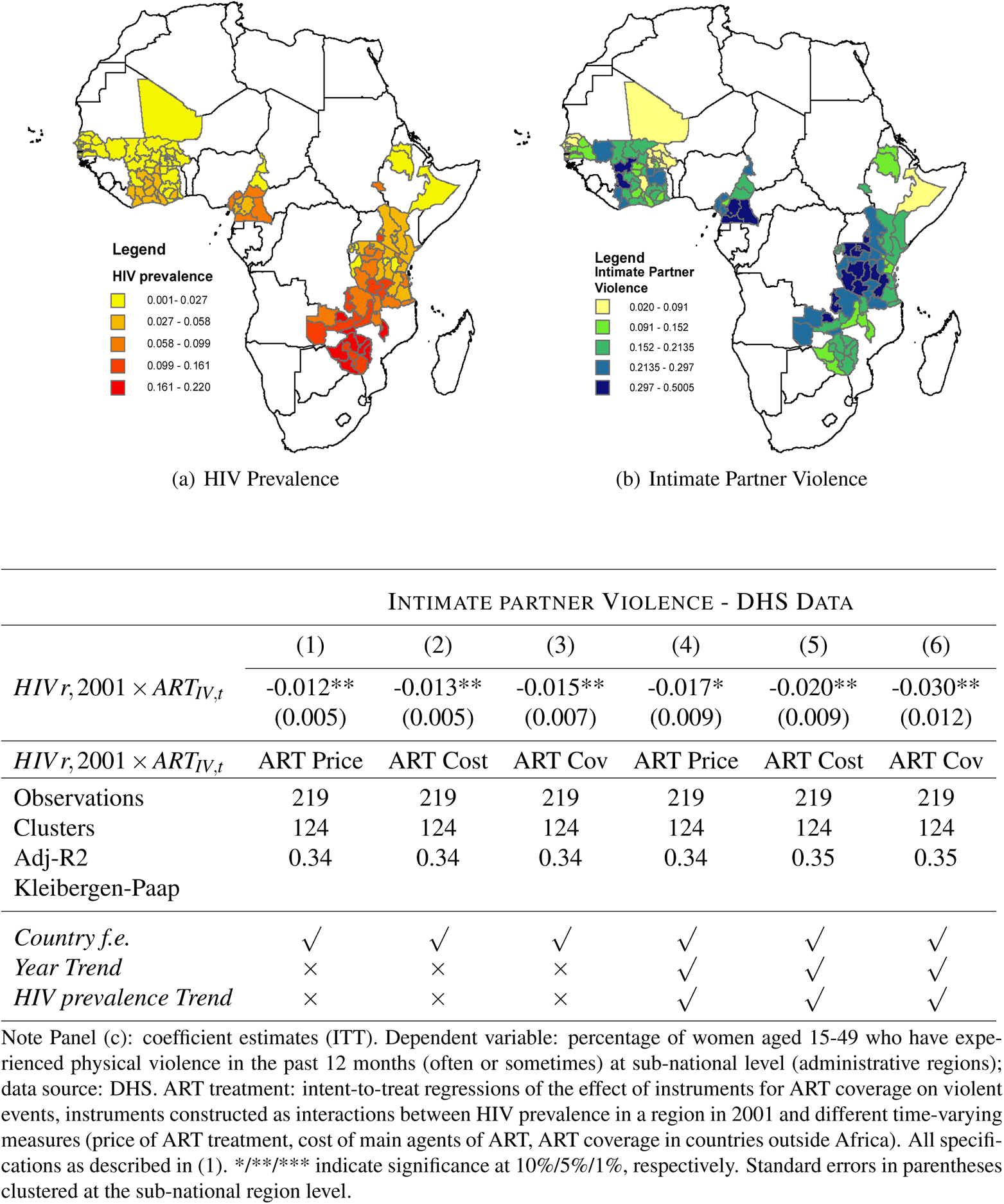
INTIMATE PARTNER VIOLENCE, DHS

### A.4.8 Channels: Tables

#### A.4.8.1 Social Violence: Types and Motives

This section contains detailed estimation results that form the basis of some results presented in Figure 4 in the main text. Estimates are based on SCAD data (see data description for details).

**Table A36:** SOCIAL VIOLENCE: TYPES AND MOTIVE

| Panel A: Country Level |  |  |  |  |  |
| --- | --- | --- | --- | --- | --- |
| *Dependent Variable* | TYPE OF EVENT |  | MAIN MOTIVE |  |  |
|  | (1)<br>Spontaneous | (2)<br>Organized | (3)<br>Elections | (4)<br>Economic grievance | (5)<br>Human rights |
| ART | 0.188<br>(0.221) | -1.376***<br>(0.392) | -0.377*<br>(0.196) | -0.535**<br>(0.265) | -0.468<br>(0.330) |
| $HIV_{c,2001} \times ART_{IV,t}$ | ART Price | ART Price | ART Price | ART Price | ART Price |
| Observations | 1,394 | 1,394 | 1,394 | 1,394 | 1,394 |
| Countries | 50 | 50 | 50 | 50 | 50 |
| Adj-R2 | 0.11 | 0.09 | -0.03 | 0.06 | 0.01 |
| Kleibergen-Paap | 33.77 | 33.77 | 33.77 | 33.77 | 33.77 |
| *Country f.e* | ✓ | ✓ | ✓ | ✓ | ✓ |
| *Year f.e* | ✓ | ✓ | ✓ | ✓ | ✓ |
| *Time Trend* | ✓ | ✓ | ✓ | ✓ | ✓ |
| *HIV Trend* | ✓ | ✓ | ✓ | ✓ | ✓ |
| *Population* | ✓ | ✓ | ✓ | ✓ | ✓ |
| Panel B: Subnational Level: |  |  |  |  |  |
| *Dependent Variable* | TYPE OF EVENT |  | MAIN MOTIVE |  |  |
|  | (1)<br>Spontaneous | (2)<br>Organized | (3)<br>Elections | (4)<br>Economic grievance | (5)<br>Human rights |
| $HIV_{r,2001} \times ART_{IV,t}$ | -0.014<br>(0.014) | -0.177***<br>(0.033) | -0.018*<br>(0.011) | -0.049***<br>(0.018) | -0.050***<br>(0.019) |
| $HIV_{r,2001} \times ART_{IV,t}$ | ART Price | ART Price | ART Price | ART Price | ART Price |
| Mean |  |  |  |  |  |
| Observations | 4,760 | 4,760 | 4,760 | 4,760 | 4,760 |
| Regions | 170 | 170 | 170 | 170 | 170 |
| R2 | 0.07 | 0.12 | 0.02 | 0.03 | 0.04 |
| *Region f.e.* | ✓ | ✓ | ✓ | ✓ | ✓ |
| *Year f.e.* | ✓ | ✓ | ✓ | ✓ | ✓ |
| *Time Trend* | ✓ | ✓ | ✓ | ✓ | ✓ |
| *HIV Trend* | ✓ | ✓ | ✓ | ✓ | ✓ |
| *Population* | ✓ | ✓ | ✓ | ✓ | ✓ |

#### A.4.8.2 Individual Trust and Approval of Policy

This section contains detailed estimation results that form the basis of some results presented in Figure 6 in the main text. By documenting how trust in institutions or approval of government policy is affected by ART coverage, the estimates provide insights for the mechanism underlying the main results. The analysis is based on survey data from the Afrobarometer. Given the data structure of Afrobarometer surveys, which are collected in different rounds, the specification of the empirical framework needs to be slightly adjusted in comparison to the baseline analysis. In particular, year fixed effects are replaced by fixed effects for Afrobarometer round. These round fixed effects also account for variation in the precise wording of survey questions across waves. The empirical model is

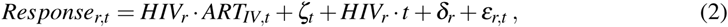

where

- *Response_r,t_* is the survey response of respondents in region *r* in survey round *t* to various questions about trust in institutions (parliament, local government, policy) or to questions about how the current government handles certain policies (related to HIV/AIDS, basic health provision, management of the economy, or combating crime);
- *HIV_r_ · ART_IV,t_* represents the instruments as described in the baseline analysis;
- *ζ_t_* round fixed effects;
- *HIV_r_ ·t* is a trend of HIV prevalence, allowing for different trends according to pre-expansion HIV prevalence across rounds;

• *δ_r_*: region fixed effects.

**Table A37:** APPROVAL TO GOVERNMENT POLICIES

| PANEL A | SPECIFIC POLICIES |  | GENERAL POLICIES |  |  |
| --- | --- | --- | --- | --- | --- |
|  | HIV/AIDS<br>(1) | Education<br>(2) | Economy<br>(3) | Health<br>(4) | Crime<br>(5) |
| $HIV_{r,2001} \times ART_{IV,t}$ | 0.088***<br>(0.029) | -0.005<br>(0.030) | 0.084**<br>(0.032) | 0.088***<br>(0.029) | 0.119***<br>(0.032) |
| $HIV_{r,2001} \times ART_{IV,t}$ | ART Price | ART Price | ART Price | ART Price | ART Price |
| Mean | 0.83 | 0.79 | 0.60 | 0.77 | 0.30 |
| Observations | 337 | 390 | 390 | 337 | 369 |
| Regions | 110 | 112 | 112 | 110 | 110 |
| R2 | 0.19 | 0.26 | 0.25 | 0.19 | 0.22 |
| PANEL B | SPECIFIC POLICIES |  | GENERAL POLICIES |  |  |
|  | HIV/AIDS<br>(1) | Education<br>(2) | Economy<br>(3) | Health<br>(4) | Crime<br>(5) |
| $HIV_{r,2001} \times ART_{IV,t}$ | 0.142***<br>(0.041) | 0.007<br>(0.040) | 0.099***<br>(0.036) | 0.142***<br>(0.041) | 0.130***<br>(0.041) |
| $HIV_{r,2001} \times ART_{IV,t}$ | ART Cost | ART Cost | ART Cost | ART Cost | ART Cost |
| Mean | 0.83 | 0.79 | 0.60 | 0.77 | 0.30 |
| Observations | 337 | 390 | 390 | 337 | 369 |
| Regions | 110 | 112 | 112 | 110 | 110 |
| R2 | 0.23 | 0.26 | 0.25 | 0.23 | 0.19 |
| PANEL C | SPECIFIC POLICIES |  | GENERAL POLICIES |  |  |
|  | HIV/AIDS<br>(1) | Education<br>(2) | Economy<br>(3) | Health<br>(4) | Crime<br>(5) |
| $HIV_{r,2001} \times ART_{IV,t}$ | 0.334***<br>(0.104) | -0.025<br>(0.094) | 0.255***<br>(0.077) | 0.202**<br>(0.082) | 0.134**<br>(0.066) |
| *Instrument* | ART Cov | ART Cov | ART Cov | ART Cov | ART Cov |
| Mean | 0.83 | 0.79 | 0.60 | 0.77 | 0.30 |
| Observations | 337 | 390 | 390 | 390 | 390 |
| Regions | 110 | 112 | 112 | 112 | 112 |
| R2 | 0.23 | 0.26 | 0.61 | 0.22 | 0.82 |
| *Region fixed effects* | ✓ | ✓ | ✓ | ✓ | ✓ |
| *Year fixed effects* | ✓ | ✓ | ✓ | ✓ | ✓ |
| *HIV prevalence Trend* | ✓ | ✓ | ✓ | ✓ | ✓ |

Notes: Coefficient estimates (ITT). Dependent variable: Survey responses to questions about how well the policy of the current government handles societal problems (various dimensions); original answers are in categories from 1 to 4, with 1-2 negative, 3-4 positive; the dependent variable is the share of positive answers (3-4). Data source: Afrobarometer. ART treatment: intent-to-treat regressions of the effect of instruments for ART coverage on violent events, instruments constructed as interactions between HIV prevalence in a region in 2001 and time-varying measures of the median price of ART treatment (ART Price). All specifications as described in equation (2). \*\*\*/\*\* indicate significance at 10%/5%/1%, respectively. Standard errors in parentheses clustered at the sub-national region level.

**Table A38:** INDIVIDUAL TRUST IN INSTITUTIONS

| PANEL A | TRUST IN<br>INSTITUTIONS |  | LAW AND ORDER |
| --- | --- | --- | --- |
|  | Parliament<br>(1) | Local Government<br>(2) | Police<br>(3) |
| $HIV_{r,2001} \times ART_{IV,t}$ | 0.058**<br>(0.024) | 0.039<br>(0.024) | -0.014<br>(0.016) |
| $HIV_{r,2001} \times ART_{IV,t}$ | ART Price | ART Price | ART Price |
| Mean | 0.83 | 0.79 | 0.73 |
| Observations | 294 | 300 | 381 |
| Regions | 112 | 112 | 112 |
| R2 | 0.62 | 0.68 | 0.80 |
| PANEL B | TRUST IN<br>INSTITUTIONS |  | LAW AND ORDER |
|  | Parliament<br>(1) | Local Government<br>(2) | Police<br>(3) |
| $HIV_{r,2001} \times ART_{IV,t}$ | 0.068**<br>(0.028) | 0.055**<br>(0.027) | -0.017<br>(0.020) |
| $HIV_{r,2001} \times ART_{IV,t}$ | ART Cost | ART Cost | ART Cost |
| Mean | 0.83 | 0.79 | 0.73 |
| Observations | 294 | 300 | 381 |
| Regions | 112 | 112 | 112 |
| R2 | 0.62 | 0.68 | 0.80 |
| PANEL C | TRUST IN<br>INSTITUTIONS |  | LAW AND ORDER |
|  | Parliament<br>(1) | Local Government<br>(2) | Police<br>(3) |
| $HIV_{r,2001} \times ART_{IV,t}$ | 0.165***<br>(0.061) | 0.136**<br>(0.056) | 0.082<br>(0.056) |
| $HIV_{r,2001} \times ART_{IV,t}$ | ART Cov | ART Cov | ART Cov |
| Mean | 0.83 | 0.79 | 0.73 |
| Observations | 294 | 300 | 381 |
| Regions | 112 | 112 | 112 |
| R2 | 0.62 | 0.68 | 0.80 |
| *Region fixed effects* | ✓ | ✓ | ✓ |
| *Year fixed effects* | ✓ | ✓ | ✓ |
| *HIV prevalence Trend* | ✓ | ✓ | ✓ |

Notes: Coefficient estimates (ITT). Dependent variable: Survey responses to questions about trust in institutions (various dimensions); original answers are in categories from 1 to 4, with 1-2 negative, 3-4 positive; the dependent variable is the share of positive answers (3-4). Data source: Afrobarometer. ART treatment: intent-to-treat regressions of the effect of instruments for ART coverage on violent events, instruments constructed as interactions between HIV prevalence in a region in 2001 and time-varying measures of the median price of ART treatment (ART Price). All specifications as described in equation (2). \*\*\*/\*\*/\* indicate significance at 10%/5%/1%, respectively. Standard errors in parentheses clustered at the sub-national region level.

## Notes

✯ The authors are grateful for comments by Jeremy Lucchetti, Massimo Morelli, Michele Pellizzari, Lukas Rosenberger, Mathias Thoenig, and Joachim Voth. Support by Alexander Lehner and Alessandro Saia with the implementation of some of the robustness checks is gratefully acknowledged. Dominic Rohner gratefully acknowledges financial support from the ERC Starting Grant POLICIES FOR PEACE-677595. Uwe Sunde gratefully acknowledges financial support by Deutsche Forschungsgemeinschaft through CRC TRR 190 (project number 280092119).

1 See, e.g., the December 2000 AIDS epidemic update by UNAIDS/WHO.

2 The construction of the measure HIV*_geo,_*_16_*_K_* involves a minimum criterion of a population of 16.000 inhabitants per grid cell, which corresponds to the 8^th^ decile of the distribution of population density. In the Appendix, we report results for alternative thresholds.

3 Figure A4 in the Appendix shows the corresponding results when relating ART coverage to HIV prevalence.

4 In fact, an additional analysis documents that the expansion of ART coverage in the context of the HIV/AIDS epidemic in Africa did not lead to a significant improvement in aggregate economic prosperity as measured by income per capita, but it did lead to a significant increase in life expectancy (see Appendix Tables A21–A23). This implies that it is unlikely that the main findings are driven to a large extent by overall economic prosperity. A similar conclusion emerges from the earlier results from extended specifications that account for international aid, institutional quality, and economic development (see Table A16).

5 Unreported estimates for trust in other institutions, like the president, the ruling party, or the electoral commission reveal similar findings.

6 The difference between observed and predicted values is plotted as orange bars.

7 The estimates are based on ART coverage relative to the population, as in Appendix Table A17. Qualitatively similar, but quantitatively larger effects are found when using ART coverage relative to HIV prevalence as in the baseline estimates. Details are available upon request.

* Further information can be obtained from: https://www.unaids.org/en/dataanalysis/knowyourresponse/HIVdata_estimates.

† The analysis focuses on first line regimens for adults, which represent the most widely used treatments of the time and were substantially less expensive than second line treatments. This particular first line regimen is considered the most effective regimen and is the one recommended by WHO. Furthermore, because of its higher initial price it was initially not widely used in low and middle income countries. Alternative regimens (in particular the second first line regimen ZDV-3TC-NVP) are used for robustness checks on the instrumentation.

‡ In particular, the price index for year *t* is constructed as *price*_*index_t_* = (*price*_2003_*/price_t_*) *−* 1.

§ In particular, the cost index for year *t* is constructed as *cost*_*index_t_* = (*cost*_2003_*/cost_t_*) *−* 1.

¶ Specifically, the construction uses data on the price for the first line regimen ZDV-3TC-EFV in 2003, *P*_0_, and the average price in the years from 2015 to 2017, *P_lim_*, which corresponds to the minimum price since price and production costs for the ZDV-3TC-EFV regimen effectively stabilized after 2015. With a mark-up in the first year given by *M*_0_ = *P*_0_ *− P_lim_*, the synthetic price index for the subsequent years is computed as 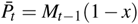, where *x ∈* (0, 1) denotes a fixed proportional reduction in the mark-up in each year, and with the mark-up each year given by 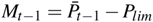. As baseline, the mark-up is assumed to be reduced by 15 percent per year (*x* = 0.15); robustness checks are conducted for *x* = 0.2 and *x* = 0.25. For comparability with the series of actual prices and costs, the synthetic price index for year *t* is computed as 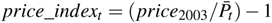.

|| (https://www.gatesfoundation.org/How-We-Work/General-Information/Grant-Opportunities).

